# SSRI treatment of obsessive-compulsive disorder as motion across a bistable fold: A calibrated circuit–plasticity model of response, remission, and augmentation

**DOI:** 10.64898/2026.07.24.26358842

**Authors:** Sankaran Sundaresan

## Abstract

We present a model for an OCD patient’s response to SSRI treatment that combines serotonin pharmacokinetics, a basal-ganglia–thalamocortical circuit, and corticostriatal plasticity, read out as a Y-BOCS score. Within it, that response is dictated by a plasticity-dynamics equation in which the OCD state is an elevated fixed point, separated from the healthy state by a saddle. This healthy-saddle-OCD structure is what leaves patients stuck. Effective treatment annihilates the saddle and the OCD fixed point together, making full remission possible. A response can occur even when annihilation does not, but those patients relapse once treatment is withdrawn. Calibrated to SSRI trial trajectories, the model shows that SSRIs achieve annihilation in mild-to-moderate OCD but not the most severe. Antipsychotic and memantine augmentation enter the dynamics at points distinct from the SSRI, so the model predicts they combine to annihilate the attractor an SSRI alone cannot, a rationale for augmentation in severe OCD.

## 1 Introduction

Psychiatric drugs act across scales that are usually modeled apart. A compound’s pharmacokinetics set receptor occupancy; occupancy perturbs a neural circuit; the circuit’s plasticity lets that perturbation accumulate over weeks; and only at the end of the chain does a clinician read a symptom score. Quantitative accounts of treatment usually live at one level: statistical dose– response curves with no mechanism, or biophysical circuit models with no link to drug exposure or the clinical rating. The questions clinicians actually ask, why a patient responds, plateaus, or briefly worsens, and what to add when the first drug fails, are answered in the coupling between these levels, which is rarely built.

Obsessive-compulsive disorder is a clear case of this gap. SSRIs are the pharmacological first-line, and the response they produce is characteristic but only partly explained: it is partial, it builds over weeks rather than days, and it improves little once the dose passes a threshold. We will argue that it is also gated by baseline severity. That prediction runs against the strongest current evidence ^1^, which finds the SSRI-versus-placebo response nearly independent of severity; the same mismatch appears in every model we analyzed, bistable and fold-free alike, so it is a feature of the model class rather than a verdict on bistability (S12). Roughly 40–60% of patients respond inadequately^2^, and treatment then turns to augmentation with antipsychotics or glutamatergic agents, chosen largely by trial and error ^3,4^. No account has yet tied these observations together: the transporter occupancy the drug produces, the corticostriatal circuit it perturbs, the plasticity that fixes the disease state, and the Yale–Brown Obsessive-Compulsive Scale (Y-BOCS; ^5^) that measures response.

A useful way to think about such transitions has taken hold in psychiatry over the past decade. A disorder is treated not as a fixed label but as one of several stable states of a nonlinear system, separated by basins, so that treatment changes the state landscape rather than nudging a symptom score in proportion ^6–8^; dynamical-systems models have begun to build this picture for OCD circuitry itself ^9^. The view is measurable: as a basin flattens near a transition, recovery from perturbations slows, so rising autocorrelation and variance in densely sampled symptom series serve as early-warning signals, seen before depressive transitions in real patients^10^. For OCD, attractor-network accounts cast the disorder as an over-stable corticostriatal state ^11^; a Wilson–Cowan model of the cortico-striato-thalamo-cortical circuit ties OCD hyperactivity to the excitation/inhibition balance of striatal projection neurons ^12^; and a frontostriatal model fit to imaging places OCD at one of a pair of saddle-node attractors ^13^. These models establish that the circuit can be bistable, but each treats the bifurcation control as an abstract parameter: none is coupled to a pharmacokinetic front end, transporter occupancy, or the clinical dose–response, so none can say how a particular SSRI at a particular dose moves the landscape.

Here we build that coupling for OCD. The model combines a mean-field basal-ganglia–thalamocortical circuit ^14^, a Bienenstock–Cooper–Munro ^15^ (BCM) corticostriatal plasticity rule referenced to the healthy baseline firing rate (so the healthy state is a fixed point by construction), and an extracellular-serotonin engine with transporter and autoreceptor pharmacokinetics, read out onto the Y-BOCS. In this assembly the OCD state is a stable, elevated fixed point whose severity is set by a single per-patient plasticity gain. Serotonin tunes a plasticity sink, so an SSRI moves that fixed point rather than merely damping activity: treatment is the motion of a *bistable* fixed point, and remission is its annihilation at a saddle-node fold, the point at which the OCD state ceases to exist. The result is a single calibrated model that runs from receptor occupancy, across real drugs and doses, through the circuit and its weeks-long plasticity, to the rating scale; it makes response, non-response, onset, and relapse features of one landscape. That lets us ask a sharp question: does SSRI treatment cross a bistable fold, or slide along a graded response?

Calibrated to SSRI trial trajectories, the model reproduces the severity-graded response and its weeks-long onset. Because the disease state is set by plasticity rather than by the serotonin level, effective augmentation should act on the plasticity balance; the antipsychotics and glutamatergic agents already used clinically map onto distinct “routes to the fold.” Comparing the fold against a matched graded alternative, we find the two indistinguishable on the population means, while the fold predicts individual-level signatures a graded model cannot produce (§4.1).

## 2 The model

We represent OCD treatment response as a single system coupling four layers: (i) an *extracellular-serotonin engine* in which an antidepressant’s transporter occupancy sets serotonin dynamics (Methods, Eqs. 3–4); (ii) a mean-field *cortico–basal-ganglia–thalamic circuit* (Eq. 2) whose caudate output encodes symptom severity; (iii) a slow, activity-dependent *remodeling of the corticostriatal synapse* over weeks (Eq. 5); and (iv) an *observational readout* onto the clinical score (Eq. 6). A single quantity is the model’s disease-severity coordinate: the *excess corticostriatal synaptic weight G*_CS_ ≥ 0, the corticostriatal projection strength above the healthy circuit. *G*_CS_ = 0 is the healthy state, and the larger *G*_CS_, the more severe the OCD; it raises caudate firing and, through the readout, the model score *Y*_model_ (Fig. 1). Symptoms are read from the circuit through an explicit observational law, and the expectation (placebo) response enters at that readout only, not the synaptic dynamics. Each component is fixed from the literature or calibrated to trial time-courses; the governing equations are collected in Methods (§5.1) and justified, with all parameter values, in S1–S2.

**Figure 1.**
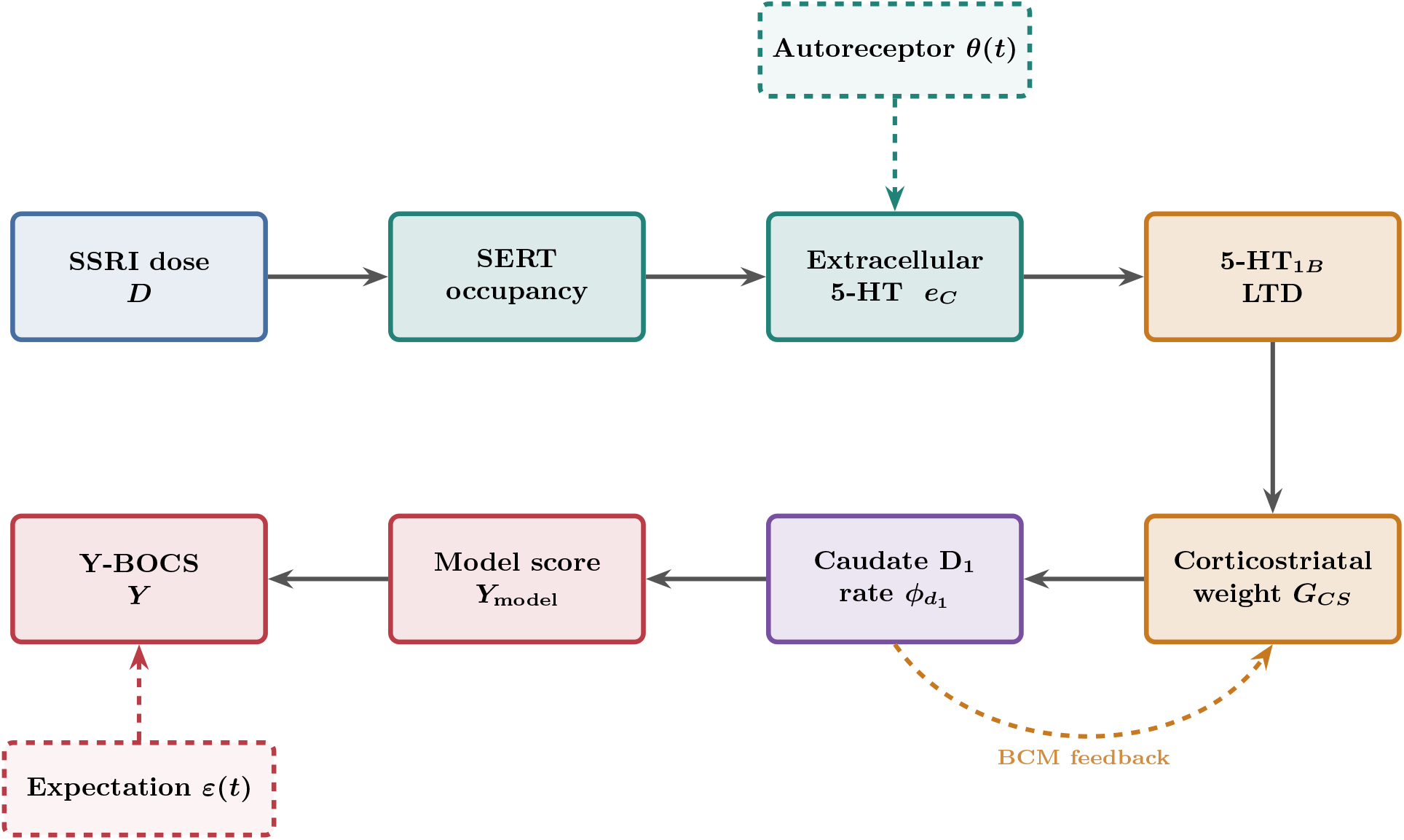
The treatment cascade as a multi-scale signal chain. An administered SSRI dose *D* sets fractional SERT occupancy, which lowers reuptake and raises extracellular serotonin *e*_*C*_; the rise is gated by a serotonin autoreceptor *θ*(*t*) that desensitizes over weeks (dashed, top). Elevated *e*_*C*_ drives presynaptic 5-HT_1*B*_ long-term depression of the corticostriatal synapse, relaxing its effective weight *G*_CS_ (the model’s disease-severity coordinate: *G*_CS_ = 0 is healthy, larger *G*_CS_ is more severe OCD); *G*_CS_ sets the caudate *D*_1_-MSN firing rate 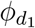 through the mean-field basal-ganglia–thalamocortical circuit. The observational law maps 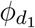 to the model score *Y*_model_ (the pure circuit prediction); the observed Y-BOCS is *Y*_model_ minus the transient expectation (placebo) offset *ε*(*t*), which enters only at this readout step (dashed, bottom-left). The circuit output feeds back onto the plasticity through the bidirectional (BCM) source term (dashed), which is what makes the untreated state bistable. Box color denotes the modeled scale: dose (blue), serotonin (teal), plasticity (amber), circuit (violet), and readout (rose); symbols and governing relations are defined in §2–2.1.

### 2.1 Observational readout and model attributes

The Y-BOCS (0–40; ^5^) is a *behavioral* rating (the endpoint of the causal chain and the quantity a clinician actually records), whereas the circuit outputs *firing rates*. No first-principles law converts a caudate firing rate into a symptom count, so the two cannot be identified directly. Yet any model meant to speak to treatment must connect them. We therefore posit an explicit observational law (Methods, Eq. 6) that maps the caudate *D*_1_ rate 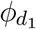 to a model score *Y*_model_, our proxy for the Y-BOCS (Fig. 2); which rate is mapped is not load-bearing, since the circuit’s rates rise together (S3), and the results are robust to the assumed *D*_1_:*D*_2_ overdrive ratio (S21).

**Figure 2.**
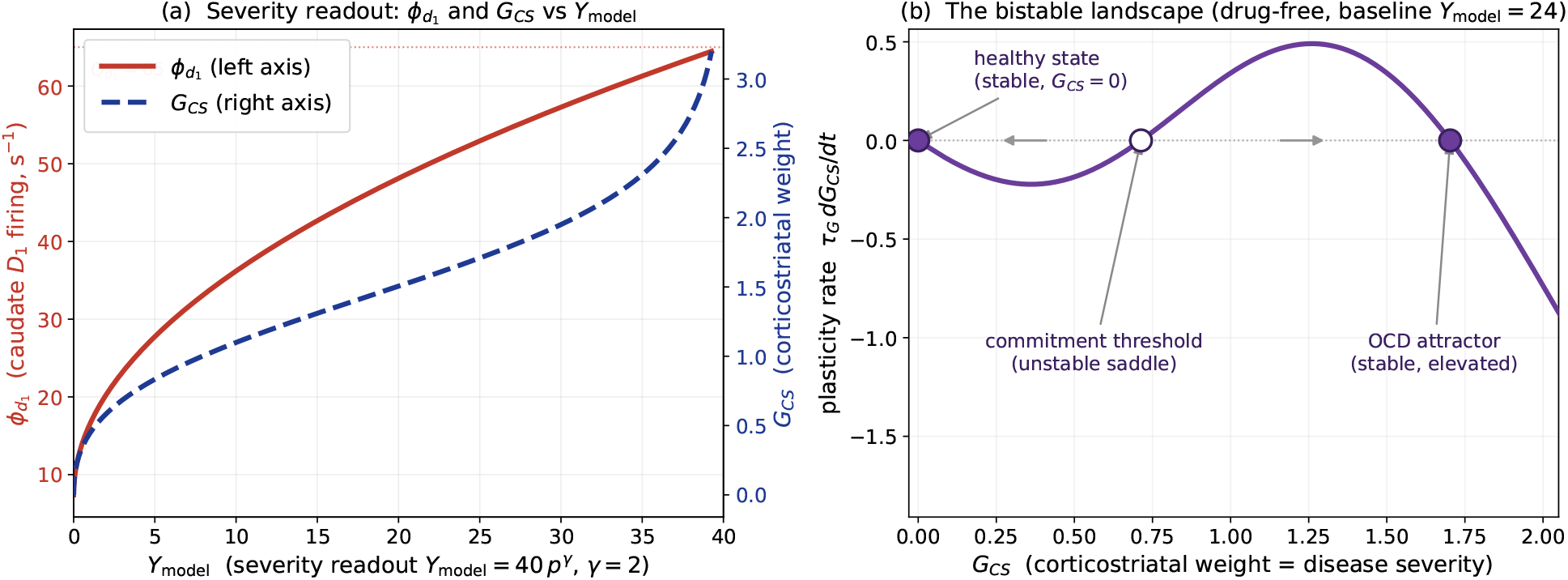
*(a)* The observational readout mapping circuit state to symptoms: caudate *D*_1_ firing 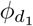 (red, left axis) and the corticostriatal weight *G*_CS_ (blue, right axis) as one-to-one functions of 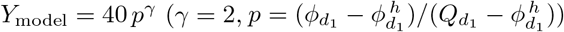. The model score reads back to an underlying firing rate and synaptic weight; near the top of the scale *G*_CS_ rises steeply as 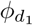 approaches its ceiling 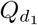. *(b)* The bistable landscape the plasticity rule (Methods, Eq. 5) produces, drug-free, for a moderate patient (*Y*_model,0_ = 24): the plasticity rate *τ*_*G*_ *dG*_CS_/*dt* against *G*_CS_ crosses zero at three fixed points: the *healthy state* (*G*_CS_ = 0, stable, filled), the *commitment-threshold saddle* (unstable, open; the subclinical/clinical divide), and the elevated *OCD attractor* (stable, filled). Below the saddle the weight relaxes to health, above it to the OCD state (axis arrows). Treatment steepens the sink and slides the OCD attractor leftward until it collides with the saddle and both vanish: the saddle-node fold that is remission. Population firing rates versus *G*_CS_ (the choice and grounding of the severity coordinate) are shown in S3.

This circuit-to-clinic step is the recognized difficulty in relating brain models to psychiatric outcome, and it has been taken before. Attractor-network and frontostriatal accounts read OCD severity off an over-stable ^11^ or bistable ^13^ corticostriatal state, while a separate line treats the rating scale itself as the observable of a dynamical or statistical process ^16–18^; each makes some version of the commitment we make here, that a clinical score can serve as the readout of a brain-level state. We adopt it openly, with its caveat: because the law joins a firing rate to a behavioral test score, it is inescapably *qualitative*, not a metrologically exact conversion. Our quantitative calibration is aimed accordingly: at getting the *landscape* right (the gradient of outcome with dose and severity, the location of the fold, the durability of remission) rather than at point accuracy on the 40-point scale.

Two properties govern how this readout meets data. First, built from the caudate *D*_1_ firing rate through a fixed power law, it tracks Y-BOCS in direction and gradient but not point for point. Second, because every calibration target here is a net, drug − placebo difference, any fixed offset between *Y*_model_ and Y-BOCS cancels and never enters the fit. The one comparison in which the absolute level of *Y*_model_ matters is the population responder fraction (S10), where we accordingly read the model as a trend across baseline severity and report the registration offset explicitly.

Three attributes give the model its behavior. First, the slow plasticity of *G*_CS_ (Methods, Eq. 5) is *bistable*: a healthy state (*G*_CS_ = 0) and an elevated OCD attractor coexist, separated by an unstable saddle whose calibrated location marks the subclinical/clinical boundary (the three fixed points are the zeros of the plasticity rate 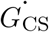; Fig. 2b). Because *G*_CS_ remodels under a bidirectional (Bienenstock–Cooper–Munro) rule with a homeostatic zero at the healthy firing rate, health is a fixed point and the untreated disease is a genuine alternative stable state, not a mere elevation. The bistability follows structurally from the source’s three ordered roots and survives changes to its functional form, so it is posited from grounded plasticity rather than tuned to the data (S4). We hold the interior modification threshold *θ*_*M*_ *constant* rather than letting it slide with activity: corticostriatal plasticity runs on two dopamine-gated thresholds (Shen et al. ^19^), and in single-neuron learning models the metaplasticity that slides them stabilizes individual synaptic *weights* rather than the network state (Khodadadi et al. ^20^; Trpevski ^21^); whether a sliding threshold would instead stabilize or erode the disease attractor is taken up, with its evidence grade, in S5.1.

Second, treatment lowers the OCD attractor. Raising serotonin steepens the plasticity’s depression sink, so a patient driven past the saddle *remits* into the healthy basin, whereas one driven only part-way *improves* on a lowered OCD branch. Remission is thus gated by baseline severity through the per-patient gain *α*, while a ≥ 35% response can be met without crossing (response and remission are established as distinct clinical endpoints in OCD; ^22^). Because *α* is set from presenting severity and *γ* is fixed so the fold coincides with the remission threshold, the *existence* of this gating is in part built in; its *quantitative* content (the steepness of the remission-versus-severity relation and the threshold location) is the empirical, falsifiable prediction (§4.5).

Third, a clinical arm is a *population* of baseline severities, integrated over the enrolment Y-BOCS distribution (Methods, §5.3); the expectation (placebo) response is carried at the readout only and cancels in the net drug − placebo quantity used throughout. The model is deliberately *parsimonious*: nearly every constant is fixed from the literature or the circuit, and the net trajectories identify only two slow couplings: the 5-HT_1*B*_ drug coupling *b* and the autoreceptor coupling *κ*_des_ (Methods, §5.4; S4).

## 3 Results

### 3.1 The plasticity gain *α*: vulnerability and the making of OCD

In the model, the patient-specific plasticity gain *α* scales the potentiating plasticity source: it is the one locus of individual vulnerability, set by a patient’s genetics and by the cumulative burden of stressful experience. It is a slow, structural parameter, distinct from the fast dynamical state it governs, the corticostriatal weight *G*_CS_. The model does not itself generate the accrual of *α*; that accrual enters as an input.

For an unmedicated individual (serotonin at its untreated level), the right-hand side 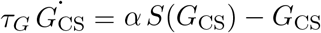 can be drawn as a function of *G*_CS_ for any fixed *α* (Fig. 3a). Below a threshold vulnerability *α*_*c*_ the only fixed point is health (*G*_CS_ = 0), and it is *globally* stable: any elevation relaxes back over a few times the remodeling time *τ*_*G*_. For such a person OCD is dynamically impossible. As *α* accrues past *α*_*c*_, a saddle-node fold appears: an unstable fixed point, the *commitment threshold*, and an elevated OCD attractor are born together, near *Y*_model_ ≈ 12 (Fig. 3b).

**Figure 3.**
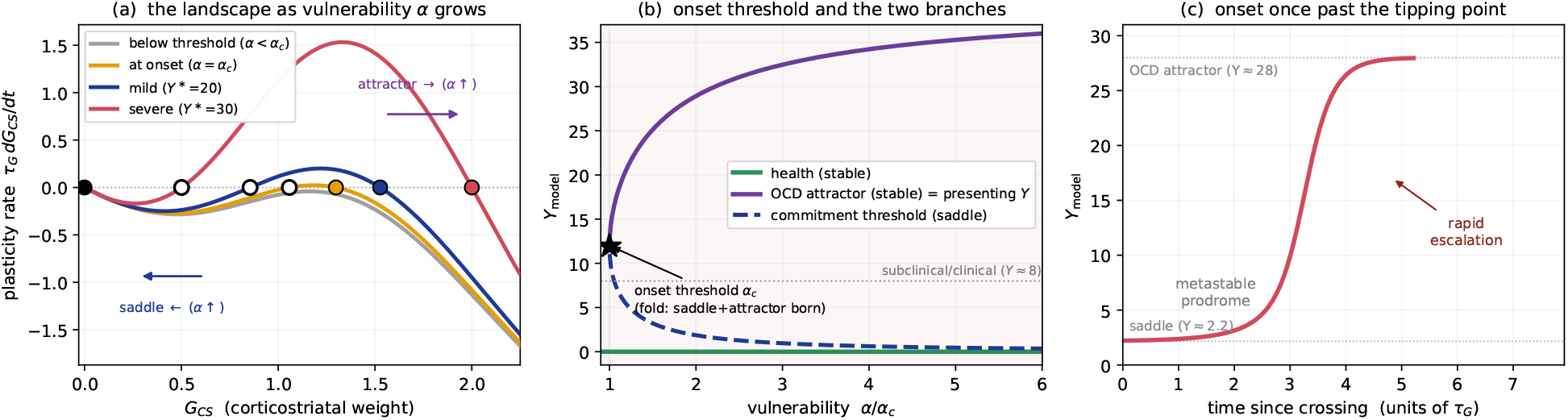
The plasticity gain *α* sets vulnerability, and presenting severity calibrates it. Filled markers are stable fixed points, open markers the saddle; the healthy state (black) lies at *G*_CS_ = 0. *(a)* Drug-free plasticity rate 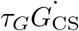 versus *G*_CS_ as *α* increases. Below threshold (gray) the only fixed point is health; at onset (*α* = *α*_*c*_, orange) a saddle and an OCD attractor are born together near *Y*_model_ ≈ 12; for larger *α* (mild, blue; severe, red) the saddle slides left and the attractor slides right. *(b)* The saddle-node bifurcation in *α*: health (green) is stable for every *α*, while the commitment threshold (blue, saddle) and the OCD attractor (purple, stable) are born together at *α*_*c*_ and separate as *α* grows. For *α < α*_*c*_ OCD is dynamically impossible. The attractor branch is the patient’s presenting severity, hence the target from which *α* is calibrated. *(c)* The onset transient for a predisposed patient (*Y*_model,0_ = 28) nudged just past its saddle (drug-free): a metastable prodrome near the subclinical/clinical boundary, then rapid escalation to the OCD attractor. The two plateaus are the saddle and attractor branches of *(b)*. Time is in units of *τ*_*G*_, so the shape (long prodrome then abrupt escalation) is parameter-free; only its *absolute* duration scales with *τ*_*G*_ (≈ 12 weeks, the value that also sets the SSRI onset). Panels use the LTP/LTD crossover *θ*_*M*_ = 9.15 s^−1^ (the interior zero of the source; §2), for which the onset threshold is *α*_*c*_ ≈ 3.4 × 10^−6^ s^4^.

After this onset the geometry determines what follows. So long as *G*_CS_ stays below the commitment threshold it still relaxes to health; two things carry a person across it. An acute stressor lifts *G*_CS_ toward the threshold, while continued accrual of *α* lowers the threshold and deepens the well (in Fig. 3a the saddle slides left and the attractor slides right as *α* grows). As the health basin shrinks, an ever smaller trigger suffices; equivalently, a descending threshold can overtake a *G*_CS_ still relaxing from an earlier perturbation. Health at *G*_CS_ = 0 is never itself overtaken, so onset requires both a raised gain and a trigger. Once across, the potentiating source takes over and the weight climbs to the elevated attractor: clinical OCD. Should *α* rise further, and compulsive behavior could itself raise it, the threshold recedes and the attractor deepens, so recovery grows ever less likely and the score climbs.

This is what the calibration rests on. A patient sitting at a steady, elevated Y-BOCS has settled at the particular *α* whose attractor lies at that score; inverting the statement, a patient’s presenting severity fixes the model’s *α*. That is exactly how we set it: one *α* per patient, read from baseline severity (Fig. 3b, where the attractor branch *is* presenting severity). With *α* so defined, we turn to calibrating the shared parameters against trial trajectories.

### 3.2 A single shared plasticity coupling reproduces six SSRIs

Rather than fit each drug in isolation, we fix some 35 constants from the literature or the circuit and assign every drug its SERT occupancy from independent PET/SPECT imaging (Table S1), exposing only *two* shared parameters to the trial data: the corticostriatal-plasticity coupling *b* and the autoreceptor coupling *κ*_des_. The readout, its threshold, and the per-patient gain are all set outside the drug fit (degrees-of-freedom accounting in Methods). Evaluated at the imaging (literature) *D*_50_, these two constants reproduce the net (drug − placebo) Y-BOCS response across thirteen dose-arms of six SSRIs at an RMS of 0.83 Y-BOCS points over all 84 arm-timepoints, capturing both the dose gradient and the weeks-long onset (Fig. 4). The net response is Δ*Y*_model_, since the expectation offset cancels in the difference. A single systematic departure survives, in the fluoxetine dose-ordering (Fig. 4a): the 20/40/60 mg rank is recovered but the 60 mg increment is under-shot, because SERT is already 88–96% occupied at these doses and the serotonergic drive has saturated, a ceiling on the drive rather than a mis-estimated potency.

**Figure 4.**
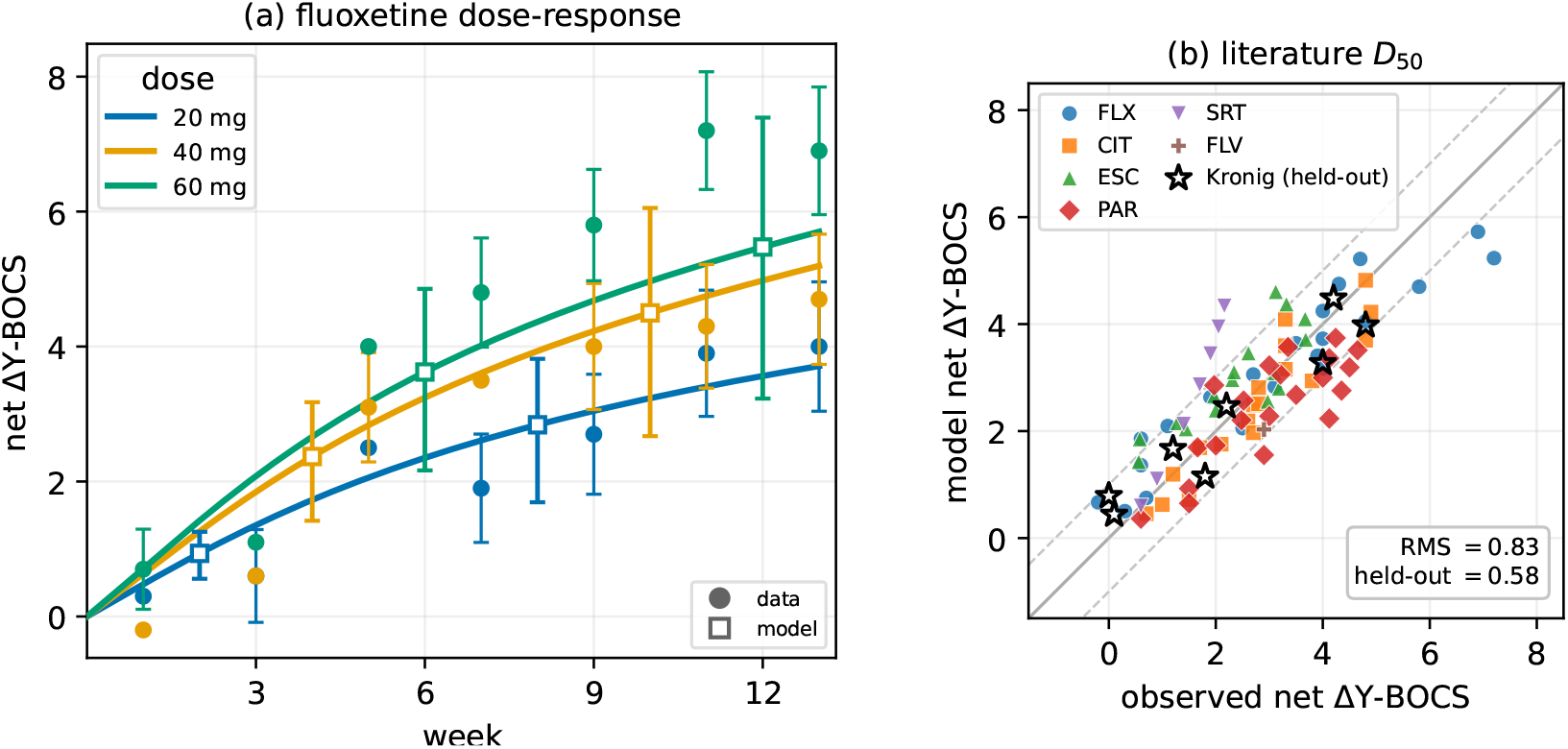
One shared plasticity coupling reproduces the SSRI dose- and time-course of response across six drugs. *(a)* Fluoxetine dose-response^25^: net (drug−placebo) Y-BOCS trajectory for 20, 40 and 60 mg. Solid curves are the model at the calibrated coupling *b*; filled circles are trial means (± standard error (SE) bars at a staggered subset of weeks). Open squares are the model at even weeks, their *asymmetric* bars spanning the *b*-confidence interval: asymmetric because *b*’s profile-likelihood interval is wider toward higher *b* (where *κ*_des_ compensates; Fig. S4) and because near the fold a stronger drive tips more of the baseline distribution across the saddle-node. The model captures the dose-*ordering* but under-predicts the 60 mg increment: across 20–60 mg SERT is already 88–96% occupied, so the drive saturates and cannot generate the full high-dose benefit (revisited in the Discussion). *(b)* Model vs. observed net Y-BOCS at all 84 calibration arm-timepoints (colored by drug) at the literature *D*_50_; dashed lines mark ±1 Y-BOCS, the median per-point SE. The two shared couplings (*b* and *κ*_des_) reproduce all six SSRIs (RMS 0.83; every digitized trajectory and its trial source is tabulated in S22). Black stars are an *out-of-sample* sertraline trial not used in calibration ^24^, predicted by the frozen model at held-out RMS 0.58. The *D*_50_-optimized fit is given in S4.1; *the calibration retains the measured (literature) D*_50_ *throughout*.

The two sertraline trials in the corpus lie beyond what any single-potency account can span. The calibration arm ^23^ and an independent, held-out trial ^24^ diverge sharply enough that no common occupancy accommodates both: at the imaging *D*_50_ the frozen model tracks the held-out data of Kronig et al. ^24^ closely (RMS 0.58) while fitting Greist et al. ^23^ only loosely (RMS 1.34). Improving the Greist fit would require depressing sertraline’s *D*_50_ to ∼ 26 mg, a value irreconcilable with the measured PET occupancy and one that surrenders the Kronig prediction. We retain the imaging *D*_50_ throughout and read Greist as the low outlier, an assignment the held-out Kronig arm reaches independently (Fig. 4b, stars). Greist remains in the calibration set, so the label is interpretive rather than an exclusion, and a leave-one-arm-out cross-validation (mean held-out RMS 0.81 vs in-sample 0.83) confirms no single arm drives the fit (S4.2).

The remaining held-out arm departs in the opposite direction, and by construction: the frozen model *under* -predicts clomipramine (2.2 against an observed 10 Y-BOCS points) because, as a mixed serotonin–norepinephrine reuptake inhibitor, it must exceed any transporter-selective account by the increment its noradrenergic action contributes. A SERT-only model is thus *obliged* to fall short here, a specificity test passed, not a calibration failure.

The identifiability of the two coupled constants, and the uncertainty they propagate, are developed in S4 and S4.1.

### 3.3 Treatment across the fold: remission and response

With the two couplings fixed, the model’s account of treatment is a statement about the geometry of the slow dynamics, not a further fit. The untreated OCD state is one of two stable attractors of the corticostriatal-plasticity equation, separated by an unstable saddle, its position set by the patient’s *α* (Fig. 3b). Raising serotonin steepens the depression sink, so as the dose climbs the OCD attractor and the saddle move together and, at a critical SERT occupancy, collide and annihilate; with the elevated state gone, only health remains and the patient is carried to remission. Treatment is therefore the motion of a fixed point across a saddle-node fold, a geometry that follows from the plasticity rule (§2.1) rather than from the data.

To speak precisely about remission and response we fix two notions of the treated *endpoint*. The *attractor endpoint* is the *t* → ∞ outcome: the value *G*_CS_ settles to once the medication has driven it to the attractor set by that steady dose. The *finite-horizon endpoint* is the value of *G*_CS_ (equivalently *Y*_model_) reached at the end of a treatment protocol of finite duration (weeks, as in a clinical trial), and it depends on how long treatment has run. The attractor endpoint tells us what is *possible*; the finite-horizon endpoint tells us what is *achieved*. Consider first what is possible.

Whether the fold is reached at a tolerable dose depends on how far the attractor begins from it: at typical SSRI efficacy a mild patient (*Y*_model,0_ = 20) folds at fractional SERT occupancy *u* ≈ 0.82, a moderate one (24) only at near-maximal occupancy (*u* ≈ 0.97), and a severe one (28) at no attainable dose (Fig. 5a–c). The robust content is the ordering (mild folds, severe does not), not the precise occupancies or its slope; the steepness of this gating in fact over-predicts mild-patient benefit relative to the largest meta-analysis (§3.5). Even so, a patient who does not cross the fold can still *improve* along the OCD branch: as the dose rises the OCD attractor itself descends (the family of attractors in Fig. 5b), and if the attainable drop in *G*_CS_ (set by the distance between the untreated (*u* = 0) attractor and the treated one) exceeds a stipulated response threshold (typically a ≥ 25–35% Y-BOCS reduction), *response* is attainable with sustained treatment. But such improvement is drug-held: discontinuation returns the OCD attractor to its *u* = 0 position and relapse is inevitable. Within this model, then, patients with *Y*_model,0_ ≳ 24 (for *u* capped at 0.95) can at best be *responders who relapse* on discontinuation. (Placebo and psychotherapy are omitted here.)

**Figure 5.**
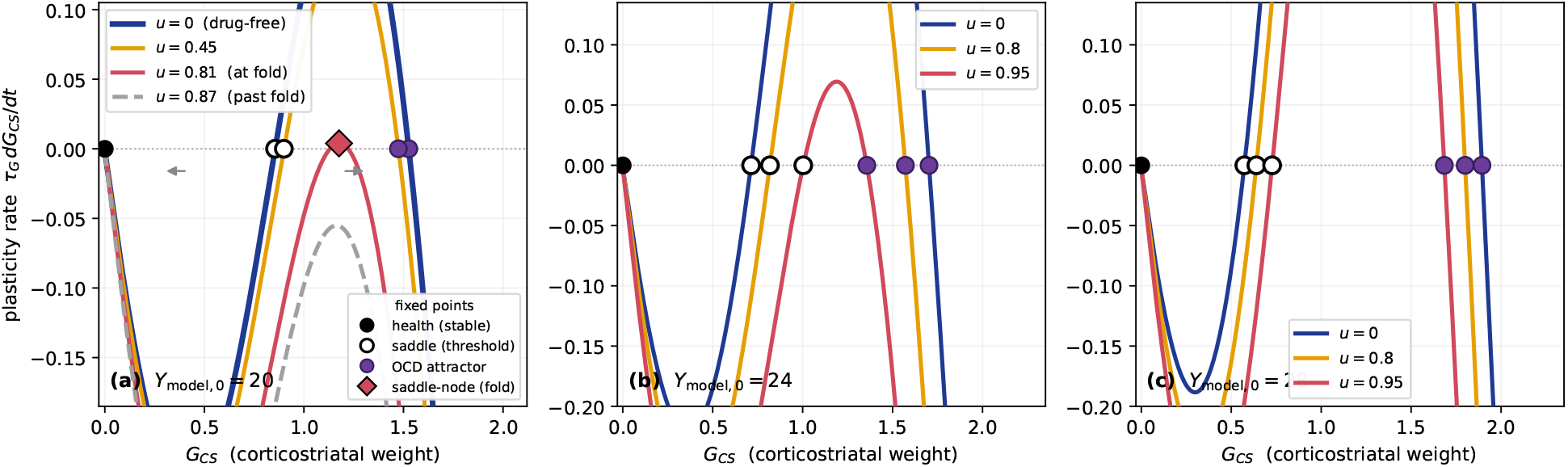
Treatment is the motion of a bistable fixed point, and baseline severity sets whether the fold is reachable. Each panel plots the corticostriatal-plasticity rate *dG*_*CS*_/*dt* (right-hand side of the plasticity equation) against the corticostriatal weight *G*_CS_; zeros of a curve are fixed points of *G*_CS_. Throughout, the *black filled circle* is the healthy stable state (*G*_CS_ = 0, *Y*_model_ = 0); the *open circle* is the saddle, the separatrix between spontaneous recovery and progression, at *Y*_model_ ≈ 6, the subclinical/clinical boundary; and the *purple filled circle* is the elevated, stable OCD attractor (at rest equal to the panel’s baseline by construction). Dose enters only through the fractional SERT occupancy *u* = *D*/(*D* + *D*_50_); as *u* rises, extracellular serotonin *e*_*C*_ increases and steepens the linear sink, tilting each curve downward. *(a)* A representative patient at baseline *Y*_model_ = 20, at four doses; gray arrows mark the two basins. As the dose rises the saddle and OCD attractor approach and, at the *saddle-node fold* (*u* ≈ 0.82, red; diamond marks the merged pair), annihilate, leaving only health: remission. *(b)* At *Y*_model_ = 24 the attractor sits deeper and reaches the fold only at near-maximal occupancy (*u* ≈ 0.97): at *u* = 0.95 (red) the saddle and attractor are all but merged (on the verge of remission). *(c)* At *Y*_model_ = 28 the attractor barely moves and stays far from the saddle even at *u* = 0.95, so no attainable dose folds it: serotonergic monotherapy cannot carry this patient to *remission*, though the attractor still descends (a graded *response* without crossing, the response/remission split of §3.3) (*Y*_model_ = 28 maps to Y-BOCS ≈ 24–25 under the registration offset, a moderate-to-severe rather than extreme patient). Panels (b,c) show *u* = 0, 0.8, 0.95. Whether an SSRI drives a patient to remission is thus set by how far the OCD attractor starts from the fold, i.e. by baseline severity (Supplementary Fig. S4); the descent of the saddle with severity and the onset that follows are taken up next (§3.4).

A milder patient (*Y*_model,0_ = 20; Fig. 5a) can instead be carried to *remission* once *u* > 0.82. Whether such a patient then stays well or relapses on discontinuation turns on how far along the descent they have travelled. If their *G*_CS_ has dropped below the *untreated* (*u* = 0) saddle during treatment, they are durably remitted: off drug they continue toward health (*G*_CS_ = 0), more slowly than on drug, and relapse is possible only under a large external trigger, not inevitable.

What a finite course of treatment *achieves*, by contrast, depends on how long it has run, because the corticostriatal descent is slow (of order *τ*_*G*_, weeks). A patient who *can* remit (whose treated attractor is health) may not have descended below the untreated saddle within a short trial, and sopresents at that horizon as a responder rather than a durable remitter; sustained treatment carries them across, a timing distinction that returns at onset (§3.1) and at relapse (§3.4).

### 3.4 Distinguishing the fold from a graded response

To test the paper’s central question (should SSRI treatment be viewed as crossing a bistable fold or sliding along a smooth, graded response?), we built a matched graded-null model: the identical assembly with the fold-creating source replaced by a monotone one (the textbook indirect-response form), refit to the thirteen arms, each with its own drug coupling. It reproduces the means as well as the bistable model (RMS 0.90 vs 0.83 Y-BOCS points, statistically indistinguishable; S6). The fold is no special ingredient; it is what a baseline-referenced Bienenstock–Cooper–Munro plasticity source generically produces once read through the saturating circuit, present even for a threshold-free Hebbian rule, and reached as a graded null only by abandoning plasticity itself (S8). The population mean is thus a degenerate observable that two mechanistically opposite accounts fit alike; the distinction lives in what the mean averages away: *who* remits, and in what pattern. Four categorical, individual-level signatures separate the fold from a graded account.

Two are *attractor-endpoint* signatures, read off the *t* → ∞ outcome of §3.3. First, individual remission is *discontinuous* in dose: as occupancy rises the endpoint falls smoothly for the graded model but drops abruptly at the saddle-node for the fold (Fig. 6A). Second, a treated cohort’s endpoint distribution becomes *bimodal* (a remission cluster plus a residual OCD mode; Fig. 6C), a prospective, powered test (S6). Both are sharpest at the attractor and attenuate at a finite trial horizon, so neither is yet established in data. The other two signatures are visible at finite time.

**Figure 6.**
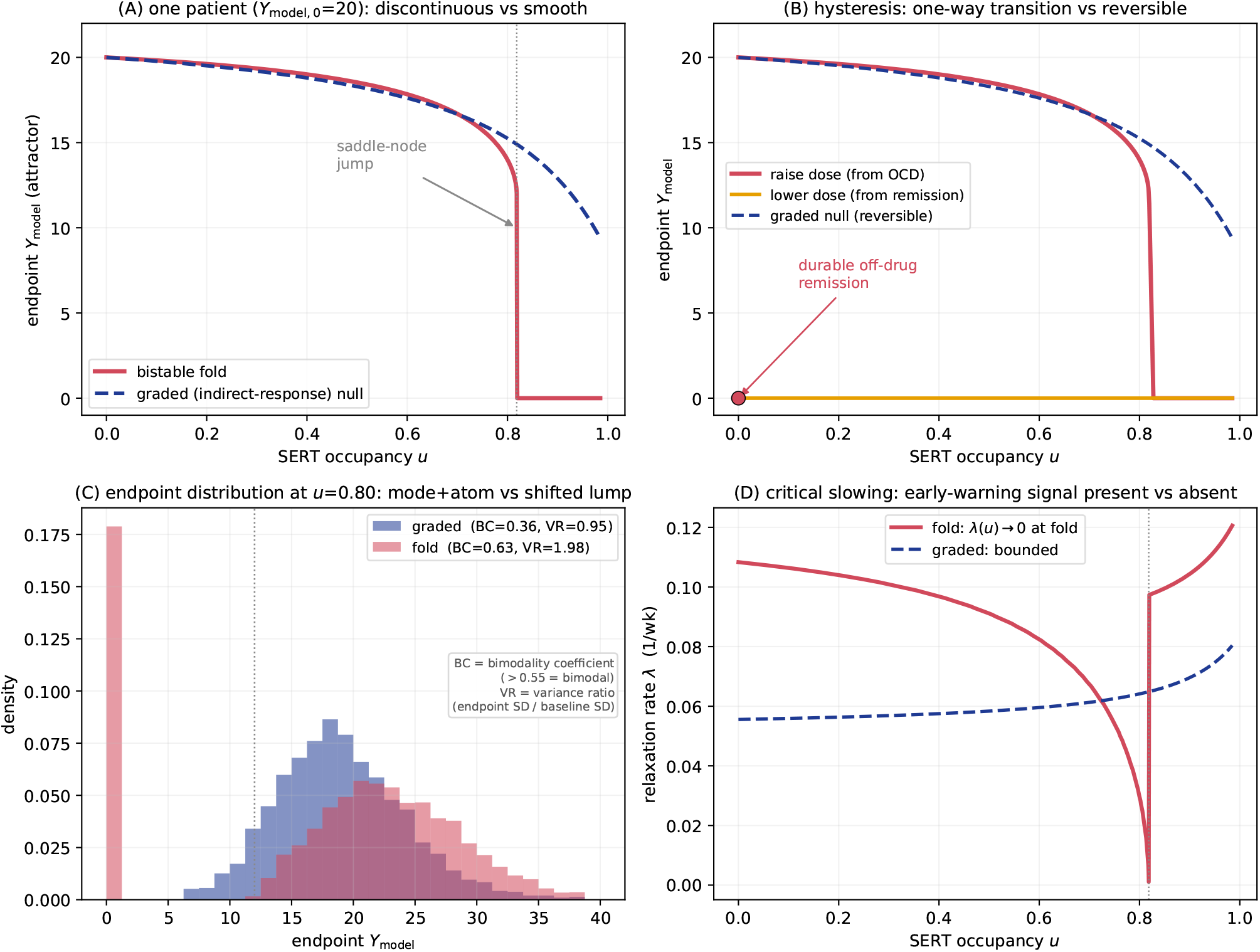
The bistable fold versus a matched graded model. A graded (indirect-response) model built from the identical pharmacokinetic and circuit machinery, with the fold removed, is refit to the same SSRI trajectories; it matches the population means (S6) yet diverges on four observables: two decisive, data-backed signatures *(B,D)* and two sharper predictions that develop only over long times *(A,C). (A)* Individual endpoint falls *discontinuously* at the saddle-node as occupancy *u* rises (fold, red) versus a smooth decline (graded, blue). *(B)* **Irreversibility**. Raising then lowering the dose: the fold shows a one-way transition with *durable off-drug remission* while the graded model is fully reversible. *(C) Simulated* treated-cohort endpoint distribution at the attractor: the fold produces a residual mode plus a remission atom (BC= 0.63, VR= 1.98) versus a single graded lump (BC= 0.36, VR= 0.95); at a realistic trial horizon this split is weaker (S6). *(D)* **Critical slowing**. Recovery rate *λ*(*u*) falls to zero at the fold (early-warning signal) but stays bounded for the graded model.

The first is *irreversibility*. Relapse-prevention trials measure exactly the remission/response distinction of §3.3, and two have addressed it directly. Fineberg et al. ^26^ and a nine-trial meta-analysis (Kishi et al. ^27^, *n* = 1084) took SSRI responders, continued or withdrew the drug, and followed relapse. Both demonstrate the same pattern: after withdrawal responders do *not* all relapse (about half remain well off drug over six months), so the time-to-relapse curve falls and then *plateaus* rather than decaying to zero, while continued medication holds relapse lower still. A durable off-drug remission subgroup, and a plateauing relapse curve, are what these trials reveal.

We do not attempt a quantitative match: the durable fraction the model predicts moves substantially with the placement of *θ*_*M*_, with how the expectation (placebo) response is represented, and with the firing-rate-to-Y-BOCS readout, none pinned tightly enough for the number to be meaningful (S7). We make only the limited, structural comparison of Table 1. The graded-null model *as formulated* (a fully reversible dose–response) cannot reproduce these trends: withdrawing the drug returns every responder toward baseline, yielding neither a durable subgroup nor a plateau. The present model, with its bistable fold, reproduces both, because a responder whose weight has crossed the untreated saddle remains in the health basin off drug. On the trends these trials reveal, the graded-null we built fails and the bistable-fold model captures them.

**Table 1:** Discontinuation trends: the graded-null model (as formulated) versus the bistable-fold model. Rows 1–2 are what the relapse-prevention studies demonstrate (Fineberg et al. ^26^; Kishi et al. ^27^); row 3 is a model prediction not yet tested. Absolute durable fractions are *not* compared (S7); only the qualitative trend is.

| Trend | Graded-null (as formulated) | Bistable-fold model |
| --- | --- | --- |
| Durable off-drug remission subgroup (about half of responders stay well after withdrawal) | Absent: fully reversible, so every responder relapses | Present: fold-crossers remain in the health basin off drug |
| Relapse-free (survival) curve plateaus rather than decaying to zero | Absent: decays toward zero | Present: near-saddle crossers relapse slowly, deeper crossers not at all |
| Durable fraction grows with treatment duration before withdrawal (*model prediction*) | Zero at any duration | Grows: longer treatment carries more patients across the saddle |

The second is *critical slowing*. Near the fold the return rate to equilibrium falls toward zero, so autocorrelation and variance in a densely sampled symptom signal rise before a shift, absent in the graded model, which relaxes at a fixed rate (Fig. 6D). It is the same slow-near-the-fold dynamics that shapes the drug-free onset transient (a long prodrome, then abrupt escalation; Fig. 3c), and it has been measured before mood transitions^8,10^ and, closest to our case, in daily assessments during cognitive-behavioral OCD treatment, where discontinuous improvement was preceded by rising instability^28,29^. These four signatures (discontinuity, bimodality, irreversibility, and critical slowing) separate the fold from a graded account; three have concordant support in existing data, and the bimodal-endpoint test is prospective (S6).

### 3.5 What population data can and cannot say

At the population level the model reproduces the meta-analytic SSRI responder band ^30^ but also predicts a responder fraction that *falls with baseline severity* (S10; Table S4). The strongest test of that dependence is the largest individual-patient-data meta-analysis to date^1^ (eleven trials, *n* = 2372), which finds the SSRI-versus-placebo benefit *nearly independent of baseline severity* (S11). Because that analysis pools unnamed SSRIs across various doses, only the severity *slope*, not the absolute level, is a well-posed target. On it, the model predicts a drug-benefit gradient about 1.6× steeper than the upper bound of Cohen’s interval (a ∼ 4.6-point mild-to-severe spread against the ≲ 2.9 Cohen permits). The model matches the severe end and over-predicts the *mild* -patient benefit, not a claim that severe patients fail to respond.

That gradient, however, is not a signature of the fold: every model we analyzed, several baseline-referenced BCM variants and the fold-free graded-null model alike, produces it (S12), because it originates in how presenting severity enters the plasticity source, which they share. It is therefore a class-wide limitation of this model class, not evidence bearing on bistability. And because the graded-null model reproduces the trial means as well (§3.4), population data (the means and the severity slope alike) cannot distinguish a fold from a graded slide. That discrimination rests on the individual-level signatures of §3.4, not on population means. The degeneracy is general: wherever opposite mechanisms fit the same trial average, discrimination must move to individual-level dynamics (critical slowing, endpoint bimodality) in OCD and beyond.

## 4 Discussion

### 4.1 From discrimination to treatment: implications of the fold

Taking the individual-level signatures of §3.4 (not the trial means) as the model’s empirical grounding, we turn to what the fold implies for treatment.

#### Clinical implications, and non-implications

The severity-gated-remission prediction is easily misread, so we state its limits. It does *not* license withholding or under-dosing SSRIs in severe OCD: severe patients still improve substantially, and the framework supports *earlier augmentation*, not less serotonergic treatment. The subclinical commitment threshold is a theoretical vulnerability construct, not a screening protocol.

The calibration attributes the SSRI plateau not to an unresponsive circuit but to a *toxicity-bounded ceiling* on how far transporter blockade can raise serotonin. The *e*_*C*_ needed to fold the attractor rises steeply with severity, so remitting severe OCD (*Y*_model,0_ ≳ 28) by serotonin alone would demand supraphysiological, serotonin-syndrome-range elevations (an extrapolation beyond the calibrated *e*_*C*_ range, under a region/receptor-uniform toxicity mapping; Table 2). Raising *e*_*C*_ further, whether by higher dose, uptake-2/OCT3 blockade, or MAO inhibition, buys diminishing benefit at rising risk (OCT3-targeting augmentation is developed in S13). Because the disease state is set by plasticity rather than serotonin, effective augmentation should act on the plasticity balance instead. That locus is the gain *α* and the threshold *θ*_*M*_, where much of current practice already works: the dopaminergic, receptor-level serotonergic, and glutamatergic agents below enter the corticostriatal balance by routes other than raising *e*_*C*_. The gain *α* is the biological seat of predisposition, a plasticity susceptibility shaped by genetics (SAPAP3, SLC1A1; ^31^). *(We emphasize that α is a calibrated effective gain identified from baseline severity; the etiological reading is a framing the model motivates, not a proven mechanism, and we neither derive α from specific exposures nor exclude b or θ*_*M*_ *as contributors*.)

**Table 2:** Three routes to the fold. What each augmenter must achieve to annihilate the OCD attractor, by presenting severity. Serotonin (sink) enters the serotonin-syndrome range for *Y*_model,0_ ≳ 27; memantine (source, via the LTP gain *α*) and antipsychotics (source, via lowering caudate *D*_1_ firing 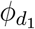 through the circuit) stay bounded, but each route has a severity ceiling beyond which it cannot fold at all (serotonin ≈ 24, antipsychotic ≈ 30, memantine ≈ 34). All entries are illustrative model quantities: the serotonin *e*_*C*_-multiples extrapolate beyond the ≲ 5.8× calibration data under a region/receptor-uniform assumption, and the memantine/antipsychotic block-to-gain step is uncalibrated (SI). Antipsychotics are taken up in §4.3.

| presenting<br>$Y_{\text{model},0}$ | serotonin (sink)<br>$e_C$ to fold | memantine (source)<br>LTP-gain reduction | antipsychotic (source)<br>caudate-$D_1$ reduction |
| --- | --- | --- | --- |
| 24 | $6.4\times$ baseline | 29% | 15% |
| 28 | $12.3\times$ (toxicity) | 46% | 25% |
| 30 | $17.3\times$ (toxicity) | 55% | 32% |
| 32 | $25.1\times$ (toxicity) | 64% | n/a |

### 4.2 Memantine: augmenting through the plasticity source

NMDA receptors set the *magnitude* of corticostriatal potentiation ^32^, so memantine (which preferentially blocks excessive NMDA activity while sparing normal transmission ^33^) lowers the LTP gain *α* and shifts striatal plasticity toward depression (experimentally via D2/endocannabinoid signalling) ^34^. Where the SSRI folds the attractor by steepening the *sink* (∝ *e*_*C*_), memantine folds it by shrinking the *source* (*α*), a route independent of *e*_*C*_ and so not bounded by the serotonin ceiling: to fold a severe (*Y*_model,0_ = 28–32) attractor the sink route needs *e*_*C*_ of 12–25× baseline (serotonin-syndrome range), the source route a bounded 46–64% gain reduction (Table 2). These serotonin multiples are extrapolations beyond the calibrated *e*_*C*_ range; only their *ordering*, that the sink route reaches toxicity while the source route stays bounded, is a model claim.

For a refractory patient held on an SSRI, a sufficient memantine gain reduction draws the OCD attractor and its saddle together until they annihilate (Fig. 7, dotted curve), leaving health the sole stable state. The mechanism yields a falsifiable prediction: memantine should help precisely where the SSRI does not fold the attractor and be null where it does, so benefit tracks SSRI non-folding (a low fractional 5-HT_1*B*_ modulation *b*), not baseline severity. The clinical-trial evidence on memantine augmentation is thin and contradictory (the largest controlled trial^35^ (*n* = 99) was null), so we present memantine as a mechanism and a qualitative prediction. The trial reconciliation under this folding-based reading, a pharmacokinetic dose-to-engagement map, and its one uncalibrated step are developed in S14. Because it acts on the gain *α*, the model also predicts a slow onset and, in principle, a prophylactic role.

**Figure 7.**
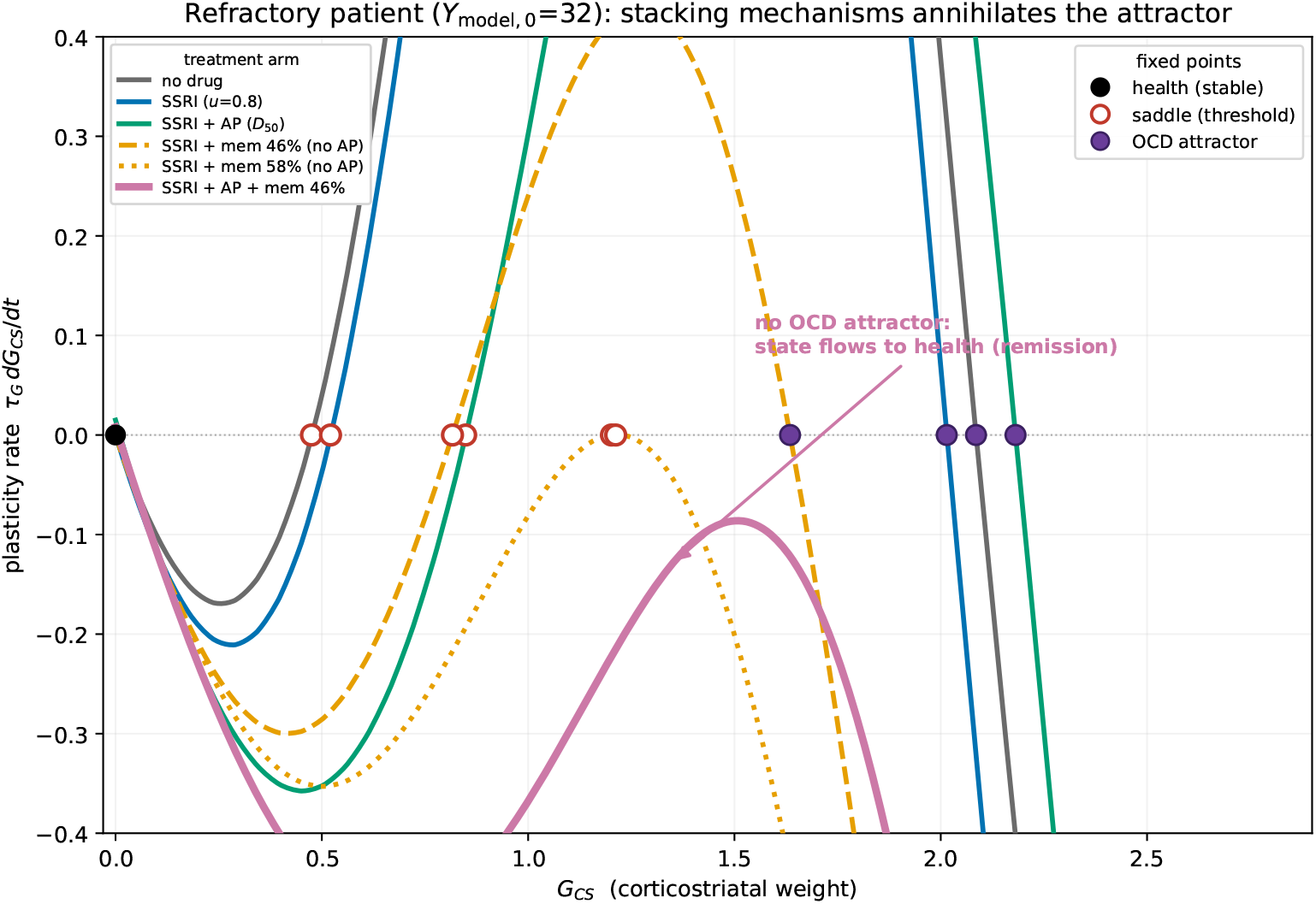
Complementary routes to the fold in a refractory patient (*Y*_model,0_ = 32). Plasticity rate *dG*_*CS*_/*dt* vs. *G*_CS_; a filled circle on the zero line marks a diseased attractor and the black circle at *G*_CS_ = 0 the healthy state. Stacking cumulatively lowers the curve but keeps the attractor: no drug (gray), +SSRI at *u* = 0.8 (blue), +AP at *D*_50_ (the SSRI+AP curve, green). Adding memantine (46% LTP-gain reduction) then removes the attractor (SSRI+AP+memantine, pink), leaving health the sole stable state. Holding the SSRI at *u* = 0.8 without the antipsychotic, SSRI+memantine at 46% (orange dashed) still retains the attractor, while raising memantine to 58% (orange dotted) folds the pair alone; each of the three contributes. Single mechanisms applied alone are shown in the Supplement.

The SSRI and memantine share a target, the plasticity balance that *sets* the fold. The remaining strategies act instead on the *circuit* that expresses it: antipsychotics at the striatal *D*_2_/indirect-pathway node, ondansetron at 5-HT_3_ receptors.

### 4.3 Folding through the circuit

Antipsychotics act one step removed from the plasticity. *D*_2_ blockade at the striatal *D*_2_-MSN population disinhibits the indirect pathway and, through the closed cortico-striato-thalamo-cortical loop, lowers caudate *D*_1_ firing 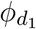, the argument of the plasticity source. Shrinking the source folds the attractor by the same destination as memantine but via the *circuit variable* 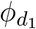 rather than the *gain α*; and because 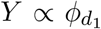, it also carries an immediate partial effect on top of the slow fold. Folding a moderate-to-severe attractor needs a 15–32% reduction of 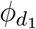, attainable by *D*_2_ blockade up to *Y*_model,0_ ≈ 30 (Table 2); in Figure 7, adding this *D*_2_ blockade to the SSRI lowers the plasticity-rate curve toward the fold, though for the refractory patient shown (*Y*_model,0_ = 32) it does not annihilate the attractor alone (the SSRI+AP curve). The picture is consistent with the augmentation meta-analysis ^3^ (32% vs. 11% response on a stable SRI). We work here in receptor coverages (% *D*_2_ blockade); the dose-to-occupancy pharmacokinetics (risperidone, aripiprazole) and the Bloch comparison are in S15. The extrapyramidal and metabolic burden of the antipsychotics motivates a receptor-level alternative.

A receptor-level alternative is *ondansetron*: 5-HT_3_ antagonism lowers excitatory and dopaminergic drive into the loop and hence the caudate rate 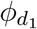, reaching the fold by the same circuit route as the antipsychotic but without *D*_2_ blockade or its burden, with adjunctive trials showing a Y-BOCS reduction ^36^ and worsening on discontinuation ^37^. The framework accommodates any circuit-level intervention that shrinks the source by lowering 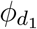 (S16).

### 4.4 Complementary mechanisms and combination therapy

A unifying consequence is that these strategies reach the *same* fold by acting on *different terms* of the plasticity equation: the SSRI steepens the sink (*be*_*C*_), memantine shrinks the source gain (*α*), and the antipsychotic and ondansetron lower its circuit argument (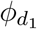). If the levers are separable and additive their effects stack, so interventions each individually sub-threshold can jointly annihilate an attractor none folds alone (Fig. 7; the three-drug combination is quantified in S17): for a refractory patient (*Y*_model,0_ = 32) an SSRI, an antipsychotic, or a 46% memantine gain reduction each leaves the attractor intact alone, yet stacked they remove it. This is the mechanistic rationale for the combination augmentation already standard in treatment-resistant OCD^3,4^, presented as a proof of principle, not a dosing rule: the model omits pharmacokinetic interactions, shared side-effects, and adherence, and drug and dose selection remains a clinical judgment outside our scope.

Finally, the same picture indicates where non-pharmacological treatment fits. The gain *α* is not intrinsic: the model reads it from presenting severity but does not generate its accrual (§3.1), which reflects an external drive of experience and learning. Behavioral therapy (ERP/CBT) plausibly acts on *that* drive rather than on the serotonergic sink or the receptor levers above, so pharmacotherapy and psychotherapy would attack different terms of the same plasticity equation. We flag this only as a conceptual mapping, untested here.

The same landscape suggests two further extensions, both developed in S16.2. First, non-pharmacological therapies (rTMS and exposure/response prevention) enter the circuit off the serotonin axis and, acting through the shared orbitofrontal node, should combine super-additively. Second, in comorbid depression and anxiety, coupling the circuit to mood loops would let one calibrated engine drive two bistable circuits at once. A pharmacological corollary is also left to future work: the serotonergic sink could be driven directly at its 5-HT_1*B*_ receptor rather than by raising serotonin (a route psilocybin recruits at cortical terminals^38^), sidestepping the toxicity ceiling that bounds serotonergic elevation.

### 4.5 Scope and limitations

Three features bound the claims, each developed where it arises. First, the readout is a *proxy* : *Y*_model_ maps a caudate firing rate to a behavioral score, so it constrains the model’s *gradient* (how outcome moves with dose and severity, on which every net drug − placebo result rests) but not its absolute registration. The one comparison in which the absolute level enters, the population responder fraction, carries a three-to-four-point offset between *Y*_model_ and Y-BOCS that we read as a property of the proxy rather than as a fit (S10). Second, the modification threshold *θ*_*M*_ is held *constant* rather than sliding with activity; this is grounded in the stabilizing, two-threshold character of corticostriatal metaplasticity and is conservative for the fold, with the supporting calculation and its evidence grade in S5.1. Third, two constants are set by criteria rather than fitted to the trial outcomes: the readout exponent *γ* is fixed so the saddle-node coincides with the remission threshold (not tuned to the responder rate), and the per-patient gain *α* is an effective calibrated quantity. The *direction* of severity-gating is therefore in part definitional, while its steep *shape* and the remission threshold are its empirical, falsifiable content (S10). Two narrower caveats complete the list: the high-dose fluoxetine miss is a SERT-saturation ceiling, not a potency error (§3.2); and the combination result is a proof of principle that omits pharmacokinetic interactions, shared side-effects, and adherence (§4.4).

## 5 Methods

### 5.1 Model specification

#### Circuit

The cortico–basal-ganglia–thalamic circuit is the nine-population mean-field model of van Albada & Robinson ^14^, in the neural-mass tradition of Wilson & Cowan^39,40^ (cortical *e*/*i*, striatal *D*_1_/*D*_2_ MSNs, GPi/SNr *p*_1_, GPe *p*_2_, subthalamic *ς*, thalamic relay *s* and reticular *r*). Each population has mean firing rate *ϕ*_*a*_ = *S*_*a*_(*V*_*a*_),

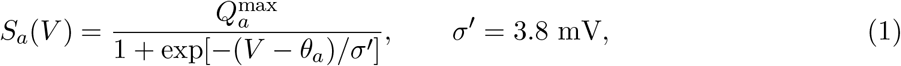

and a delay-and-filter mean-field potential 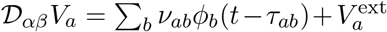 with 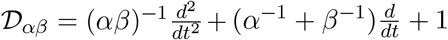. On the weeks-long treatment timescale the fast circuit is at quasi-steady state (D_*α*β_ → 1), giving nine net inputs

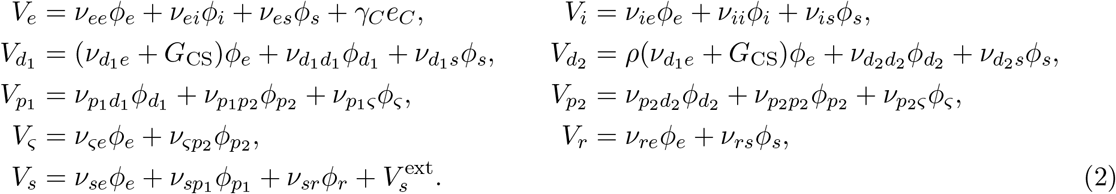

All 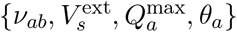 are the van Albada–Robinson resting values (Table S1; S1.1). Two terms are added to that resting network: the excess corticostriatal weight *G*_CS_ ≥ 0 and a direct cortical serotonergic drive *γ*_*C*_*e*_*C*_ (calibration gives *γ*_*C*_ → 0, S18, a data-identifiability verdict for this SSRI fit, not a claim that fast cortical serotonergic action is absent, since a fast circuit channel is exactly how ondansetron acts below), with 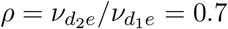.

#### Serotonin engine

Extracellular serotonin *e*_*C*_ is quasi-static, balancing autoreceptor-gated release against SERT and uptake-2/OCT3 clearance,

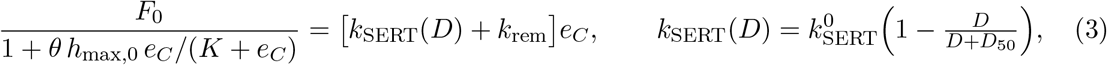

with *K* = 10 nM (5-HT_1*B*_ affinity; ^41^), 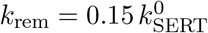 the SSRI-insensitive removal floor, *D*_50_ the [^11^C]DASB half-occupancy dose ^42^, and *F*_0_ pinned so the drug-free balance (*D* = 0, *θ* = 1) returns 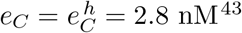. The 5-HT_1*A*_ autoreceptor gates release, not clearance, and desensitizes overweeks,

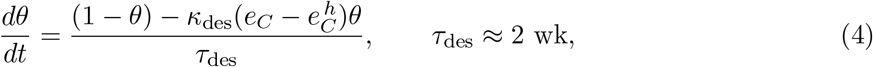

so acute reuptake blockade alone produces little net rise; the delayed lift of release suppression is part of the antidepressant onset (S1.3).

#### Corticostriatal plasticity

The weight *G*_CS_ remodels slowly under a baseline-referenced Bienenstock– Cooper–Munro rule: a bounded potentiation source opposed by a serotonin-modulated depression and a constitutive decay,

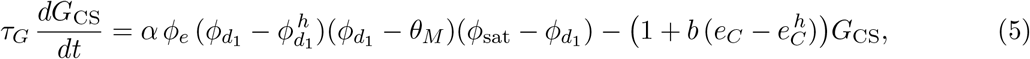

with *τ*_*G*_ the intrinsic remodeling time and *α* (per patient) the LTP gain. Because the source is a quartic product of firing rates (each in s^−1^) while *G*_CS_ is dimensionless, *α* carries units of s^4^, rendering the source term dimensionless. The constitutive decay is normalized to unit coefficient, so the drug enters as a fractional modification 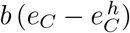 (*b* = 0.0268 nM^−1^, so the modification 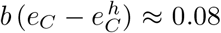 at clinical *e*_*C*_). The source’s first zero, at the healthy rate 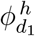, makes health (*G*_CS_ = 0) a fixed point of the remodeling; its interior zero, the modification threshold *θ*_*M*_, is the LTP/LTD crossover, set so the untreated remission/progression divide falls at the Y-BOCS subclinical boundary (S1.4). Serotonin drives 5-HT_1*B*_ depression of the synapse ^44^, so a drug-induced rise in *e*_*C*_ lowers *G*_CS_, the caudate rate, and the score.

#### Readout

Symptoms are read from the caudate *D*_1_ rate through a power law anchored at both ends by circuit quantities,

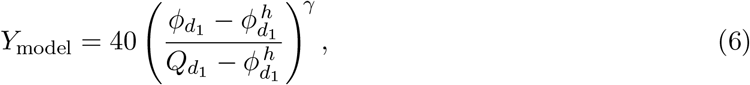

with 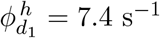 (healthy) and 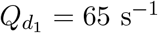 (maximal *D*_1_ rate) not fitted and *γ* = 2 (S9); the four monotone circuit rates are mutually monotone, so the choice of 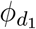 is not load-bearing (S3).

#### Expectation (placebo) response

The non-specific response enters only at the readout, *Y*_obs_(*t*) = *Y*_model_(*G*_CS_(*t*)) − *ε*(*t*) with 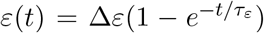 and *ε*(0) = 0^45^; it does not modify *G*_CS_, so it cancels in the net (drug − placebo) quantity used for calibration, and hypothetical interventions with unknown Δ*ε* are reported as net improvements (S19).

### 5.2 Data sources and digitization

The model was calibrated against the *net* (drug−placebo) Y-BOCS response of 13 fixed- or titrated-dose arms drawn from seven double-blind, placebo-controlled OCD trials, spanning six selective serotonin reuptake inhibitors: fluoxetine (20/40/60 mg; ^25^), citalopram (20/40/60 mg; ^46^), escitalopram (10/20 mg) and paroxetine (40 mg; ^47^), paroxetine (37.5 mg; ^48^), sertraline (pooled 50–200 mg; ^23^), paroxetine (20 → 40 mg titrated, up to 50; ^49^), and fluvoxamine (200 mg; ^50^). Two clomipramine arms ^48,51^ were held out entirely and used only as an out-of-sample falsification test.

For each arm we extracted, from the source figures and tables, the mean Y-BOCS trajectory (as absolute score, change from baseline, or already net of placebo, as reported), the assessment weeks, the baseline mean and standard deviation, and the per-timepoint standard error; drug-arm data reported as absolute or change scores were converted to the net (drug−placebo) quantity using the trial’s own placebo arm. Trajectory values read from figures were cross-checked against the reported endpoint statistics. We estimate the figure read error at roughly ±0.2–0.3 Y-BOCS points (a fraction of the plotted symbol size), small beside the reported per-point standard errors (∼ 0.4–1.5 points) but not separately propagated; because it is comparable to the 0.83-point fit RMS, that residual should be read as approaching the digitization-plus-reporting resolution floor rather than as a tighter claim, and the leave-one-arm-out cross-validation (S4) bounds its influence on the fitted coupling. Each digitized series was extracted from the corresponding trial report tabulated in S22. Per-drug half-occupancy doses *D*_50_ and the maximal SERT occupancy were taken from the [^11^C]DASB positron-emission occupancy study of Meyer et al. ^42^ for fluoxetine, citalopram, paroxetine, and sertraline; escitalopram’s *D*_50_ is set at half the citalopram value ^52,53^ (escitalopram 10 mg ≡ citalopram 20 mg), and fluvoxamine’s (*D*_50_ = 50 mg) is set from the single-dose SERT-occupancy time course of Takano et al. ^54^ (73% peak occupancy at 5 h, decaying to ∼ 50% by 24 h), matched to the sustained daily-averaged occupancy that *u* represents rather than the transient peak (order-of-magnitude potency). The healthy extracellular-serotonin reference 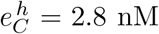 is the striatal zero-net-flux value of Mathews et al. ^43^; the remaining fixed constants and their sources are listed in Table S1.

### 5.3 Numerical implementation

All computation was performed in Python (NumPy/SciPy). The nine-population van Albada– Robinson mean-field circuit fixed point was obtained with a Powell hybrid root-finder (fsolve, tolerance 10^−10^) from multiple initial conditions to select the physiological branch; the resulting caudate 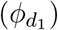 and cortical (*ϕ*_*e*_) rates were tabulated on a grid in the corticostriatal overdrive *G*_CS_ and evaluated during integration by bicubic spline interpolation. The fast extracellular-serotonin balance was treated as quasi-static: at each timestep the steady-state *e*_*C*_ was found by Brent’s method (`brentq`, tolerance 10^−7^). The slow corticostriatal-plasticity ODE (Eq. 5) was integrated by forward Euler with a step of 0.15 weeks over the trial horizon; the autoreceptor-desensitization variable was advanced on the same grid.

A trial’s predicted net trajectory is the baseline-weighted population mean: the untreated attractor is placed for each of a 15-node deterministic quadrature of the trial’s baseline Y-BOCS distribution (a Normal with the reported mean and standard deviation, truncated to ±2.5 standard deviations within [13, 35]), each node is integrated under the arm’s dose, and the nodes are averaged with their Gaussian weights. Linear stability of the fixed points, and the separation of the OCD (non-oscillatory) from the Parkinsonian (damped-resonance) regime, were established from the rightmost root of the delay-resolved characteristic (dispersion) equation, validated against the published van Albada et al. ^55^ oscillation thresholds (S20).

### 5.4 Parameter estimation

*Degrees of freedom*. The model’s adjustable quantities fall into six classes. (1) *Shared free parameters fit to the drug response*: the 5-HT_1*B*_ drug coupling *b* and the autoreceptor-desensitization coupling *κ*_des_, two constants, common to all thirteen arms and six drugs. (2) *Set by criteria, not fit to the drug data*: the readout exponent *γ* = 2 (well-posedness/parsimony) and the modification threshold *θ*_*M*_ (placed so the untreated remission/progression divide falls at the subclinical/clinical Y-BOCS boundary). (3) *Per-trial nuisance parameters*: each arm’s expectation (placebo) response carries a fitted asymptote Δ*ε* (and shared onset *τ*_*ε*_), which enters only the placebo arm and cancels identically in the net (drug−placebo) quantity used for calibration, so it does not add drug-response freedom. (4) *Fixed from independent data or the circuit* (not adjusted to the trajectories, and free to contradict them): the full van Albada–Robinson circuit weights, the readout anchors 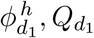, and the serotonin-engine constants (β_SERT_, six *D*_50_, 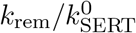, *K, τ*_des_, 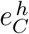, *h*_max_, *τ*_*G*_; Table S1). (5) *Determined* : the per-patient LTP gain *α*, back-solved so the untreated attractor sits at that patient’s baseline Y-BOCS. (6) *Assumed* : each trial’s baseline-severity distribution is taken as a truncated Normal with the reported mean and standard deviation. The drug-response fit therefore rests on the two shared constants of class 1; classes 2–3 add no drug-specific freedom, and classes 4–6 are inputs rather than fitted outputs.

The objective is the arm-weighted sum of squared net-trajectory residuals,

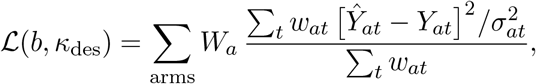

where *Ŷ*_*at*_ and *Y*_*at*_ are the modeled and observed net responses and *σ*_*at*_ the reported per-timepoint standard error. Timepoints enter inverse-variance weighted through the 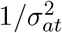 term and are additionally scaled by the arm’s enrollment, *W*_*a*_ = *N*_*a*_/⟨*N* ⟩ (the “*N* -weighting”), so larger and more precisely measured arms dominate; the per-timepoint weight is *w*_*at*_ = 1 except that the earliest, pre-drug-separation timepoint is set to zero (*w*_*at*_ = 0) in the titrated arms, where the net (drug placebo) response is near zero by construction. The minimum was located by differential evolution (population 16, tolerance 10^−7^) followed by Nelder–Mead polishing. Clomipramine was excluded from the fit and predicted post hoc.

### 5.5 Uncertainty quantification

Confidence intervals were obtained by profile likelihood: each coupling was fixed on a grid while the other was re-optimized (Nelder–Mead), and the interval is the range over which the pooled root-mean-square residual remains within 1.2× its minimum. Because *b* and *κ*_des_ both scale the drug-induced rise in extracellular serotonin, they are correlated and their marginal intervals are correspondingly broad. An identifiability analysis (S4) shows that the trajectory data determine the drug coupling *b* and the autoreceptor coupling *κ*_des_; the remodeling time constant *τ*_*G*_ is fixed on clinical grounds (of order the trial duration; §2.1), so downstream predictions (the plateau net response, the drug-attributable responder fraction) are stable across the sloppy parameter directions even where the raw constants are not.

## Data availability

The Y-BOCS trajectory data analyzed here were digitized from the published figures and tables of the cited clinical trials ^23,25,46–50^. The digitized per-arm values, weeks, and standard errors are provided in Supplementary Information, Section S22 (Digitized trajectory data), with source provenance (table versus figure-estimated) recorded in the table footnote. Every load-bearing endpoint was cross-checked against the reported values by independent re-extraction from the source-trial PDFs: table-sourced values match the reported numbers, and figure-digitized values fall within the digitization-plus-reporting resolution floor (order 0.2–0.5 Y-BOCS points) that is already propagated in the fits. The full digitized dataset is included in the public code repository (see Code availability); the detailed re-extraction cross-check is available on request.

## Supporting information

One-page summary of the work

## Data Availability

This study used data produced in prior clinical trials. These results have already been published by the researchers who conducted the clinical trials and did metadata analyses. We used published results to analyze a model. All data used are summarized in the Supplemental Information.

https://github.com/sundar1955/ocd-ssri-bistable-fold

## Code availability

All model and analysis code (Python) is publicly available at https://github.com/sundar1955/ocd-ssri-bistable-fold (MIT-licensed), and will be archived under a persistent DOI (Zenodo) on acceptance. The deposit includes the circuit, serotonin-engine and plasticity modules; the calibration, profile-likelihood, cross-validation and prediction-interval scripts; the graded-model and discriminating-signature scripts (including the detectability analysis); the fixed-parameter table; the digitized trajectory dataset; and the scripts and random seeds that regenerate every figure.

## Author contributions

S.S. is the sole author and is responsible for the design of the project scope, model selection, interpretation of the results, selection of the figures and tables, and the multiple revisions of the manuscript.

## Use of large language models

During the preparation of this work the author used a large language model (Anthropic Claude) as a programming and editing assistant: to help write and debug the Python simulation, calibration, and figure-generation code, and to assist with L^A^T_E_X typesetting, reference formatting, and copy-editing. The author formulated the hypotheses, specified the mathematical model and the analyses, and interpreted the results; all scientific content and conclusions are the author’s own. The author verified every code output and numerical result and takes full responsibility for the manuscript. The language model did not originate any hypothesis, data, or result: the empirical data are digitized from the cited clinical trials, and all predictions are produced by numerically simulating the mathematical model presented here.

