## Supplemental Information for "SSRI treatment of obsessive-compulsive disorder as motion across a bistable fold: A calibrated circuit–plasticity model of response, remission, and augmentation"

Sankaran Sundaresan

Princeton University, Princeton, NJ 08544, USA

### S1 Model justification

#### S1.1 The corticostriatal circuit

We adopt the van Albada–Robinson mean-field basal-ganglia–thalamocortical model[1] as the circuit substrate (the same cortico-striato-thalamo-cortical pathway that Rădulescu et al.[2] modeled in the Wilson–Cowan tradition, there with excitation/inhibition balance as an abstract bifurcation control rather than, as here, a drug-coupled plasticity variable) and represent OCD as a *graded, elevated but stable* fixed point: as the corticostriatal overdrive $G_{\mathrm{CS}}$ grows, the fixed point moves smoothly to higher striatal and caudate ($D_1$-MSN) firing. That this fixed point remains stable and non-oscillatory across the clinical range of $G_{\mathrm{CS}}$ (and how this is compatible with the same circuit producing Parkinsonian oscillations under dopamine loss) is established in the stability analysis (S20).

This graded elevation reproduces the imaging phenotype of OCD (caudate and striatal hypermetabolism[3–5] and elevated caudate glutamate[6], which scale with symptom severity and normalize with treatment[4–6]) and it is what makes the caudate rate $\phi_{d_1}(G_{\mathrm{CS}})$ the model's natural severity coordinate (S1.2). Across the clinical range the model's single, lumped cortical population (which includes the frontal cortex implicated in OCD imaging[5]) also rises, but far less than the caudate (Fig. S1); the cortical hyperactivity is thus a *downstream loop consequence* of the striatal lesion, not its driver. This ordering (striatal lesion primary, cortical elevation secondary) is a closed-loop output of the circuit, not an assumption.

Finally, the ratio $\rho = \nu_{d_2 e}/\nu_{d_1 e} = 0.7$ by which the lesion splits between $D_1$ and $D_2$ is read directly from the van Albada–Robinson corticostriatal weights[1]: absent information on how the overdrive should partition, we let it augment $D_2$ in the same proportion as the base corticostriatal input, so that the entire $D_2$ drive collapses to $\rho\,(\nu_{d_1 e} + G_{\mathrm{CS}})\,\phi_e$ (Methods, Eq. 2).

#### S1.2 The observational readout: choice of severity coordinate

The main text adopts the caudate $D_1$-MSN rate $\phi_{d_1}$ as the coordinate that the readout law $Y_p = 40\,[(\phi_{d_1} - \phi^h_{d_1})/(Q_{d_1} - \phi^h_{d_1})]^\gamma$ maps onto the Y-BOCS.

As the disease coordinate $G_{\mathrm{CS}}$ increases, four steady-state rates move monotonically: the caudate $D_1$-MSN rate $\phi_{d_1}$ (which rises most, $7.4 \to 64.7\ \mathrm{s}^{-1}$), the $D_2$-MSN and subthalamic (STN) rates, and the external-pallidal (GPe) rate (which falls). The orbitofrontal/cortical rate rises only modestly ($\sim$+20% at moderate severity) and non-monotonically, and the output, relay, and reticular rates change little. Any of the four monotone rates is, in principle, an admissible severity coordinate, and each has independent human support in OCD: caudate activity tracks symptom severity and normalizes with treatment[4,5]; the subthalamic nucleus is an established therapeutic node, its stimulation lowering the Y-BOCS relative to sham[7]; and external-pallidal delta power scales with obsession severity[8].

These human signals are *different observables* from the model's firing rates (glucose metabolism, receptor binding, and oscillatory field power, respectively) so the defensible claim is that these nodes *co-vary* with severity, as the model requires of a coordinate, not that any one is numerically identical to a model rate. The GPe case is instructive: the model's tonic GPe *firing* falls with severity, and reduced tonic pallidal firing is classically what permits the growth of low-frequency oscillatory *power*: so the model's falling GPe rate is consistent with the *rising* GPe delta power that is reported. By contrast, striatal $D_2$ receptor *binding* is reduced in OCD but shows no correlation with Y-BOCS[9]; it is an abnormality, not a severity coordinate, and we do not use it as one.

We adopt the caudate rate for two reasons: it has the largest and, being bounded by the $D_1$-MSN sigmoid ceiling $Q_{d_1}$, a *saturating* excursion (the natural shape for mapping onto a bounded 0–40 scale) and it has direct single-unit support: striatal MSN firing is elevated in OCD-model mice while orbitofrontal firing is not[10,11], whereas the human *metabolic* picture is more equivocal and is weighed separately in S3. The choice is not load-bearing: because $\phi_{d_1}$ and the STN rate are both strictly monotone in $G_{\mathrm{CS}}$, each is a monotone function of the other, so reading symptoms out through the STN rather than the caudate is a reparametrization of the same map. We verified this directly: rebuilding the observational law on the STN rate and recalibrating, with the readout exponent fit for each coordinate, changes the aggregate trajectory fit by only $\sim 0.6\%$ and leaves the predictions unchanged (robustness in S3). The severity coordinate is thus underdetermined by design, with the caudate a convenient, best-anchored representative.

#### S1.3 The serotonin engine

*SERT is fully blockable.* The dose-dependent clearance $k_{\mathrm{SERT}}(D) = k^0_{\mathrm{SERT}}\,[1 - D/(D + D_{50})]$ takes SERT to be *fully* blockable at saturating dose. This is supported by the [$^{11}$C]DASB occupancy study of Meyer et al.[12]: the per-drug dose–occupancy Hill fits asymptote at 86–102%, and single high doses reach 92–95% in animals, so the clinically observed $\sim 80\%$ occupancy is the value at *therapeutic* dose, not an intrinsic ceiling. Setting the maximal block to unity also makes the model's $D_{50}$ coincide, by definition, with Meyer et al.'s[12] half-occupancy $\mathrm{ED}_{50}$.

*The removal floor $k_{\mathrm{rem}}$.* The parallel, SSRI-insensitive removal route (Best et al.[13]) comprises glial and capillary uptake, diffusion, and (retained explicitly) uptake-2 by the organic cation transporter OCT3 and the plasma-membrane monoamine transporter (PMAT). Its magnitude is anchored by the striatal serotonin elevation in SERT-knockout mice of Mathews et al.[14] (6.4-fold, i.e. 18 vs. 2.8 nM by zero-net-flux, implying $k_{\mathrm{rem}}/k^0_{\mathrm{SERT}} \approx 0.18$), together with the uptake-2 (OCT3/PMAT) clearance route that Baganz et al.[15] establish qualitatively; we set $k_{\mathrm{rem}} = 0.15\,k^0_{\mathrm{SERT}}$ (the pinned value is tabulated in S2). The *functional* significance of this floor (and its blockade as an SSRI-augmentation strategy) is established by work in which decynium-22 (an OCT/PMAT blocker) with a sub-effective SSRI produces antidepressant-like effects attenuated in OCT3-knockout mice[16]; uptake-2 is stress-upregulated[17] and elevated where SSRIs are less effective[18–20]. The magnitude carries two caveats: uptake-2 is low-affinity and concentration-dependent (dominant at micromolar serotonin or when SERT is compromised, minor at tonic nanomolar) so a constant $k_{\mathrm{rem}}$ is a coarse-grained effective value; and it is partly degenerate with the autoreceptor gain. No OCT3-targeting augmentation trial has been conducted in humans, so the OCT3-targeting augmentation analysis (S13) is a novel, falsifiable prediction, not a validated result.

*The autoreceptor.* The autoreceptor gates *release*, not clearance (Best et al.[13]; Artigas et al.[21]): somatodendritic and terminal serotonin autoreceptors suppress firing and release in proportion to their sensitivity $\theta$ and to ambient serotonin, so that acutely, reuptake blockade alone produces little net rise in forebrain serotonin (Artigas et al.[21]). Over weeks of treatment the autoreceptor desensitizes (Methods, Eq. 4); its timescale $\tau_{\mathrm{des}} \approx 2$ weeks and its expression through reduced 5-$\mathrm{HT}_{1A}$ function are taken from the chronic-fluoxetine electrophysiology of Le Poul et al.[22] and the review of Artigas et al.[21], and this desensitization is the biological origin of part of the antidepressant delay. The maximal suppression of release is capped physiologically ($1 + h_{\max} = 3$-fold; terminal 5-$\mathrm{HT}_{1B}$ $\sim 1.5$-fold, Hjorth & Tao[23]; somatodendritic 5-$\mathrm{HT}_{1A}$ up to $\sim 5$-fold, Casanovas et al.[24]), *not* the $\sim 600$-fold an unconstrained fit would choose.

#### S1.4 The plasticity rule

*BCM identification.* The cortex-to-MSN synapse expresses bidirectional, dopamine-gated LTP and LTD, with the direction set by an activity threshold (Calabresi et al.[25]; Spencer & Murphy[26]; Shen et al.[27], for the $D_1$-MSN). This is the Bienenstock–Cooper–Munro (BCM) rule[28], whose biophysical basis is the calcium-control hypothesis: moderate NMDA/$Ca^{2+}$ influx yields LTD, larger influx LTP[29]. We write the plasticity of $G_{\mathrm{CS}}$ as a baseline-referenced BCM source (main-text Eq. 5), bounded above by a saturation factor (striatal synapses hold their strength within a bounded range; Calabresi et al.[25]).

*Placement of the two zeros.* The first zero, at the healthy caudate rate $\phi_{d_1}^{h}$, makes plasticity vanish at the physiological set-point, so the healthy state $G_{\mathrm{CS}} = 0$ is itself a fixed point. The second, at the modification threshold $\theta_M$, is an *interior* crossover sitting above the operating point, as the BCM rule requires (the threshold tracks and exceeds mean postsynaptic activity; Bienenstock et al.[28]). We do not fix $\theta_M$ from clinical measurement: a plasticity threshold has never been estimated from patient data in any psychiatric disorder, and transcranial-magnetic-stimulation probes in fact find cortical plasticity *normal* in OCD (Suppa et al.[30]), so we make no claim of a pathologically shifted threshold; the disorder is carried entirely by the elevated weight $G_{\mathrm{CS}}$. Instead $\theta_M$ is set so that, untreated, the divide between spontaneous remission and clinical progression falls at the Yale–Brown subclinical/clinical boundary (0–7 subclinical). Because it is set this way rather than tuned to the response data, the model's clinical-responder fraction is a *consistency check* against meta-analysis rather than a fit target; though it is set jointly by the calibrated sinks and the assumed baseline population, and is only weakly sensitive to $\theta_M$ (S5).

*The drug term.* The drug acts through the sink $b\,e_C\,G_{\mathrm{CS}}$: serotonin drives long-term depression of the corticostriatal synapse via presynaptic 5-$\mathrm{HT}_{1B}$ receptors, an effect demonstrated directly (and shown to be SSRI-inducible) by Mathur et al.[31], who establish the mechanism, sign, and persistence but not a rate constant (so $b$ is a fitted gain). The same presynaptic 5-$\mathrm{HT}_{1B}$ gain-control of cortical glutamate release has since been demonstrated at a separate cortical projection (anterior cingulate cortex to claustrum), engaged by both serotonin and the psychedelic psilocybin[32]: independent evidence for the receptor-level mechanism the sink assumes. The tonic baseline term $b\,e_C^{h}\,G_{\mathrm{CS}}$ is folded into the resting balance that holds the untreated attractor in place, so it is the drug-induced *elevation* $b\,(e_C - e_C^{h})\,G_{\mathrm{CS}}$ that drives recovery.

*Slow-plasticity route: optogenetic and genetic support.* Repeatedly hyperactivating the orbitofronto-striatal projection in mice induces a *progressive*, synaptic-plasticity-based increase in striatal drive that builds over days (acute stimulation is ineffective) and produces compulsive grooming that *persists* for weeks after the drive is withdrawn (Ahmari et al.[11]), precisely the slow build-up and self-sustaining character of $G_{\mathrm{CS}}$. That the corticostriatal synapse is the causal locus is supported genetically: deletion of the postsynaptic scaffolding protein *Sapap3* produces compulsive behavior rescued by striatal re-expression (Welch et al.[33]). In both models the phenotype is reversed by a serotonin-reuptake inhibitor only after *repeated* dosing, two weeks but not one (Ahmari et al.[11]), six days but not a single dose (Welch et al.[33]), mirroring the delayed onset of the model. The two models implicate the circuit through *different* synaptic routes (enhanced excitatory drive vs. reduced feed-forward inhibition of MSNs), so $G_{\mathrm{CS}}$ is best read as the *net effective* corticostriatal drive onto the $D_1$-MSNs rather than a commitment to a specific synaptic change.

*The bistable fold is structural, not fitted.* The disease geometry (two stable attractors separated by an interior saddle) is not fitted to the treatment trajectories. It is the generic consequence of three independently-motivated features of the source: the homeostatic zero at the healthy set-point $\phi_{d_1}^{h}$, the interior metaplastic threshold $\theta_M$ (the experimentally-established LTP/LTD crossover;

Kirkwood et al.[34], Cooper & Bear[35], Yashiro & Philpot[36], Matta et al.[37]), and the firing ceiling $\phi_{\text{sat}} = Q_{d_1}$. Any smooth source carrying these three roots in the order $\phi^h_{d_1} < \theta_M < \phi_{\text{sat}}$ is bistable; the cubic of Eq. 5 is one such choice, and the bistability does not depend on its details. In particular, freezing the presynaptic $\phi_e$ prefactor to a constant leaves the bistable structure intact at every baseline and shifts the fold occupancy by under 5% (S5). What the biology fixes is the existence and ordering of the zeros and the sign of the source; what the model chooses is the cubic form and the value of $\theta_M$. The saddle-node fold is therefore a structural property of the assumed rule, not tuned to the data, but equally not a finding independent of the rule.

#### S1.5 Serotonin acts as a sink, not a source

The drug enters the plasticity equation in one place (the sink $b\,(e_C - e^h_C)\,G_{\text{CS}}$) and not in the source (the LTP gain $\alpha$). Three considerations justify this, the last of which makes the choice not merely reasonable but the only identifiable one.

*The direct striatal evidence is a sink.* The best-characterized action of serotonin on the corticostriatal synapse is presynaptic 5-HT$_{1B}$ long-term depression of glutamate release: a reduction of synaptic *weight*, SSRI-inducible, occluding dopamine/endocannabinoid LTD (Mathur et al.[31]). That is literally a sink on $G_{\text{CS}}$. A source (LTP-gain) action would instead have to rest on a different, weaker, and directionally contested mechanism extrapolated from cortical and hippocampal preparations, with no comparably direct striatal demonstration.

*Non-identifiability makes the distinction moot for our data: the decisive point.* Suppose serotonin *also* lowered the LTP gain, through a proportional term $\dot{\alpha} = F_\alpha - \alpha - b_\alpha\,\Delta e_C\,\alpha$ with $\Delta e_C \equiv e_C - e^h_C$. At drug steady state $\alpha \to \alpha_0/(1 + b_\alpha\,\Delta e_C)$ (with $\alpha_0 = F_\alpha$ the drug-free gain), so the plasticity balance becomes

$$\alpha_0\,S(G_{\text{CS}}) = (1 + b\,\Delta e_C)\,(1 + b_\alpha\,\Delta e_C)\,G_{\text{CS}}.$$

The two couplings $b$ and $b_\alpha$ enter symmetrically and multiplicatively: the endpoint data constrain only their product. A source term is therefore *not separately identifiable* from a sink term. It merely renormalizes $b$. (Distinguishing them would require resolving distinct onset timescales $\tau_\alpha \neq \tau_G$ from the noisy early trajectory, which the data do not support.) Whatever net modulation serotonin exerts on the source, our calibration absorbs it into the sink coupling; parsimony and identifiability then dictate the sink-only form.

*The coupling is best read as an effective net sensitivity.* Accordingly we interpret $b$ not as the 5-HT$_{1B}$ rate constant alone but as the *effective net serotonergic sensitivity* of the corticostriatal weight: dominated by the Mathur 5-HT$_{1B}$ LTD but absorbing any additional source- or gain-side modulation, of either sign, that shares its steady-state signature. This also settles the source-versus-sink comparison of §S12: because a serotonergic source term only rescales potency uniformly, it cannot alter which patients cross the fold and so cannot relieve the severity gradient. A fully explicit, separately identified source+sink metaplasticity (requiring source-resolving data we do not have) is left as future work.

### S2 Model parameters, values, and justification

Table S1 lists every parameter with its status; the notes that follow justify each value.

Table S1: Model parameters and their status. Fixed/grounded quantities are taken from the experimental literature or the circuit; $\gamma$ and $\theta_M$ are set by criteria (well-posedness and subclinical/clinical saddle placement, not fit). The net (drug−placebo) trajectories calibrate two constants: the drug coupling $b$ and the desensitization coupling $\kappa_{\rm des}$; the constitutive remodeling time constant $\tau_G$ is pinned on clinical grounds (of order the trial duration; §2.1 and S4). The per-patient LTP gain $\alpha$ is determined by baseline severity, not fitted.

| Symbol | Meaning | Value / status |
|---|---|---|
| *Fixed / grounded, not adjusted in fitting* | | |
| $\nu_{ab}$ | cortico–BG–thalamic weights | canonical BG architecture[1] |
| $\rho$ | $D_2/D_1$ corticostriatal drive ratio | 0.7 van Albada & Robinson[1] ($\nu_{d_2e}/\nu_{d_1e}$) |
| $\phi^h_{d_1}$ | healthy caudate ($D_1$) rate | 7.4 $s^{-1}$ (circuit output at $G_{\rm CS}$=0; van Albada & Robinson[1]) |
| $Q_{d_1}\equiv\phi_{\rm sat}$ | $D_1$ ceiling / plasticity saturation | 65 $s^{-1}$ (circuit sigmoid ceiling; van Albada & Robinson[1]) |
| $\gamma_C$ | serotonin→cortex coupling | 0 (data-driven; §5.1, S18) |
| $k^0_{\rm SERT}$ | maximal SERT clearance rate | 7 $s^{-1}$ (physiological $V_{\max}/K_m$, Best et al.[13]; scale degenerate[§]) |
| $\beta_{\rm SERT}$ | maximal SERT block | 1 (fully blockable; Meyer et al.[12] asymptote 86–102%) |
| $D_{50}$ | half-SERT-occupancy dose | FLX 2.7, CIT 3.4, PAR 5.0, SRT 9.1 mg (Meyer et al.[12]); ESC 1.7 (≈CIT/2; Klein et al.[38]); FLV 50 (Takano et al.[39]) |
| $k_{\rm rem}$ | SSRI-insensitive removal (uptake-2/OCT3 + diffusion) | 1.05 $s^{-1}$ ($k_{\rm rem}/k^0_{\rm SERT}$=0.15; Best et al.[13], Mathews et al.[14], Baganz et al.[15]; Horton et al.[16]) |
| $K$ | 5-HT$_{1B}$ autoreceptor $K_D$ | 10 nM (Hagan et al.[40]) |
| $\tau_{\rm des}$ | autoreceptor desensitization time | 2 wk (Le Poul et al.[22]) |
| $e^h_C$ | healthy extracellular 5-HT | 2.8 nM (striatal ZNF; Mathews et al.[14]) |
| $h_{\max}$ | autoreceptor release-gain | 2 (1+$h_{\max}$=3× suppression, within ceiling) |
| *Set by criterion, not calibrated* | | |
| $\gamma$ | readout exponent, Eq. (6) | 2, ghost-closure / well-posedness (§2.1; S9) |
| $\theta_M$ | BCM modification threshold | 9.15 $s^{-1}$, subclinical/clinical saddle placement[†] (§2) |
| *Calibrated to net (drug−placebo) trajectories, 3 identified quantities* | | |
| $b$ | 5-HT$_{1B}$ drug coupling (fractional) | 0.0268 $nM^{-1}$ ($b(e_C-e^h_C)\lesssim$0.1 clinically) |
| $\tau_G$ | intrinsic remodeling time constant | 12 wk (pinned; SSE flat over $[8,15]$ wk) |
| $\kappa_{\rm des}$ | autoreceptor desensitization coupling | 0.11 (no literature value; weakly id., correlated w/ $b$) |
| *Per-patient, determined, not free* | | |
| $\alpha$ | corticostriatal LTP gain | set by each patient's baseline Y-BOCS |

[†] $\theta_M$ is set so the untreated saddle (the health/disease basin boundary) falls at the Y-BOCS subclinical/clinical threshold (§2); the trajectory fit is insensitive to $\theta_M$ across $[8,11]$ (S5), and the resulting drug-attributable responder fraction (∼0.2) is a *consistency check* against meta-analysis, developed in S10.

[§] The absolute clearance scale is likewise unidentifiable: scaling ($k^0_{\rm SERT}, k_{\rm rem}$) by any factor rescales the pinned release rate $F_0$ identically, leaving $e_C$ (and the fit) exactly unchanged; only the ratio $k_{\rm rem}/k^0_{\rm SERT}$ and the dose-shape ($D_{50}$, $\beta_{\rm SERT}$) are identifiable. $k^0_{\rm SERT}$ is therefore fixed at its physiological value for concreteness.

#### S2.1 Notes on the parameter values

**Circuit weights $\nu_{ab}$, the pathway ratio $\rho$, and the readout anchors ($\phi^h_{d_1}$, $Q_{d_1}$).** The connection weights $\nu_{ab}$, maximal firing rates, and sigmoid thresholds are the resting healthy parameterization of van Albada & Robinson[1] (their nominal set), adopted unchanged; we do not refit any circuit constant. The direct/indirect drive ratio $\rho = \nu_{d_2e}/\nu_{d_1e} = 0.7$ is *read* from those same weights rather than fitted. The two readout anchors are circuit *outputs*, not independent free parameters: $\phi^h_{d_1} = 7.4\ \mathrm{s}^{-1}$ is the caudate $D_1$-MSN rate the circuit produces at $G_{\mathrm{CS}} = 0$ (the untreated healthy baseline of the readout), and $Q_{d_1} = \phi_{\mathrm{sat}} = 65\ \mathrm{s}^{-1}$ is the $D_1$-MSN sigmoid ceiling of the same parameterization. No range or pinning is involved for these.

**The absolute clearance scale $k^0_{\mathrm{SERT}}$ and the removal floor $k_{\mathrm{rem}}$.** The overall clearance scale is formally *unidentifiable*: scaling ($k^0_{\mathrm{SERT}}, k_{\mathrm{rem}}$) by any common factor rescales the pinned release $F_0$ by the same factor and leaves the steady-state $e_C$ (and hence every fitted quantity) unchanged. Only the ratio $k_{\mathrm{rem}}/k^0_{\mathrm{SERT}}$ and the dose-response shape carry information. We therefore fix $k^0_{\mathrm{SERT}}$ at a physiological value rather than fit it: the terminal-clearance model of Best et al.[13] gives an effective low-concentration SERT rate $V_{\max}/K_m \approx 7.7\ \mathrm{s}^{-1}$, and we pin $k^0_{\mathrm{SERT}} = 7\ \mathrm{s}^{-1}$. The removal floor is set through its *ratio*, anchored by the striatal 5-HT elevation in the SERT-knockout of Mathews et al.[14] (a 6.4-fold rise, implying a residual non-SERT clearance of $\sim$0.18), together with the uptake-2 (OCT3/PMAT) clearance route established qualitatively by Baganz et al.[15]. *We pin* $k_{\mathrm{rem}}/k^0_{\mathrm{SERT}} = 0.15$ (modestly below the knockout-implied value), giving $k_{\mathrm{rem}} = 1.05\ \mathrm{s}^{-1}$. This floor is the substrate of the OCT3-targeting augmentation prediction, which is presented as net-only precisely because the floor's magnitude is range-bounded rather than point-known.

**Maximal SERT block $\beta_{\mathrm{SERT}} = 1$.** Meyer et al.'s[12] [$^{11}$C]DASB dose–occupancy fits asymptote at 86–102% across drugs, and single high doses reach 92–95%; the familiar "$\sim$80% at the minimum therapeutic dose" is a point on the Hill curve, not its ceiling. *From this* $\gtrsim$86% *asymptotic range we take SERT to be fully blockable,* $\beta_{\mathrm{SERT}} = 1$. This choice is also a modeling convenience: with $\beta_{\mathrm{SERT}} = 1$ the half-block dose $D_{50}$ coincides exactly with Meyer et al.'s[12] measured half-*occupancy* $\mathrm{ED}_{50}$, so no separate potency parameter is introduced. The small residual occupancy implied by an asymptote below 100% is absorbed, without loss, into the $k_{\mathrm{rem}}$ floor above.

**Half-block doses $D_{50}$.** Taken per drug from Meyer et al.[12]: fluoxetine 2.7, citalopram 3.4, paroxetine 5.0, sertraline 9.1 mg day$^{-1}$, point estimates, not ranges. Escitalopram and fluvoxamine are absent from Meyer et al.'s[12] panel. Escitalopram's $D_{50} = 1.7$ mg day$^{-1}$ is set at half the citalopram value, justified by the clinical-dose equivalence (10 mg escitalopram $\equiv$ 20 mg citalopram); Klein et al.[38] found 10 mg escitalopram reaches at least the SERT occupancy of 20 mg citalopram. Fluvoxamine's ($D_{50} = 50$ mg day$^{-1}$) is anchored to the single-dose SERT-occupancy time course of Takano et al.[39]: a single 50 mg dose gives $\sim$73% peak SERT occupancy at 5 h, decaying to $\sim$50% by 24 h and $\sim$25% by 53 h. Because $u = D/(D + D_{50})$ represents the *sustained* occupancy that drives weeks-long plasticity rather than the transient peak, $D_{50} = 50$ mg (giving $u = 50\%$ at 50 mg) targets this daily-averaged level ($\sim$half the peak); it is an order-of-magnitude potency, not a precise half-occupancy fit. Neither $D_{50}$ is a free assumption.

**Autoreceptor affinity $K$, desensitization time $\tau_{\mathrm{des}}$, and ceiling $h_{\max}$.** The autoreceptor affinity $K = 10$ nM is the 5-HT$_{1B}$ $K_D$ of Hagan et al.[40], a point value. The desensitization timescale is drawn from a range: Le Poul et al.[22] document somatodendritic 5-$HT_{1A}$ autoreceptor desensitization developing over 2–3 weeks of chronic SSRI; *we pin the low end,* $\tau_{\rm des} = 2$ *wk*, consistent with the 2–4-week clinical onset the trajectory data themselves prefer. The maximal release suppression is likewise range-bounded: measured autoreceptor ceilings run from $\sim$1.5-fold (terminal 5-$HT_{1B}$) to $\sim$5-fold (somatodendritic 5-$HT_{1A}$). *Within this* $[1.5, 5]$*-fold range we pin a* 3*-fold ceiling,* $1 + h_{\max} = 3$ *(*$h_{\max} = 2$*)*; the fit is insensitive to $h_{\max}$ across this range because the autoreceptor operates below saturation at therapeutic $e_C$.

**Healthy serotonin reference $e_C^h = 2.8$ nM.** The striatal basal extracellular 5-HT of Mathews et al.[14] by zero-net-flux microdialysis, reported as $2.8 \pm 1$ nM in wild-type animals. *We use the central value* 2.8 *nM*; it also pins $F_0$ through the baseline balance and so is not free.

**Criterion-set constants $\gamma$ and $\theta_M$.** These are *not* literature values and are not fit to trajectories. The readout exponent $\gamma = 2$ is fixed by the well-posedness / ghost-closure criterion of §2.1 (details in S9). The BCM modification threshold $\theta_M = 9.15$ s$^{-1}$ is placed by a criterion (the untreated saddle is required to fall at the subclinical/clinical Y-BOCS boundary) and the trajectory fit is insensitive to $\theta_M$ across $[8, 11]$ s$^{-1}$ (S5).

**Remodeling time constant $\tau_G$.** In the normalized plasticity $\tau_G\, dG_{CS}/dt = \alpha S - (1 + b(e_C - e_C^h))G_{\rm CS}$ the constitutive decay is normalized to unit coefficient (rate $1/\tau_G$) and the drug is a fractional modification $b(e_C - e_C^h)$. $\tau_G$ is only weakly constrained by the short trials (SSE flat over $\tau_G \in [8, 15]$ wk; S4.2), so rather than fit it we pin it within that flat window at a representative $\tau_G = 12$ wk, of order the trial duration (main text §2.1). Only $b$ and $\kappa_{\rm des}$ are fitted.

**Calibrated couplings $b$ and $\kappa_{\rm des}$.** These are the only quantities estimated from the net trajectories (main-text §2.1; procedure in Methods). The autoreceptor coupling $\kappa_{\rm des}$ has no independent literature value and is therefore, in effect, wholly calibrated. The 5-$HT_{1B}$ drug coupling $b$ is anchored by Mathur et al.[31] in *sign and persistence* (terminal 5-$HT_{1B}$-dependent corticostriatal LTD) but not in magnitude, so its size is fitted; the reported value is $b = 0.0268$ nM$^{-1}$ (at $\tau_G = 12$ wk).

**Per-patient LTP gain $\alpha$.** Not a shared constant and not a literature quantity: $\alpha$ is back-solved individually so that each patient's untreated attractor sits at that patient's baseline $Y_{\rm model}$ score (Methods). It carries the between-patient heterogeneity that the pooled circuit and serotonin constants deliberately do not.

### S3 The severity coordinate: empirical grounding and readout robustness

The main text reads symptom severity from the caudate ($D_1$-MSN) firing rate $\phi_{d_1}(G_{\mathrm{CS}})$ (its §2.1).

#### S3.1 Empirical grounding

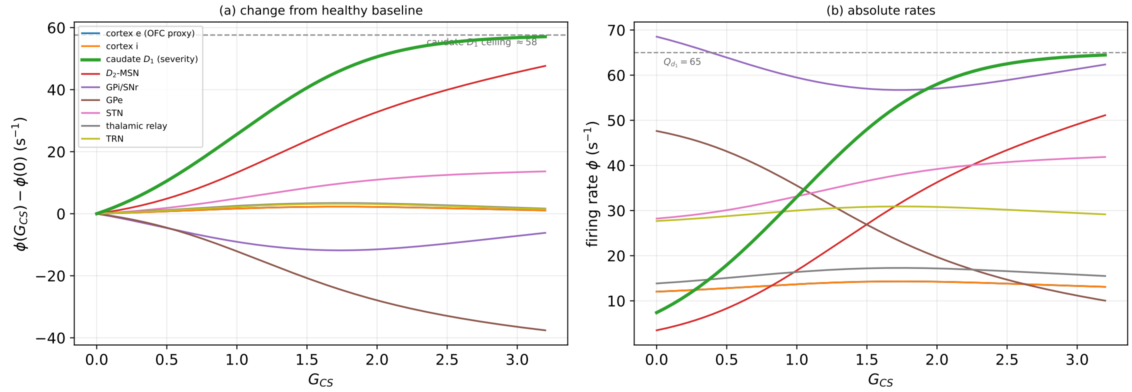

Figure S1: **Steady-state population firing rates vs. the corticostriatal excess $G_{\mathrm{CS}}$ (drug-free).** *(a)* Change in each population's rate relative to the healthy baseline. Four rates vary monotonically, caudate $D_1$ ($\phi_{d_1}$, largest excursion; $7.4 \to 64.7$ s$^{-1}$ absolute), $D_2$-MSN, subthalamic (STN), and external pallidal (GPe, decreasing), while the cortical/orbitofrontal rate rises only modestly and non-monotonically and the remaining populations barely change. Any of the four monotone rates is an admissible severity coordinate; $\phi_{d_1}$ has the largest, *saturating* excursion (bending toward the dashed ceiling $Q_{d_1} - \phi^h_{d_1}$, the natural shape for a bounded 0–40 scale) and the best empirical support. *(b)* The same rates in absolute terms: $\phi_{d_1}$ (thick) rises from 7.4 toward its sigmoid ceiling $Q_{d_1} = 65$ s$^{-1}$ (dashed). These panels underpin the readout map of main-text Fig. 2a.

Three considerations bound the choice of the caudate rate as severity coordinate. *First*, the coordinate itself is supported by single-unit electrophysiology in mouse models of OCD: in *Sapap3*-mutant mice (compulsive over-grooming) the baseline firing rate of striatal MSNs is significantly *elevated* while that of orbitofrontal pyramidal cells is unchanged (Burguière et al.[10]), and repeatedly driving the orbitofronto-striatal projection produces a progressive rise in postsynaptic striatal firing that parallels the emergence of compulsion (Ahmari et al.[11]). Here $\phi_{d_1}$ is a *firing rate*: human caudate *glucose metabolism* is reported inconsistently, elevated in absolute terms (Baxter et al.[3]), at trend level in chronic childhood-onset patients (Swedo et al.[41]), and *reduced* in a recent voxel-based analysis (Hou et al.[42]), so the proxy rests on the single-unit firing evidence and makes no claim about metabolism. *Second*, the direction of the map is validated *within patients* (Baxter et al.'s[4] percentage-change correlation); a cross-sectional, between-patient monotonicity of $\phi_{d_1}$ against severity has not been measured directly and is, at this stage, a structural assumption of the model rather than an established fact. *Third*, the model's modest, non-monotone cortical rise is itself consistent with the data: orbitofrontal *firing* is comparatively unchanged in *Sapap3* mice (Burguière et al.[10]), and human orbitofrontal hyper-metabolism, though robustly replicated (Baxter et al.[3]; Swedo et al.[41]; Hou et al.[42]), is quantitatively modest where quantified (of order +20% in

Baxter and Swedo); the model further predicts that the cortical rate turns over at supra-severe $G_{\mathrm{CS}}$ (main-text Fig. 1), so that orbitofrontal activity behaves as a predictor of state rather than a faithful tracker of it.

#### S3.2 Robustness to the choice of readout ($D_1$ vs. STN)

The model requires only a *monotone* severity coordinate; the specific rate is not load-bearing. Over the clinical range ($G_{\mathrm{CS}} \in [0, 3.45]$) both the caudate rate ($\phi_{d_1} : 7.4 \to 64.7\ \mathrm{s}^{-1}$) and the subthalamic rate ($\phi_{\mathrm{STN}} : 28.2 \to 42.1\ \mathrm{s}^{-1}$) are strictly monotone in $G_{\mathrm{CS}}$, so $\phi_{\mathrm{STN}}$ is itself a strictly monotone function of $\phi_{d_1}$ (Fig. S2). Because $\phi_{\mathrm{STN}} = g(\phi_{d_1})$ with $g$ monotone and invertible over the clinical range, any readout $Y_{\mathrm{model}} = F(\phi_{d_1})$ can be rewritten identically as $Y_{\mathrm{model}} = F\big(g^{-1}(\phi_{\mathrm{STN}})\big)$: choosing the STN rather than the caudate as the severity coordinate is a relabeling of the *same* map, not a different model. The severity coordinate is thus underdetermined by design, and the caudate is a convenient, best-anchored representative rather than a load-bearing assumption.

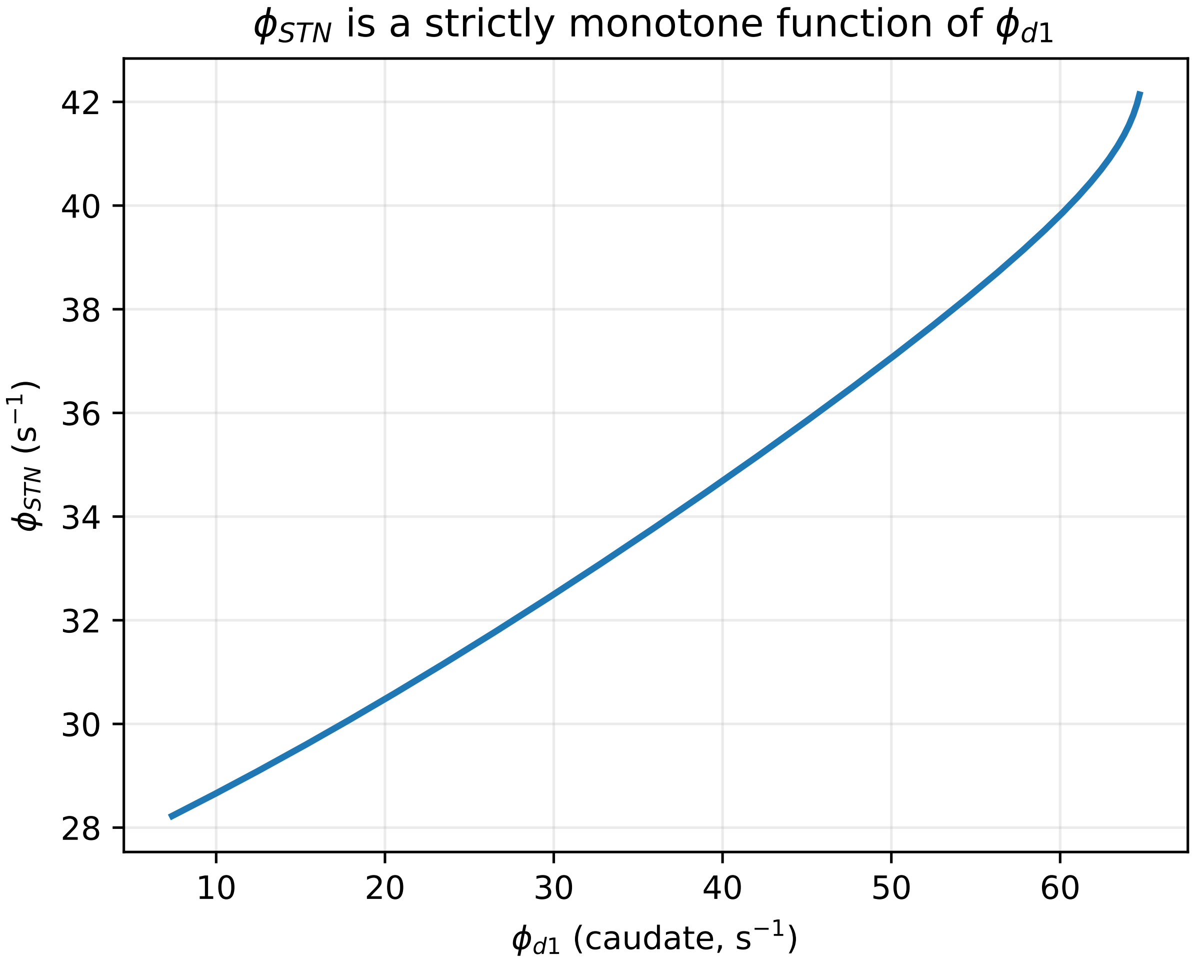

Figure S2: **The severity readout is robust to the choice of monotone coordinate.** The subthalamic rate $\phi_{\mathrm{STN}}$ is a strictly monotone (invertible) function of the caudate rate $\phi_{d_1}$ across the clinical range, so either can serve as the severity coordinate: the two are related by a reparametrization, and any readout built on one is identically a readout built on the other.

### S4 Calibration, identifiability, and validation

#### S4.1 The 13-arm calibration

The slow model is calibrated to the net (drug−placebo) Y-BOCS trajectories of 13 dose-arms across six SSRIs (fluoxetine, citalopram, escitalopram, paroxetine, sertraline, fluvoxamine; digitized sources and doses in §S22), with clomipramine held out for a falsification test (main text §5). The trajectories fix *two* constants: the 5-HT$_{1B}$ drug coupling $b$ and the autoreceptor-desensitization coupling $\kappa_{\mathrm{des}}$; every other constant is set independently, from the literature or by criterion (Table S1). At the imaging-measured per-drug SERT potencies $D_{50}$, the two shared couplings reproduce all 13 arms with a pooled RMS of 0.83 Y-BOCS points over 84 net timepoints (Fig. S3). We retain the measured $D_{50}$ throughout. One arm (sertraline, Greist et al.[43]) fits less well; rather than absorb it by floating that drug's potency, we carry it into the Discussion as a falsifiable, drug-specific prediction (an apparent under-availability of sertraline relative to its measured SERT occupancy).

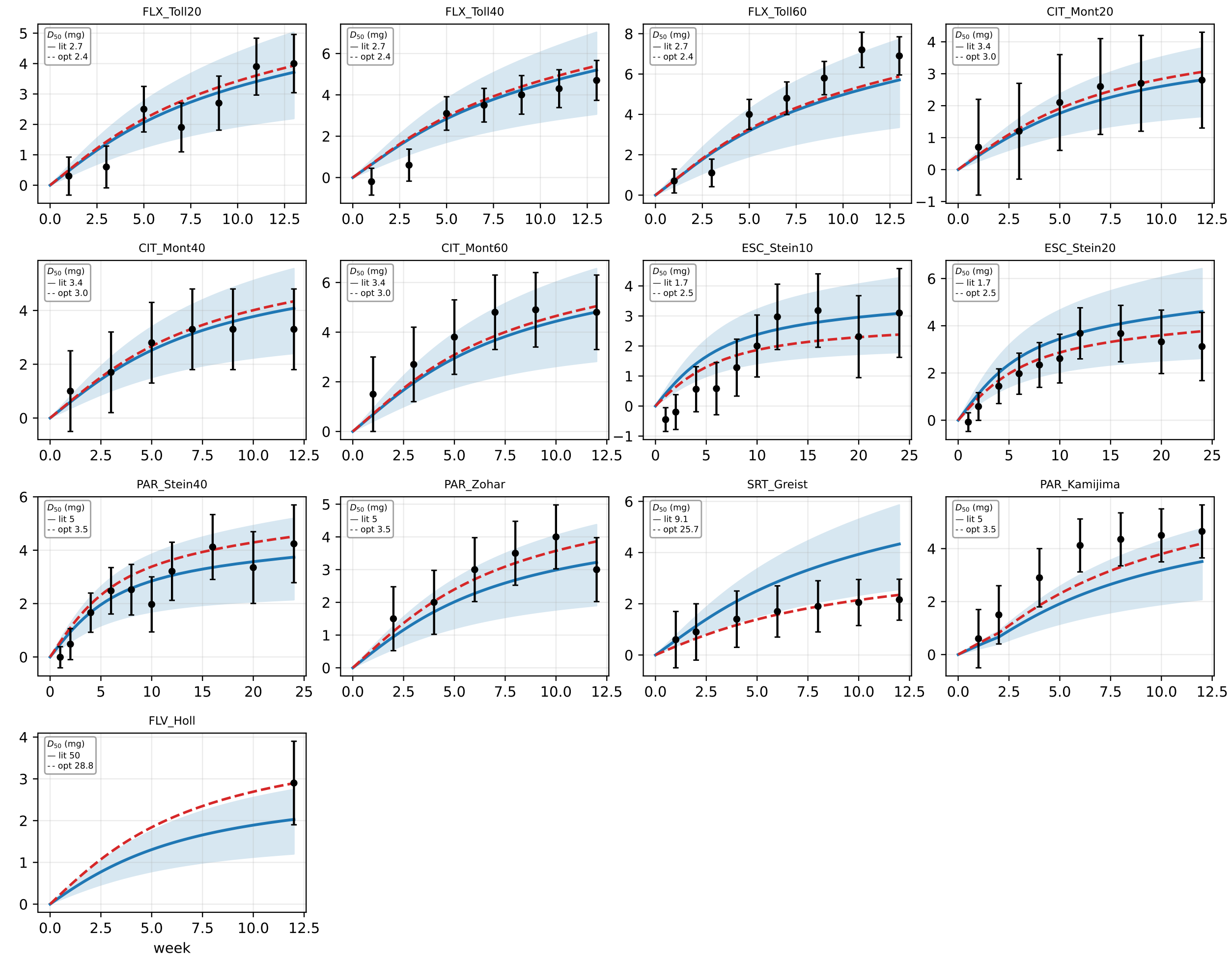

Figure S3: **Calibration: all 13 SSRI dose-arms.** Model net (drug−placebo) Y-BOCS trajectory (blue, with the $b$ confidence band) against digitized trial means ±SE (black), at the measured per-drug $D_{50}$. Two shared couplings ($b, \kappa_{\mathrm{des}}$) reproduce every arm (pooled RMS 0.83).

#### S4.2 Identifiability: two fitted constants, stiff predictions

In the normalized plasticity $\tau_G \dot{G}_{\mathrm{CS}} = \alpha S - (1 + b(e_C - e_C^h))G_{\mathrm{CS}}$, the trajectories determine $b$ and $\kappa_{\mathrm{des}}$ (the latter appearing indirectly, through the equation for $e_C$) but constrain the remodeling time $\tau_G$ only weakly (the pooled residual is flat over $\tau_G \in [8, 15]$ wk; Fig. S4b); we fix $\tau_G = 12$ wk, of order the trial duration. The remaining constants are fixed on independent grounds: the serotonin-engine parameters and the autoreceptor release-gain $h_{\max} = 2$ (a 3× suppression ceiling, within the measured 5-HT$_{1B}$/5-HT$_{1A}$ autoinhibition range; §S2), and the BCM threshold $\theta_M$ by the subclinical-saddle criterion: the value that places the untreated health/disease basin boundary at the Y-BOCS clinical threshold (§S5). The joint optimum is $b = 0.0268$ nM$^{-1}$, $\kappa_{\mathrm{des}} = 0.11$. Because $b$ and $\kappa_{\mathrm{des}}$ both scale the drug-induced serotonin rise, they are individually *sloppy* and correlated ($b \in [0.016, 0.036]$ nM$^{-1}$ within 1.2× the minimum RMS; Fig. S4a). The model's *outputs*, however, are stiff along this degenerate direction: the citalopram-40 twelve-week plateau holds at $4.5 \pm 0.15$ Y-BOCS and the pooled RMS at $\sim 0.83$ across the manifold. We therefore report the headline predictions (fold occupancies, remission fractions) as bands propagated across the $b$ interval, not as point values; the severity-gating structure ($u_{\mathrm{fold}}$ (the SERT occupancy $u$ at which the saddle-node annihilation leaves the healthy state as the only attractor) rising monotonically with baseline severity, the most severe attractors resisting folding at any tolerable occupancy) holds at every draw and is a property of the model class, not of a single fit. Leave-one-arm-out cross-validation (refit on twelve arms, predict the thirteenth) gives a mean held-out RMS of 0.81, essentially the in-sample 0.83, so no single arm drives the fit.

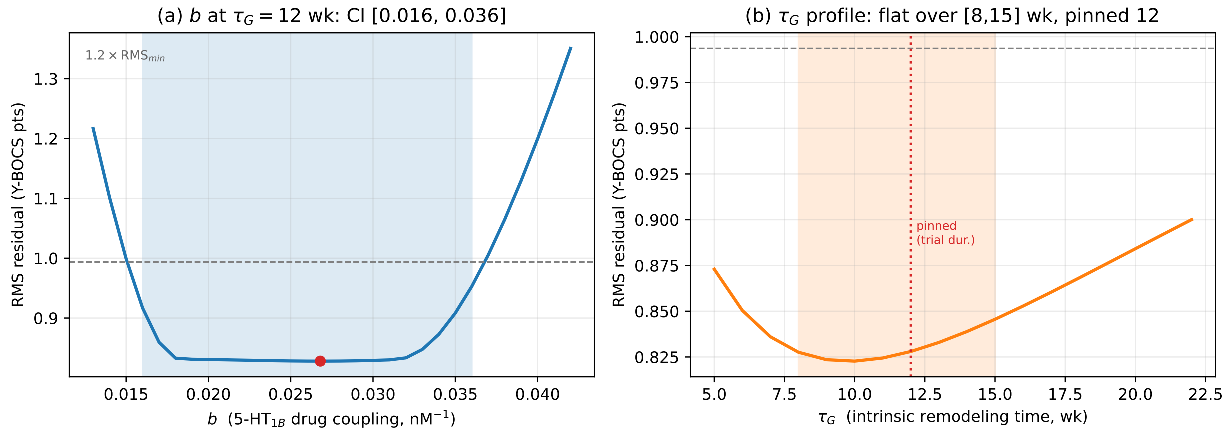

Figure S4: **Two fitted constants; $\tau_G$ pinned on clinical grounds.** (a) Marginal profile of the 5-HT$_{1B}$ coupling $b$ (with $\kappa_{\mathrm{des}}$ co-fit); the shaded band is the RMS $\leq 1.2\times$RMS$_{\min}$ interval, $b \in [0.016, 0.036]$ nM$^{-1}$. (b) The residual is flat along the $\tau_G$ profile over $\tau_G \in [8, 15]$ wk (shaded; dotted line = pinned $\tau_G = 12$ wk), so $\tau_G$ is only weakly constrained by the short trials and is fixed on clinical grounds.

#### S4.3 Validation

The *frozen* model (calibrated couplings, literature $D_{50}$, no refitting) predicts an independent multicenter sertraline trial not used in calibration, Kronig et al.[44] ($N = 85$ vs. 79, baseline Y-BOCS 25.2), at a held-out RMS of 0.58 Y-BOCS points, comparable to the in-sample 0.83 (net response 4.5 predicted vs. 4.2 observed). As a specificity test, clomipramine (a mixed serotonin–norepinephrine reuptake inhibitor, held out entirely) is *under*-predicted by the SERT-only model (endpoint 2.2 vs. observed 10, Greist et al.[45]; 1.4 vs. 3.0, Zohar & Judge[46]; a +3.4-point mean bias, far outside the 0.83 SSRI error). The model therefore correctly *fails* for a non-SERT drug: this is evidence of mechanistic specificity, since a model that fit clomipramine as well as the SSRIs would be fitting the wrong mechanism.

##### S4.4 Severity-gated response and remission

The drug-attributable improvement is gated by baseline severity (Fig. S5): mild presentations clear the 35% response threshold on drug alone while severe ones plateau below 25%, and remission (endpoint $< 12$) falls off far more steeply than response. The steep remission gradient is the model's distinctive, as-yet-untested prediction. Its relationship to the flat drug-attributable effect of Cohen et al.[47] is analysed in §S11 (main text §3.4): on the cleanest comparison (the between-arm mean difference) the model predicts a gradient in the right direction but somewhat too steep, over-predicting the benefit for *mild* rather than severe patients, a discrepancy shared with fold-free variants (§S12). Cohen's endpoint is moreover a placebo-inclusive $\geq$35% *response*, not remission; a remission-stratified re-analysis of the already-assembled individual-patient sets would test the distinctive remission prediction directly, without a new trial.

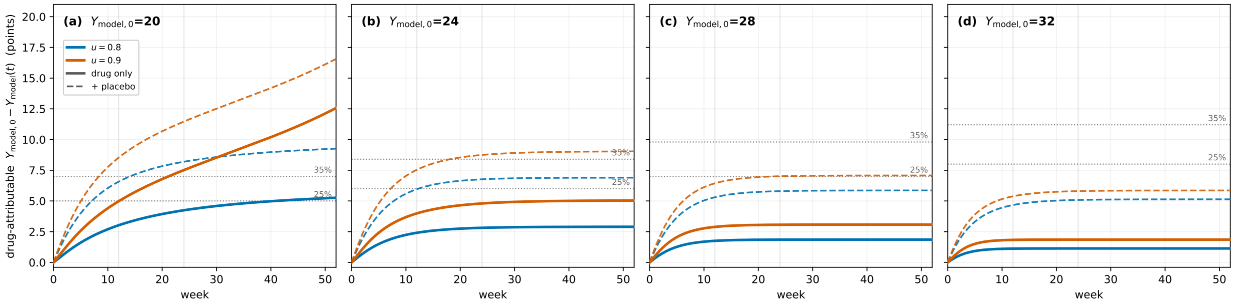

Figure S5: **Drug-attributable improvement is gated by baseline severity.** Trajectories of the drug-attributable drop $Y_{\text{model},0} - Y_{\text{model}}(t)$ for four baselines at two SERT occupancies ($u = 0.8, 0.9$); dotted lines mark 25%/35% response. Solid curves are the drug-only (net) response; dashed curves add an illustrative placebo/expectation contribution $\varepsilon(t) = \Delta\varepsilon\,(1 - e^{-t/\tau_\varepsilon})$ ($\Delta\varepsilon = 4$ Y-BOCS points, $\tau_\varepsilon = 5.5$ wk), floored so total Y-BOCS $\geq 0$; the parameter values for the placebo effect are chosen as an illustration within the range found from the data sets analyzed. Mild baselines clear 35% on drug alone; severe ones plateau below 25%. Full remission of the milder baselines is a $t \to \infty$ outcome (critical slowing near the fold), so the 24-week endpoint understates it.

### S5 The trajectory fit does not identify $\theta_M$

$\theta_M$ is fixed not by the treatment trajectories but by an independent criterion: the value at which the untreated saddle (the health/disease basin boundary) sits exactly at the Y-BOCS subclinical/clinical threshold (main text §2). This criterion determines $\theta_M$ *sharply*: it is the numerical solution of a single placement condition, which is why the adopted value carries three significant figures ($\theta_M = 9.15\ \mathrm{s}^{-1}$). The precision is inherited from the saddle-placement condition, not claimed from the trajectory fit, which is in fact nearly insensitive to $\theta_M$: profiling it (refitting the other constants at each value) leaves the pooled discrepancy flat to within 0.6% over $\theta_M \in [8, 11]$, degrading only for $\theta_M \gtrsim 14$. That insensitivity is precisely why we fix $\theta_M$ by the saddle criterion rather than fit it (a trajectory fit would be ill-posed) and it guarantees that no trajectory-based conclusion depends on the exact value.

#### S5.1 The sliding-threshold question

BCM theory's defining feature is a modification threshold that *slides* with time-averaged activity (metaplasticity), whereas we hold $\theta_M$ fixed. We do not merely assert this: we built the activity-slaved sliding threshold explicitly and compared it with the constant one on the same trajectories. It helps first to distinguish three objects the symbol "$\theta_M$" can denote, because they are not limits of one another. (A) The *constant separatrix* we adopt is a *low* threshold ($\theta_M = 9.15\ \mathrm{s}^{-1}$, just above the healthy caudate rate $\phi^h_{d_1} \approx 7.4\ \mathrm{s}^{-1}$): it is the interior boundary between the health and disease basins, and severity is carried not by it but by the per-patient gain $\alpha$. (B) A *fast* BCM slide ($\tau_\theta \ll \tau_G$) tracks running activity, so $\theta_M$ sits *high* (near the diseased rate, $\sim$30–56 $\mathrm{s}^{-1}$) and chases the weight down during treatment. (C) A *frozen* BCM slide ($\tau_\theta \gg \tau_G$) holds $\theta_M$ at the value equilibrated over the disease course, again *high* and patient-specific. The constant separatrix (A) is therefore *not* the $\tau_\theta \to \infty$ limit of the sliding family: that limit is (C), a high, patient-specific threshold, whereas (A) is a low, universal one.

Sweeping the whole family ($\tau_\theta$ from $\ll \tau_G$ (slaved) to $\gg \tau_G$ (frozen), together with the metaplastic exponent and a patient-to-patient spread) no configuration reproduces the graded dose–response *and* a flat severity profile at once. The fast end (B) fits the trajectories but makes the severity gradient *steeper* (drug response concentrated almost entirely in mild patients); the frozen end (C) flattens the barrier but turns the dose–response all-or-nothing (each patient either fails or fully remits; pooled discrepancy $\chi^2 \sim$20–760), which the graded trial means exclude. In between, the fit degrades before the profile flattens, so the well-fitting and flat regions never overlap. The activity-slaved family is dominated throughout by the constant separatrix (A): sliding buys no improvement in fit and no reduction of the severity-slope mismatch (§S12). The choice is also defensible on the substrate. First, the sliding-$\theta_M$ phenomenon is cortical/hippocampal (Kirkwood et al.[34]) and is mechanistically linked to the NR2A:NR2B NMDA-subunit switch (Yashiro & Philpot[36]), whereas corticostriatal LTD is NMDA-*independent* and runs on two separate, dopamine-gated calcium thresholds for LTP and LTD (Calabresi et al.[25]; Shen et al.[27]), not the single cortical crossover, so the cortical sliding law does not transfer to the striatum. Second, the metaplasticity in these striatal models is itself a *sliding* threshold (two of them, dopamine-gated), and what it stabilizes is individual synaptic *weights* during learning: shielding potentiated and depressed synapses from being overwritten (Trpevski[48]; Khodadadi et al.[49]). These are single-neuron learning models: they establish that the striatal substrate is metaplastic, but say nothing about whether such metaplasticity would stabilize or erode a network-level attractor, a mapping that is ours and that (S8.5) turns on the setpoint and the timescale. We therefore treat the constant $\theta_M$ as a coarse single-threshold reduction on the treatment window, not as a mechanism these results endorse. Third, a strong slide toward a *healthy* setpoint is excluded on clinical grounds: imposed directly, it annihilates the OCD attractor below $Y_{\mathrm{model},0} \approx 24$, which would mis-classify moderate OCD as non-persistent and predict spontaneous remission, contrary to its documented chronicity (an adapted, elevated setpoint would instead sustain the state; S8.5). The argument closes with two honest boundaries. The claim established here is specific (the *canonical, activity-slaved* BCM sliding threshold does not improve the fit or reduce the severity-slope mismatch) and not the stronger claim that no metaplastic rule whatever could help; a fully general, network-scale metaplasticity (a coupled $(G_{\mathrm{CS}}, \theta_M)$ or $(G_{\mathrm{CS}}, \alpha)$ system) remains legitimate future work. And the choice is not load-bearing in any case: $\theta_M$ only relocates the interior separatrix, so the model's core conclusions (bistability, the fold, severity-gated remission, and the discriminating signatures) are invariant to how it is treated, as the profiling of §S5 and the readout- and rule-sweeps of §S12 confirm. We therefore adopt the constant $\theta_M$ as a tested, non-load-bearing working choice, grounded in the stabilizing striatal picture and safe against the clinical reductio above.

### S6 The matched graded model and the discriminating signatures

The Discussion's central claim is that SSRI treatment moves a *bistable* fixed point across a fold rather than sliding a single stable state down a graded dose–response. Here we build the fairest matched graded model, show that it matches the trial means as well as the bistable model does, and identify the individual-patient-level observables on which the two diverge (main-text Fig. 6).

#### S6.1 Construction and fit

The graded model is the identical assembly (same serotonin engine, transporter and autoreceptor pharmacokinetics, readout $Y = Y_p(G_{\text{CS}})$, and per-patient baseline placement) with the one non-monotone element that creates the fold removed: the baseline-referenced BCM plasticity source, whose interior hump produces the saddle-node, is replaced by the monotone saturating *indirect-response null* $S_{\text{IR}}(G_{\text{CS}}) = G_{\text{CS}}/(1 + G_{\text{CS}}/G_m)$ (Eq. (8)), constructed and motivated as the fair fold-free comparator in §S8, where the full source-form family is analyzed. The plasticity then takes the single-branch form

$$\tau_G \dot{G_{\text{CS}}} \;=\; \alpha_g\, S_{\text{IR}}(G_{\text{CS}}) \;-\; \big(1 + b(e_C(u) - e_C^h)\big)\, G_{\text{CS}}, \tag{1}$$

in which $G_{\text{CS}}$ is the corticostriatal plasticity variable, $\tau_G$ its remodeling time, $\alpha_g$ the graded plasticity gain (fixed so the untreated attractor, $u = 0$, sits at the patient's baseline Y-BOCS), $G_m$ the saturation scale of the source, $b$ the 5-HT$_{1B}$ drug coupling, $e_C(u)$ the steady-state extracellular serotonin at SERT occupancy $u$, and $e_C^h$ its healthy baseline. Setting $\dot{G_{\text{CS}}} = 0$ leaves a *single* stable fixed point,

$$G^*_{\text{CS}}(u) \;=\; G_m\left(\frac{\alpha_g}{1 + b(e_C(u) - e_C^h)} - 1\right), \tag{2}$$

which slides smoothly toward health ($G^*_{\text{CS}} \to 0$) as occupancy $u$ rises and the drug-driven serotonin $e_C(u)$ increases, a graded slide with no fold. Here $G_0$ is the $G_{\text{CS}}$ value corresponding to the patient's initial Y-BOCS, and the relation $G_0 = G^*_{\text{CS}}(0) = G_m(\alpha_g - 1)$ is used to calibrate $\alpha_g$ (equivalently $\alpha_g = 1 + G_0/G_m$). Refit to the same 13 net SSRI arms (84 timepoints) *with its own drug coupling $b$* (fairness requires giving each mechanism its best fit rather than forcing the graded model to inherit the fold's coupling) it reaches a weighted residual of 6.50 versus 6.27 for the bistable model (plain RMS 0.90 vs 0.83 Y-BOCS points). That $\sim$4% difference is within the noise of thirteen arms and carries no evidential weight: on the population-mean trajectories the two models are statistically indistinguishable, which is precisely why the case below is made on the individual-patient-level signatures, not on goodness-of-fit to the mean. The mean is a degenerate observable that two opposite mechanisms reproduce alike.

#### S6.2 The four discriminating signatures

*(i) Discontinuous individual remission.* As steady occupancy rises, the fold patient's endpoint holds near baseline and then drops by $\sim$13 Y-BOCS points at the saddle-node ($u \approx 0.81$ for $Y_{\text{model},0} = 20$), whereas the graded patient declines smoothly (largest step 0.4). The fold occupancy climbs with severity ($Y_{\text{model},0} = 16 \to u_{\text{fold}} = 0.46$; $20 \to 0.82$; $24 \to 0.97$; $\geq 26$ never folds within the tolerable range), the structural origin of severity-gated remission.

*(ii) Durable off-drug remission.* With the drug removed after a crossing, the fold patient remains at $Y \approx 0$ because health is a persistent attractor, while the graded patient returns to baseline. A

graded, reversible model cannot produce off-drug persistence or the re-treatment path-dependence seen after relapse.

*(iii) Bimodal endpoint distribution.* For a population (baseline $Y \sim \mathcal{N}(24, 5.5)$) at a clinical dose ($u = 0.8$), evaluated at the *attractor*, the fold gives a bimodality coefficient BC$= 0.63$ (above the 0.55 threshold) and a treated-to-baseline variance ratio VR$\approx$2.0, against BC$= 0.36$, VR$= 0.95$ for the graded model; across occupancy the fold crosses BC$= 0.55$ by $u \approx 0.5$ and inflates VR to $\approx$2.2, while the graded model stays unimodal with VR$\approx$1 throughout (Fig. S6). Because of critical slowing (iv) this develops only slowly, so at a realistic trial horizon the signal is markedly weaker (§S6.3).

*(iv) Critical slowing.* The fold's relaxation rate $\lambda(u)$ falls to zero at the saddle-node, so autocorrelation and variance in a densely sampled symptom signal rise before the transition; the graded model relaxes at a bounded rate and shows no such early warning. Critical slowing signals *proximity to a fold* (it accompanies any saddle-node), so the signature that strictly requires two coexisting attractors is the durable off-drug remission of (ii).

The *magnitudes* of these signatures (the jump size, BC, VR, the occupancy at which the jump occurs) shift with the readout exponent $\gamma$ and threshold $\theta_M$ within their defensible ranges; their *direction* does not. A discontinuous jump, a bimodal endpoint with VR$> 1$, and a vanishing relaxation rate for the fold, versus smooth, unimodal, bounded behavior for the graded model, follow from bistability itself rather than from the calibrated numbers, and survive the source-form family of §S8 and the readout-exponent ($\gamma$) and threshold ($\theta_M$) variations established in §S9 and §S5.

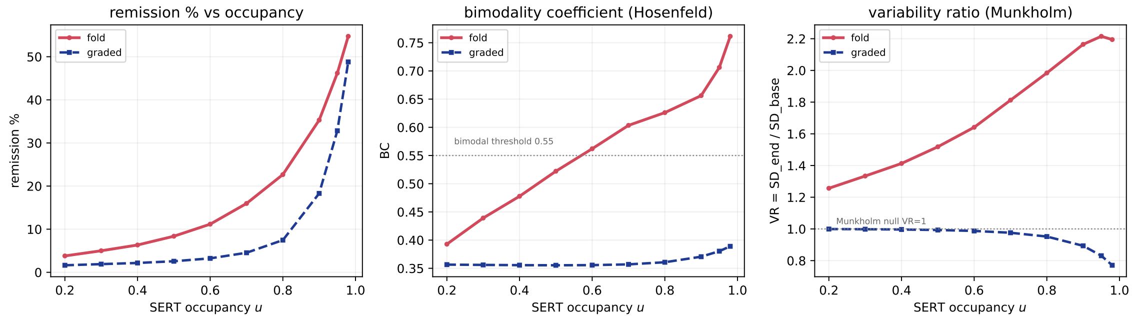

Figure S6: **Dose dependence of the endpoint-distribution signatures.** Across SERT occupancy $u$, the bistable fold (red) and the matched graded model (blue) differ in remission fraction (left), bimodality coefficient (center; dotted line, the 0.55 bimodal threshold), and variance ratio (right; dotted line, VR$= 1$). The fold crosses into bimodality by $u \approx 0.5$ and inflates variance to VR$\approx$2.2; the graded model remains unimodal with VR$\approx$1 at every dose.

#### S6.3 Trial horizon versus attractor, and a pre-registerable test

This subsection concerns the bistable-fold model alone: how its own treated-endpoint distribution (population $\mathcal{N}(24, 5.5)$ at clinical occupancy $u = 0.8$) develops over time. Signatures (i) and (iii) are evaluated at the *attractor* ($t \to \infty$), which shows *where* the two stable states lie; a finite trial shows only how far a cohort has *moved* toward them. Near the fold the approach flattens (critical slowing, iv), so at 12–24 weeks the distribution has not yet split into two modes: formal bimodality (BC$> 0.55$) is effectively an *infinite-time* property, BC staying near 0.37 through week 24 and reaching 0.63 only at the attractor. What *is* visible within a trial is a *variance inflation* that grows with duration (VR $\approx$ 1.1, 1.3, 1.9 at 12 weeks, 24 weeks, and the attractor), itself a testable prediction. This is why the main text (§4.1) rests the discriminating case on *irreversibility* and *critical slowing*: both observable at trial timescales and already concordant with OCD data (durable off-drug remission: Koran et al.[50], Hollander et al.[51], Fineberg et al.[52], Maina et al.[53]; critical slowing: Schiepek et al.[54], Heinzel et al.[55]), and treats the discontinuity and bimodality as longer-timescale predictions.

The distribution of trial-end scores nonetheless gives a concrete, pre-registerable discriminator. Here *endpoint* means each patient's Y-BOCS at the *end of the trial* (12–24 weeks), a finite-time observation across a treated cohort, not the $t \to \infty$ attractor of signature (iii). The test looks at the *spread* of those trial-end scores, not the mean. The recipe is to fit a two-component mixture (a residual-OCD mode plus a remission atom) to the treated-arm endpoints and compare the treated spread with baseline. The fold and the graded null then diverge cleanly. The fold predicts a variance ratio VR$> 1$ that *grows the longer the trial runs*, with formal bimodality (BC$> 0.55$) appearing only in the longest trials. The graded null predicts a single unimodal shift with VR$\approx 1$ at every duration. The growing variance ratio is therefore the primary thing to look for, and a longer follow-up sharpens it.

Such endpoints already exist in assembled individual-patient sets (Ramakrishnan et al.[56]; the placebo-controlled SSRI arms of Cohen et al.[47]), though data-sharing constraints place them outside this study. On model-simulated cohorts the test reaches 80% power at $N \approx 30$ endpoints near the attractor; the near-term signal is smaller and needs more patients. One caveat is worth pre-empting: a prior antidepressant meta-analysis found no excess variance (Munkholm et al.[57]), but VR$\approx 1$ does not exclude a mixture, and the same FDA data under finite-mixture modeling do show patients moving between discrete response classes (Stone et al.[58]). These population quantities depend strongly on the assumed baseline distribution (§S10).

### S7 Discontinuation and relapse: a qualitative test of fold versus graded

*This section supports the main-text claim (§3.4) that the SSRI discontinuation literature discriminates the bistable fold from the graded null* qualitatively*, and explains why we do not push that comparison to a quantitative fit.*

#### S7.1 Opposing predictions

The two models diverge sharply on what happens when treatment stops. In the graded (indirect-response) null the treated weight is $G^*_{\text{CS}} = G^0_{\text{CS}}/(1 + b\,\Delta e_C)$; setting the drug to zero returns $G_{\text{CS}} \to G^0_{\text{CS}}$ for *every* responder, so improvement is entirely drug-maintained, no off-drug remission is possible, the relapse hazard is roughly constant, and the relapse-free survival curve decays toward zero. In the fold, a responder whose OCD attractor was *annihilated* during treatment (carried across the saddle) sits in the health basin, which remains a fixed point at zero drug (that patient is durably remitted) whereas a responder who merely descended a lowered OCD branch (never crossed) climbs back. The population therefore splits; and because patients left just across the saddle relax slowly (critical slowing) while those far across never relapse, the relapse hazard *decreases* over time and survival *plateaus* at the durable fraction. The graded-null model *as formulated* therefore predicts that every responder relapses and the survival curve decays toward zero; the bistable-fold model predicts a durable off-drug subgroup and a plateau. These are the two trends we compare against the trials below.

#### S7.2 The data

Fineberg et al.[52] treated 468 patients with open-label escitalopram for 16 weeks; the 320 responders (Y-BOCS reduction $\geq 25\%$; randomization Y-BOCS $11.2 \pm 5.3$ in the placebo arm) were randomized to placebo or continued drug for 24 weeks. We digitized their Kaplan–Meier curve (their Fig. 3). The placebo (discontinued) arm does *not* decay to zero: its relapse hazard falls from $\approx 0.06$ wk$^{-1}$ near week 4 to $\approx 0.005$ wk$^{-1}$ by week 24 (late/early ratio $\approx 0.3$), and survival plateaus at $\approx 0.46$, a durable off-drug remission subgroup of about half; the continued-drug arm plateaus higher, at $\approx 0.76$. A meta-analysis of nine OCD discontinuation trials ($n = 1084$) finds the same qualitative pattern: discontinuation raises relapse (absolute risk increase $\approx 21\%$, number-needed-to-treat 5) while a majority nonetheless remain relapse-free off drug (Kishi et al.[59]). Both features (a durable subgroup and a *decelerating* hazard) are what the fold predicts and the graded null forbids.

#### S7.3 The model reproduces the shape

To compare, we initialize patients at the observed enrolled state (the per-patient gain $\alpha$ from a baseline $\mathcal{N}(26.4, 3.7)$ and the current weight $G_{\text{CS}}$ from a randomization Y-BOCS $\mathcal{N}(11.2, 5.3)$) and run the model's discontinuation ($u = 0$) and continued-drug dynamics, scoring relapse as a $\geq 5$-point Y-BOCS rise (Fineberg's definition). The model's off-drug survival curve *decelerates and plateaus* (Fig. S7a), for exactly the fold reason: patients near the saddle relapse slowly, those below it never do. A constant-hazard reversible model gives an exponential curve with no plateau. The model therefore reproduces the *shape* of the observed relapse dynamics, the qualitative fold signature.

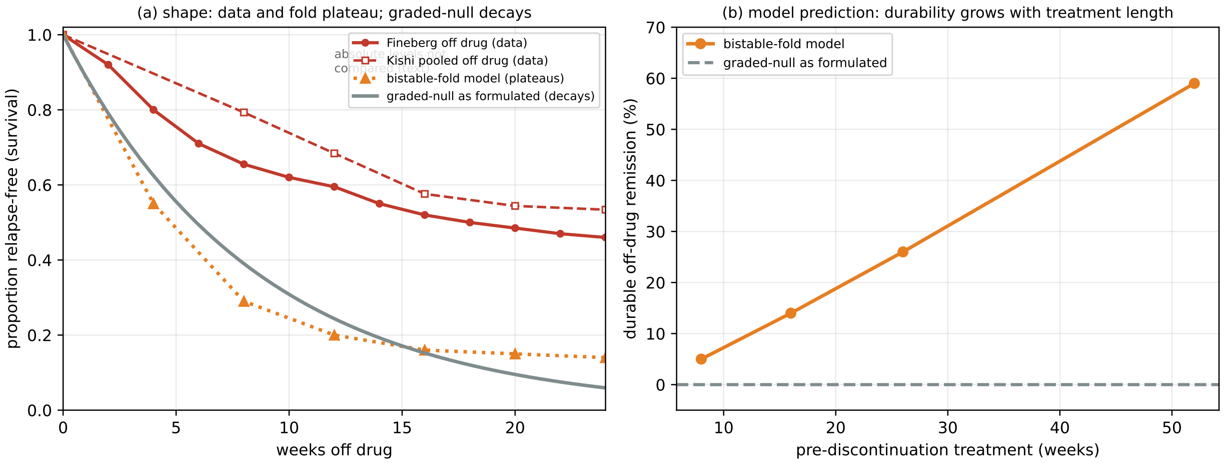

Figure S7: **Discontinuation and relapse: the trend comparison.** *(a)* Relapse-free (Kaplan–Meier) survival off drug. The data, Fineberg et al.[52] (digitized from their Fig. 3) and the nine-trial pooled rates of Kishi et al.[59] (derived from the reported weekly risk ratios and absolute risk reductions), *plateau*. The bistable-fold model, run from the observed enrolled state ($\alpha \sim \mathcal{N}(26.4, 3.7)$, randomization $Y \sim \mathcal{N}(11.2, 5.3)$), also plateaus, whereas the graded-null model *as formulated* (fully reversible) decays toward zero without a plateau. Only the *shape* (plateau vs decay) is compared here; the model's absolute survival level lies below the data and is not compared (it depends on the uncertain $\theta_M$, placebo representation, and readout; see text). *(b)* A model prediction: the fold's durable off-drug fraction rises with the duration of treatment *before* discontinuation (slow corticostriatal descent carries more patients across the saddle), whereas the graded-null model as formulated gives 0% at any duration. This duration trend is not yet tested.

#### S7.4 Why we keep it qualitative

The durable *fraction*, unlike the shape, is not robustly determined, because it turns on three ingredients the model does not pin precisely. (i) The metaplastic threshold $\theta_M$ sets where the separatrix falls on the Y-BOCS scale, hence how deep a descent locks in durability. (ii) The expectation (placebo) response is not mechanistically resolved: Fineberg's cohort reached Y-BOCS $\approx 11$ through a $\approx 15$-point *total* open-label improvement, whereas the model's drug-*attributable* effect is only $\approx 5$ points, so whether the extra descent is genuine plasticity (crossing $\rightarrow$ durable) or symptom suppression within the OCD basin (no crossing $\rightarrow$ fragile) is unresolved. (iii) The firing-rate-to-Y-BOCS readout fixes which $G_{\mathrm{CS}}$ corresponds to Y-BOCS 11. These are not small levers: two defensible specifications bracket the observed $\approx 50\%$ durable fraction from $\approx 14\%$ (severe patients *placed* at Y-BOCS 11, which the calibrated separatrix puts above the saddle) to $\approx 62\%$ (patients *driven* to low Y-BOCS by descent, which requires crossing). We therefore make no quantitative claim about the durable fraction.

#### S7.5 What is robust

The parameter uncertainty leaves two results standing, one a comparison and one a prediction. First, the *qualitative comparison*: a durable off-drug subgroup and a plateauing relapse curve appear in the data and in the bistable-fold model, and the graded-null model produces neither (fully reversible, it returns every responder toward baseline). Second, a *model prediction*: because the corticostriatal descent is slow ($\tau_G \sim 12$ wk), the durable fraction in the fold grows with the duration of treatment before discontinuation, in the model from $\approx 14\%$ at 16 weeks to $\approx 60\%$ at one year (Fig. S7b), as sustained treatment carries more patients across the saddle (a slow taper adds a few points; the $\sim 2$-week autoreceptor tail after abrupt cessation is negligible, as transporter re-opening dominates the serotonin fall), whereas the graded-null model produces no durable subgroup at any duration. This duration trend is not yet tested; a *duration-stratified* re-analysis of existing discontinuation trials would test it. On the trends these studies reveal, then, the graded-null model fails and the bistable-fold model captures them.

*Scripts:* `fineberg_km_digitized.py` (Fig. 3 digitization), `fig_relapse_km.py` / `fineberg_conditional_km.py` (enrolled-state KM), `discontinuation_relapse.py` and `taper_fast.py` (duration and taper), `autorecept_disc.py` (autoreceptor tail).

### S8 The plasticity source form: what creates the fold, and what only positions the attractor

#### S8.1 Overview

The bistable and the graded accounts of SSRI treatment share every element (the pharmacokinetic engine, the serotonin coupling, the circuit map $\Phi$, the readout $Y = 40\,p^{\gamma}$, and the per-patient placement of the untreated attractor) except one: the plasticity *source*, the production term in

$$\tau_G \dot{G_{\mathrm{CS}}} = \alpha\, S(G_{\mathrm{CS}}) - \big(1 + b\,(e_C - e_C^h)\big) G_{\mathrm{CS}}. \tag{3}$$

This section asks, precisely and quantitatively, which structural feature of the disease each ingredient of the source is responsible for. A nested family of sources is compared against the same saturating circuit map $\phi_{d_1}(G_{\mathrm{CS}})$ that both accounts use. The three structural features of the model, the bistable fold, the health basin, and the clinical placement of the OCD state, arise from three different ingredients, and only one of them is a modeling choice introduced here. The forms are defined first, with their provenance, and then evaluated.

#### S8.2 Model forms and their provenance

All variants share Eq. (3) and differ only in the source $S$. Throughout, $\phi_{d_1}(G_{\mathrm{CS}})$ is the D1 medium-spiny-neuron firing rate returned by the fast circuit, a saturating sigmoid with ceiling $\phi_{\mathrm{sat}} = Q_{d1} = 65$; $\phi_{d_1}^h = 7.4$ is the healthy D1 rate; $\theta_M = 9.15$ is the LTD/LTP crossover; and $\phi_e$ is the near-constant presynaptic (corticostriatal) drive.

##### (i) Threshold-free Hebbian.

$$S_{\mathrm{lin}}(G_{\mathrm{CS}}) = \phi_e\,(\phi_{d_1} - \phi_{d_1}^h). \tag{4}$$

The elementary correlational rule: the rate of change of the synaptic weight is proportional to the product of presynaptic drive and postsynaptic activity, referenced to the healthy rate $\phi_{d_1}^h$ so that the healthy state is a zero of the source. It has neither a modification threshold nor a saturation. An unconstrained correlational rule of this kind is intrinsically unstable, growing without bound, which is the classical motivation for adding either a sliding threshold or a bounding constraint (Miller & MacKay[60]; van Rossum et al.[61]). It is included here as the minimal source, to isolate what the further ingredients contribute.

##### (ii) BCM with a frozen threshold.

$$S_{\mathrm{BCM}}(G_{\mathrm{CS}}) = \phi_e\,(\phi_{d_1} - \phi_{d_1}^h)(\phi_{d_1} - \theta_M). \tag{5}$$

The Bienenstock–Cooper–Munro rule (Bienenstock et al.[28]; reviewed by Cooper & Bear[35]). The postsynaptic factor $(\phi_{d_1} - \theta_M)$ reverses sign at a modification threshold $\theta_M$, so that activity below $\theta_M$ drives depression and activity above it drives potentiation. The baseline reference $(\phi_{d_1} - \phi_{d_1}^h)$ is retained so that the healthy state remains a fixed point. In this variant $\theta_M$ is held fixed at its calibrated value, that is, metaplasticity is treated as quasi-static on the treatment timescale.

**(iii) BCM with a soft ceiling (the source used in the main text).**

$$S_{\text{cubic}}(G_{\text{CS}}) = \phi_e\,(\phi_{d_1} - \phi^h_{d_1})(\phi_{d_1} - \theta_M)(\phi_{\text{sat}} - \phi_{d_1}). \tag{6}$$

The BCM source multiplied by a soft upper bound $(\phi_{\text{sat}} - \phi_{d_1})$ that attenuates potentiation linearly as the postsynaptic rate approaches its ceiling $\phi_{\text{sat}}$. Multiplicative, activity-dependent soft bounds of this kind are a standard means of stabilizing an otherwise-unbounded Hebbian rule without hard clipping: the factor is exactly the $\mu = 1$ multiplicative rule of Gütig et al.[62] whose potentiation scale is $(1 - w)$, and the weight-dependent attenuation of van Rossum et al.[61], here applied to the postsynaptic firing rate rather than to the synaptic weight. The same bounded-response device appears independently in pharmacodynamics, where indirect-response models are given explicit physiological ceilings (Yao et al.[63]). This soft bound is used, in place of BCM's sliding threshold (iv), as the route to a bounded source; the reason is given in Section S8.5.

**(iv) Canonical BCM with a sliding threshold.** In the original BCM theory the threshold is not fixed but slides with recent postsynaptic activity: $\theta_M = (\bar{\phi}_{d_1}/\phi_0)^p\,\bar{\phi}_{d_1}$, with $\phi_0$ and $p$ "two fixed positive constants" and $\bar{\phi}_{d_1}$ the time-averaged rate (Bienenstock et al.[28]), equivalently $\theta_M \propto \langle \phi^2_{d_1} \rangle$ in the modern statement (Cooper & Bear[35]). Taking $p = 1$ and realizing the running average as a first-order relaxation of the threshold,

$$\tau_\theta\,\dot{\theta}_M = \frac{\phi^2_{d_1}}{\phi_0} - \theta_M, \tag{7}$$

gives at equilibrium $\theta^*_M = \phi^2_{d_1}/\phi_0$ (metaplastic time constant $\tau_\theta$), so the LTD/LTP balance $(\phi_{d_1} = \theta_M)$ sits at $\phi_{d_1} = \phi_0$. In BCM, $\phi_0$ is a *free* scale constant, not necessary for selectivity, set experimentally, and measured above spontaneous activity rather than being a physiological baseline (Bienenstock et al.[28]), so it sets *where* the homeostatic equilibrium lies. *Setting* $\phi_0 = \phi^h_{d_1}$ *is our modeling choice*, which places that equilibrium at the healthy rate; a larger $\phi_0$, at the elevated rate, would instead make the OCD state the metaplastic equilibrium of the same rule (§S8.5). This single-threshold homeostatic rule is moreover distinct from the two-threshold, dopamine-gated striatal metaplasticity of Trpevski[48] and Khodadadi et al.[49], whose sliding thresholds stabilize individual synaptic *weights* during learning (§S5.1); citing them here is not a claim that they instantiate Eq. (7). This variant restores the homeostatic threshold that (ii) and (iii) suppress.

**(v) Indirect-response null (constructed to remove the fold).**

$$S_{\text{IR}}(G_{\text{CS}}) = \frac{G_{\text{CS}}}{1 + G_{\text{CS}}/G_m}. \tag{8}$$

Unlike (i)–(iv) this is not a plasticity rule. It is a monotone, saturating source constructed for one purpose: so that the model has a single stable state and no fold, to serve as the fair "delete-the-fold" null. The null is the fold model with its non-monotone plasticity source replaced by a monotone one, and everything else (the drug engine, the readout, the per-patient placement) held identical. The functional form is taken from the indirect-response (turnover) framework of pharmacodynamics, a response variable with production and first-order loss on which the drug acts (Dayneka et al.[64]; Sharma & Jusko[65]), here with a saturable production term. It carries no mechanistic claim about plasticity; it exists only as a fold-free comparator.

#### S8.3 Result 1: the fold is generic to the saturating circuit, not to the added polynomial

Placing each source so that the untreated equilibrium sits at a patient's baseline (setting $\alpha$ from $\alpha\, S(G^0_{\text{CS}}) = G^0_{\text{CS}}$ at $G^0_{\text{CS}} = G_{\text{CS}}(Y_0)$) and enumerating the drug-free fixed points of Eq. (3) gives Table S2. Every biophysically grounded source in the family, including the bare threshold-free rule (i) with no $\theta_M$ and no ceiling, is *bistable* and folds under treatment (Fig. S8a). This follows from the structure: because $\phi_{d_1}(G_{\text{CS}})$ saturates, the ratio $S(G_{\text{CS}})/G_{\text{CS}}$ is non-monotone for any of these sources, so the linear sink line becomes tangent to it at a saddle-node. The fold is a property of a Hebbian source read through a saturating input–output function, and it is present before the BCM threshold or the ceiling factor is added. It is not created by the added polynomial.

Therefore, the bistable fold is not a special ingredient chosen in preference to a graded alternative; it is what a plasticity source generically produces in this circuit. As the following sections show, one must leave the plasticity mechanism entirely, and adopt a purely descriptive monotone dose–response, to obtain a persistent disease state without a fold.

#### S8.4 Result 2: the threshold sets the separatrix, the ceiling sets the severity of the attractor

Although the three static sources (i)–(iii) all fold, they differ in *where* they place the two boundaries that make a bistable disease clinically usable: the health basin (the separatrix) and the OCD attractor.

**The threshold sets the separatrix.** Strip it (the threshold-free source) and the health basin is uncontrolled: the saddle falls wherever the sink happens to cross, and the mildest patients cannot be held as a stable state at all (placing $Y_0 = 16$ yields an unstable equilibrium that rolls up to $Y \approx 18.5$; Table S2). Restoring the BCM factor $(\phi_{d_1} - \theta_M)$ fixes the depression region below $\theta_M$, which makes the healthy state robustly stable and defines the subclinical commitment threshold used in the main text.

**The ceiling sets the severity of the attractor.** With the BCM threshold but no ceiling (the quadratic source ii), the potentiating branch grows without an upper brake until $\phi_{d_1}$ nears its firing ceiling, so the OCD attractor *jams* near maximal severity ($Y \approx 37$). A subtlety is worth making explicit, because it is easy to misread. Choosing $\alpha$ to place a fixed point at a given baseline does not by itself make that point stable: stability is set by whether the source rises faster or slower than the linear sink there. For the quadratic source at $Y_0 = 24$ the potentiating term is still growing super-linearly (the unbraked LTP branch), so the source slope exceeds the sink slope and the placed point is a *saddle*; the trajectory runs upward until $\phi_{d_1}$ approaches its ceiling and the source finally plateaus, leaving the only stable disease state near $Y \approx 37$ (Fig. S8a, orange; Table S2). The soft ceiling $(\phi_{\text{sat}} - \phi_{d_1})$ bends the source over before $Y_0 = 24$, so that there the source slope is below the sink slope and the same placed point becomes a stable node; the attractor then tracks the patient's baseline across the clinical range $Y \in [16, 32]$ (red). The ceiling's role is therefore attractor *placement* and the reproduction of the observed severity spread, not the existence of the fold.

#### S8.5 Result 3: a sliding threshold is homeostatic: freezing it, or an adapted setpoint, is what sustains a persistent disease

Canonical BCM does not hold $\theta_M$ fixed; the threshold slides with recent activity (Eq. 7) toward a balance point at $\phi_{d_1} = \phi_0$, where $\phi_0$ is a free BCM constant (source iv), so its effect depends on where $\phi_0$ sits. With $\phi_0$ at the *healthy* rate (the choice we adopt above) the threshold homeostatically targets health: starting from the elevated state and letting $\theta_M$ track $\phi_{d_1}^2$, the threshold rises under the high firing, the source turns to net depression, and $G_{\text{CS}}$ collapses to the healthy setpoint (Fig. S8c), the faster the faster the threshold (crossing the remission line at about 30, 16 and 10 weeks for $\tau_\theta = 500, 150, 50$ weeks); the frozen-$\theta_M$ model used in the main text is the $\tau_\theta \to \infty$ limit, in which the attractor persists. With $\phi_0$ instead at the *elevated* rate, the balance point is the OCD state itself, and the sliding threshold sustains rather than erases it.

This has two consequences. First, a persistent pathological attractor requires *either* that the metaplastic threshold be effectively fixed on the disease timescale (large $\tau_\theta$) *or* that its setpoint $\phi_0$ have adapted to the elevated activity; only a homeostatic neuron whose setpoint is *health* cannot sustain OCD. Freezing $\theta_M$ is the conservative stand-in for both routes, and it localizes the disease's persistence in the pinning (or adaptation) of the LTD/LTP separatrix. Second, this is a timescale-separation (quasi-static) assumption, not a claim that metaplasticity is abnormal in OCD. Cortical plasticity measured by theta-burst TMS is in fact normal in OCD patients (Suppa et al.[30]), so the frozen threshold is not attributed to a metaplastic deficit; it is treated as a slow variable held fixed over the weeks-to-months treatment window. That the model would predict spontaneous remission were $\theta_M$ to track activity quickly *toward a healthy setpoint* is a structural, falsifiable commitment; a sliding threshold with $\phi_0$ adapted to the elevated state and a slow $\tau_\theta$ (under which the OCD state is itself the metaplastic equilibrium) is a route by which the fold might be *derived* from canonical BCM rather than imposed, which we leave to future work.

#### S8.6 Result 4: only a descriptive dose–response is graded

The only member of the family that yields a persistent disease *without* a fold is the indirect-response null (v), which is monotone by construction and therefore has a single stable state that slides smoothly with the drug (Fig. S8b). This is the point of its construction: it is not a plasticity rule but a phenomenological turnover source, adopted so that a fold-free but otherwise-matched comparator exists at all.

Of the whole family, then, only two members are able to place a persistent attractor across the clinical severity range: the cubic (which folds) and the indirect-response null (which does not). Only these two reach the calibration stage; sources (i) and (ii) are disqualified first by their placement pathologies, and the sliding-$\theta_M$ source (iv) by having no persistent disease at all.

To keep the comparison fair, each of the two is calibrated independently to the same thirteen SSRI arms, *including its own drug coupling $b$*. Borrowing a single $b$ would prejudge the comparison: the drug enters only through the sink term $(1 + b(e_C - e_C^h))G_{\text{CS}}$, and the two sources need different couplings to match the same trajectories. The cubic uses its main-text calibrated value ($b = 0.0268$); the null, refit with its own $(G_m, \tau_G, b)$, prefers a coupling about a quarter as large ($b = 0.0070$, ratio 0.26), a faster timescale ($\tau_G = 3.6$ wk), and a nearly linear source ($G_m$ at its upper search bound, so the null's parameters are only loosely determined, though its qualitative graded character does not depend on their exact values). Because $b$ enters only the sink, it affects only the treatment-response comparison (Fig. S8b), not the drug-free structure in panels (a) and (c); each curve in panel (b) is drawn with that model's own $b$.

Table S2: Behavior of the plasticity source family against the same saturating circuit. "Folds" and the fixed points are drug-free, with $\alpha$ placed so the untreated equilibrium sits at the patient's baseline where possible.

| Source | Folds? | Health separatrix | OCD attractor | Persistent disease? | Literature basis |
|---|---|---|---|---|---|
| (i) Threshold-free Hebbian | yes | uncontrolled | moderate ($Y$20–28); mild fails | yes | unstable Hebbian (Miller & MacKay[60]) |
| (ii) BCM, frozen $\theta_M$ | yes | set by $\theta_M$ | jams at ceiling ($Y \approx 37$) | yes | BCM (Bienenstock[28]) |
| (iii) Our cubic (+ soft ceiling) | yes | set by $\theta_M$ | clinical ($Y$16–32) | yes | BCM + soft bound (Gütig[62]; van Rossum[61]) |
| (iv) BCM, sliding $\theta_M$ | n/a | homeostatic setpoint | self-cures | **no** | canonical BCM (Bienenstock[28]; Cooper & Bear[35]) |
| (v) Indirect-response null | no (monostable) | none | graded slide | yes | constructed null; form from Dayneka[64] |

Given its own coupling, the null fits the trajectory means essentially as well as the fold (plain RMS 0.90 versus 0.83 Y-BOCS points across the 84 arm-timepoints; inverse-variance $\chi^2 = 6.50$ versus 6.27). It immediately follows that the population means are a degenerate observable that a bistable fold and a monotone dose–response fit alike, so they cannot decide between them. *The fold's case therefore rests not on the means or on the fit but on the individual-level signatures developed in the main text (discontinuous and bimodal endpoints, a durable off-drug remission subgroup, and pre-transition critical slowing), which the graded null cannot produce.*

#### S8.7 Summary

The three structural features of the account have three distinct origins. The fold comes from reading a Hebbian source through the saturating D1 transfer function, and is present even for a threshold-free rule; it is generic, not a consequence of the added polynomial. The BCM threshold $\theta_M$ sets the health basin and the commitment threshold. The soft ceiling $(\phi_{\text{sat}} - \phi_{d_1})$, a recognized stabilizing device rather than a classical BCM feature, sets where the OCD attractor sits on the severity scale. Canonical sliding-$\theta_M$ homeostasis does not give a graded disease; it gives no persistent disease, so freezing $\theta_M$ (a slow-variable, quasi-static assumption consistent with normal cortical plasticity in OCD) is what a persistent attractor requires. The only fold-free yet persistent model is a descriptive indirect-response dose–response, constructed as a null, which abandons the plasticity mechanism. The bistable fold is thus the plasticity mechanism's own prediction, and the graded alternative is available only by leaving that mechanism behind.

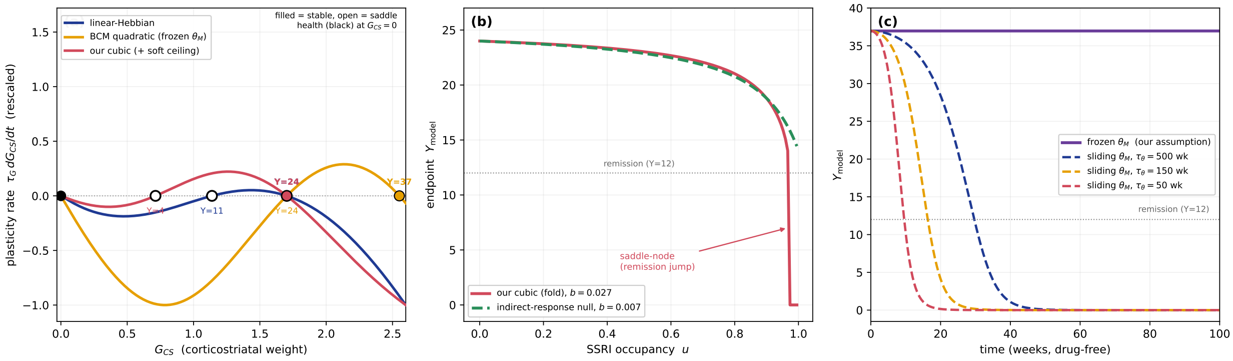

Figure S8: **What each ingredient of the plasticity source controls.** *(a)* Drug-free phase lines $\tau_G \dot{G}_{\text{CS}}$ (rescaled) for the three static sources, each placed for a baseline-$Y24$ patient. Filled circles are stable states, open circles saddles, black the healthy state at $G_{\text{CS}} = 0$. All three fold; the threshold-free (i, blue) and cubic (iii, red) sources place the OCD attractor near $Y24$, while the frozen-$\theta_M$ quadratic (ii, orange) jams it near the firing ceiling ($Y \approx 37$). *(b)* Endpoint $Y$ versus SSRI occupancy $u$, each curve drawn with that model's own calibrated $b$: the cubic crosses a saddle-node (a discontinuous remission jump) whereas the indirect-response null slides smoothly. *(c)* Drug-free $Y(t)$ from the elevated state: with $\theta_M$ frozen the disease persists, but a sliding (homeostatic) $\theta_M$ self-cures it, faster for smaller $\tau_\theta$. The frozen model is the $\tau_\theta \to \infty$ limit.

### S9 The observational exponent $\gamma$ and the ghost-zone fold

The observational bridge (main text §2.1) reads symptoms from the caudate rate through $Y_p = 40\left[(\phi_{d_1} - \phi^h_{d_1})/(Q_{d_1} - \phi^h_{d_1})\right]^\gamma$, a power law whose two anchors are circuit quantities and whose single exponent $\gamma$ sets the curvature between them. This section explains why $\gamma$ cannot simply be read off the trajectory fit, and how we fix it.

**A structural fold sets a floor on the representable severity.** The bistable corticostriatal plasticity of §2 is born in a saddle-node fold: below a critical excess weight $G^{\text{fold}}_{\text{CS}}$ there is no stable *elevated* (disease) attractor, only the healthy fixed point. The fold sits where the plasticity source $S(G)$ has *unit elasticity*. Here $S'(G) = \mathrm{d}S/\mathrm{d}G$ is the local slope of the source, and its elasticity $E(G) \equiv G\,S'(G)/S(G)$ is the fractional change in $S$ per fractional change in $G$ (equivalently $\mathrm{d}\ln S/\mathrm{d}\ln G$): $E > 1$ where the source rises faster than linearly, $E < 1$ where it rises more slowly. At $E(G) = 1$ the source's slope $S'(G)$ equals its secant slope $S(G)/G$, so the production curve $\alpha S(G)$ is tangent to the linear removal line ($\propto G$), precisely the saddle-node (fold) condition. Because unit elasticity is a property of the *shape* of $S$ alone, the fold is a structural invariant: its location in $G_{\text{CS}}$ (equivalently, in the caudate rate $\phi_{d_1}$) is essentially independent of $\gamma$, of the sinks, and only weakly dependent on $\theta_M$. What *does* depend on $\gamma$ is where that fixed fold lands on the clinical Y-BOCS axis. Because the fold occurs at a normalized caudate rate $p_{\text{fold}} = (\phi^{\text{fold}}_{d_1} - \phi^h_{d_1})/(Q_{d_1} - \phi^h_{d_1}) < 1$, and $Y_p \propto p^\gamma$, a larger $\gamma$ maps the same fold to a *lower* Y-BOCS: $Y_{\text{fold}}(\gamma) = 40\,p^\gamma_{\text{fold}}$ decreases monotonically with $\gamma$.

**The treatment-driven saddle-node, explicitly.** Figure S9 shows the bifurcation directly, rather than through the vertical phase-line slices of main-text Fig. 5. For a representative patient ($Y_{\text{model},0} = 20$), continuing the fixed points of the reduced scalar dynamics $\tau_G \dot{G}_{\text{CS}} = \alpha\,S(G_{\text{CS}}) - \big(1 + b(e_C(u) - e^h_C)\big)G_{\text{CS}}$ as SERT occupancy $u$ rises gives three branches: the stable OCD attractor, the unstable saddle, and the stable healthy state. As $u$ increases the drug steepens the linear sink; the OCD attractor and the saddle move toward each other and *collide and annihilate* at $u_{\text{fold}} = 0.82$ (the saddle-node), beyond which only the healthy state remains and the patient is carried to remission (panel a). Repeating the continuation across baseline severities traces the fold occupancy $u_{\text{fold}}(Y_0)$ (panel b): it rises from 0.45 at $Y_0 = 16$ through 0.82 at $Y_0 = 20$ to 0.97 at $Y_0 = 24$, crossing the maximal attainable SSRI occupancy ($u \approx 0.95$) near $Y_0 = 23$, so an SSRI alone folds the mildest patients (up to $Y_0 \approx 23$) and cannot reach the rest, the structural origin of severity-gated remission.

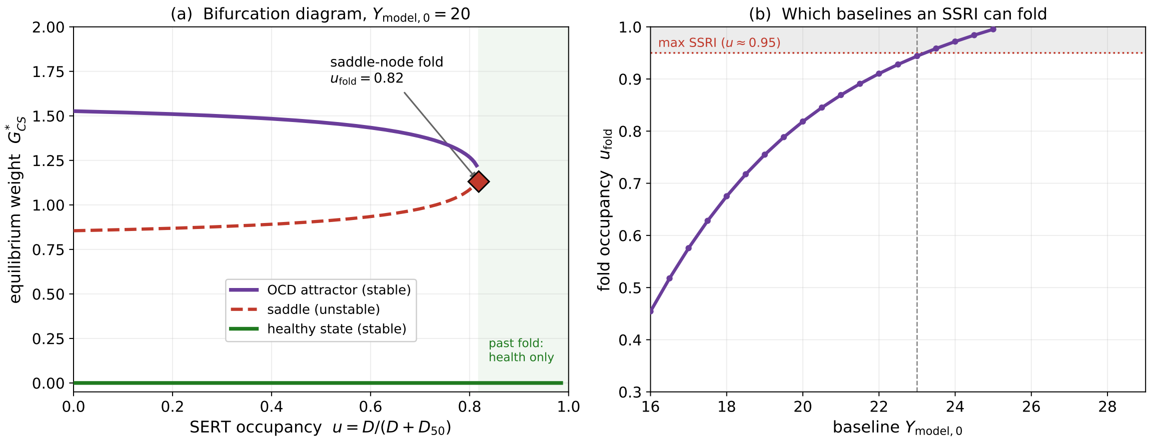

Figure S9: **The treatment-driven saddle-node bifurcation.** *(a)* Equilibrium corticostriatal weight $G^*_{\text{CS}}$ versus SERT occupancy $u$ for a patient presenting at $Y_{\text{model},0} = 20$, with the stable OCD attractor (solid), the unstable saddle (dashed), and the stable healthy state ($G^*_{\text{CS}} = 0$) continued explicitly. The OCD and saddle branches meet and annihilate at the saddle-node fold ($u_{\text{fold}} = 0.82$, diamond); past it (shaded) only health is stable. *(b)* The fold occupancy $u_{\text{fold}}$ versus baseline severity: an SSRI (maximal $u \approx 0.95$, dotted) folds baselines up to $Y_0 \approx 23$ and cannot reach more severe attractors. Script `fig_bifurcation.py`.

**Normal-form certification.** That the collision in Fig. S9 is a saddle-node, rather than a numerical near-miss, we certify directly. Writing the reduced right-hand side as $f(G_{\text{CS}}, u) = \alpha\, S(G_{\text{CS}}) - \big(1 + b(e_C(u) - e_C^h)\big)G_{\text{CS}}$ (its zeros in $G_{\text{CS}}$ are the fixed points), a saddle-node at $(G^*_{\text{CS}}, u^*)$ requires three standard conditions, each with a plain meaning here. *Tangency* ($f = \partial f/\partial G_{\text{CS}} = 0$): the two colliding fixed points (the OCD attractor and the saddle) have merged into a single point, where the plasticity-rate curve just *touches* zero rather than crossing it. *Non-degeneracy* ($\partial^2 f/\partial G_{\text{CS}}^2 \neq 0$): that contact is a clean quadratic turning point (a fold), not a flatter, higher-order touching that would behave differently. *Transversality* ($\partial f/\partial u \neq 0$): varying the control parameter (SERT occupancy $u$) actually drives the two fixed points together and through annihilation, rather than leaving them stationary. At the calibrated $Y_0 = 20$ fold ($u^* = 0.818$, $G^*_{\text{CS}} = 1.176$) we find $f = 3 \times 10^{-8}$, $\partial f/\partial G_{\text{CS}} = -8 \times 10^{-6}$ (both zero to numerical tolerance), $\partial^2 f/\partial G_{\text{CS}}^2 = -3.14$ and $\partial f/\partial u = -0.89$ (both safely nonzero). The dynamics near the fold therefore reduce to the topological normal form $\dot{x} = \tau_G^{-1}(a\,x^2 + b\,(u - u^*))$ with $a = -1.57$, $b = -0.89$, and the relaxation rate obeys the associated critical-slowing law $\lambda(u) \propto \sqrt{u^* - u}$: the computed $\lambda(u)/\sqrt{u^* - u}$ approaches a constant ($\approx 0.22$ wk$^{-1}$) as $u \to u^*$. Script `fold_certification.py`.

**Slow-manifold robustness of the quasi-steady-state reduction.** The scalar dynamics above take the extracellular serotonin $e_C$ at its desensitized steady value $e_C(u)$, i.e. treat the autoreceptor desensitization (timescale $\tau_{\text{des}} \approx 2$ wk) as fast relative to the plasticity remodeling ($\tau_G \approx 12$ wk). Because that separation is finite ($\sim 1/6$), we verify the reduction does not distort the fold by integrating the full two-timescale system, the plasticity ODE driven by the *time-resolved* $e_C(t)$ from the serotonin engine (which ramps over $\tau_{\text{des}}$), and comparing to the reduced (constant-$e_C(u)$) system. Across baselines $Y_0 \in [18, 26]$ and occupancies $u \in \{0.5, 0.8, 0.95\}$ the maximum 12-week endpoint discrepancy is 0.25 Y-BOCS points (most cells $< 0.1$), well within the 0.83-point calibration RMS, and the fold location for $Y_0 = 20$ shifts by less than one part in 400 ($u_{\text{fold}} = 0.818 \to 0.820$). The bistable fold thus survives intact when the desensitization is resolved as a second slow variable rather than adiabatically eliminated.

**The ghost zone.** If $Y_{\text{fold}}$ lies above the clinical entry floor ($Y \approx 16$), then a band of mild severities [16, $Y_{\text{fold}}$] has no stable elevated attractor: a patient presenting with such a score cannot be placed on the disease branch at all. We call this unrepresentable band the ghost zone, and we require the readout to eliminate it, $Y_{\text{fold}} \le 16$. This is a well-posedness condition on the observational map, not a fit criterion, and it is what pins $\gamma$ from below.

**What the trajectories do and do not constrain.** Holding the non-identified constants fixed, the modification threshold $\theta_M = 9.15\ \text{s}^{-1}$ (sub-clinical saddle, main text §2), and the autoreceptor gain $h_{\max} = 2$ (physiology; S4), we fit $b$ and $\kappa_{\text{des}}$ (with $\tau_G$ pinned) at each fixed $\gamma$ on the clean 13-arm set (Table S3). Here the *sum of squared errors* (SSE) denotes the pooled, inverse-variance-weighted, dimensionless trajectory discrepancy $\sum_{\text{arms}} W \sum_t w_t \, [(Y_{\text{model}} - Y_{\text{obs}})/\sigma]^2$ (lower is better). Two features fix $\gamma$, now from both sides. First, with $\tau_G$ pinned on clinical grounds the pooled SSE is essentially *flat* across $\gamma \in [1.5, 2]$ (6.28 vs 6.30, statistically indistinguishable) and then climbs steeply beyond it (to $6.9 \to 7.7 \to 9.2$ at $\gamma = 2.5, 3, 4$; Table S3), so the trajectories bound $\gamma$ from *above* but do not discriminate *within* $[1.5, 2]$. Second, the ghost-zone requirement supplies the lower bound: with the anchored $\theta_M$, the sub-clinical fold clears the clinical floor (excluded fraction $\to 0$) only at $\gamma \ge 2$: at $\gamma = 1.5$ a 2.2% sub-clinical wedge remains. The two bounds pincer onto the plain quadratic readout $\gamma = 2$ (the smallest exponent that closes the ghost zone and the largest the trajectory fit tolerates) which we adopt.

**$\gamma = 2$ is bracketed, not tuned to relocate the fold.** A reasonable concern is that $\gamma$ is chosen only to push the ghost-zone fold below the clinical floor. It is not tuned for that purpose: once $\tau_G$ is pinned (S4), the trajectory SSE independently caps $\gamma$ from above (flat to $\gamma = 2$, rising beyond), while ghost-closure sets the floor at $\gamma = 2$, so $\gamma = 2$ is fixed by the *intersection* of a data-driven upper bound and a hygiene lower bound rather than by the fold location alone. We do not claim the data *prefer* $\gamma = 2$ over $\gamma = 1.5$ (there they are statistically indistinguishable) only that $\gamma = 2$ meets both constraints. The drug coupling $b$ drifts only slightly across this range, and no downstream conclusion hinges on the precise curvature of the readout. The main text uses $\gamma = 2$ throughout.

Table S3: **The readout exponent $\gamma$ is bracketed: ghost-closure from below, fit quality from above.** At each fixed $\gamma$ the drug coupling $b$ and autoreceptor coupling $\kappa_{\text{des}}$ are fit on the clean 13-arm SSRI set, with $\theta_M = 9.15\ \text{s}^{-1}$ (saddle), $h_{\max} = 2$ (physiology) and $\tau_G = 12$ wk (clinical; S4) held. SSE is the pooled inverse-variance-weighted dimensionless trajectory discrepancy; "excluded" is the patient fraction below the plasticity fold (ghost-zone cost); $Y_{\text{fold}}$ is the Y-BOCS location of the saddle-node fold. The adopted $\gamma = 2$ is bold.

| $\gamma$ | SSE | excluded | $Y_{\text{fold}}$ | $b$ ($\text{nM}^{-1}$) |
|---|---|---|---|---|
| 1.5 | 6.28 | 2.2% | 16.1 | 0.024 |
| **2.0** | **6.30** | **0%** | **11.9** | **0.0268** |
| 2.5 | 6.90 | 0% | 8.8 | 0.027 |
| 3.0 | 7.68 | 0% | 6.5 | 0.028 |
| 4.0 | 9.18 | 0% | 3.6 | 0.029 |

### S10 Sensitivity of population-level predictions to the assumed baseline distribution

**Summary.** Three points organize this section. *(i) Mean versus width.* The population mean is the dominant control on the responder and remission *levels*: raising it carries patients above the fold and collapses both fractions monotonically. The baseline width $\sigma$ is a second-order, severity-dependent modulator, nearly inert at a mild mean, but *pro-response* at a severe mean, where a broader distribution recruits a low-baseline tail back below the fold. In short, $\mu$ sets the level and $\sigma$ only adjusts it, and materially so only when the mean is severe. *(ii) Bloch and Montgomery.* The model reproduces the dose-ranging meta-analytic responder band (Bloch et al.[66], 16–22%) only for a population mean of $\approx 28$, about 3–4 Y-BOCS points above the true trial mean ($\approx$24–25); the Montgomery population $\mathcal{N}(28, 4)$ is adopted as that reference cohort (Table S4). This constant gap survives changes of distribution *shape*, lower-tail *truncation*, and the onset timescale $\tau_G$, so it is read as a fixed *registration offset* of the firing-rate readout, not a distributional or fitting artifact. *(iii) The one parameter that would close it.* A steeper readout exponent $\gamma$ would map the fold to a lower Y-BOCS and remove the offset; but $\gamma$ is pinned to 2 by independent constraints (the ghost-zone floor from below and the trajectory-fit ceiling from above, §S9), so we accept the offset rather than tune $\gamma$ to absorb it.

Every population-level number in the paper (the drug-attributable responder fraction, the remission fraction, the mean $\Delta$Y-BOCS, and the endpoint BC/VR) is a single-patient response integrated over the assumed baseline Y-BOCS distribution (the deterministic quadrature of Methods, §5.3). Because the single-patient response is a steep, threshold-like function of baseline severity (the fold), these population numbers depend strongly on the assumed distribution $\mathcal{N}(\mu, \sigma)$. That dependence is not a nuisance to be hidden behind one chosen population: it is the severity-gating (main text §3.3) projected onto a cohort, and it separates cleanly into a *mean* effect and a *width* effect (Fig. S10).

*Mean (at fixed $\sigma$): the severity-gating collapse.* As the population mean rises, fewer patients sit below the fold, so the net $\geq$35% responder fraction falls monotonically, $\approx$53% at $\mu = 24$, 27% at $\mu = 28$, 16% at $\mu = 30$ ($\sigma = 5.5$), and remission falls with it (33% at $\mu = 24$ to 12% at $\mu = 28$, 6% at $\mu = 30$). The dose-ranging meta-analytic responder band (Bloch et al.[66], ARD 16–22%) is reproduced only near $\mu \approx$ 29–30 at this width ($\sigma = 5.5$), or near $\mu \approx 28$ at the narrower Montgomery baseline width ($\sigma = 4$; Table S4), in either case *above* the empirical trial mean ($\approx$ 24–25; the calibration arms span 23.5–27.0). At the empirical trial mean the model returns $\approx$46–53% where Bloch reports 16–22%; as the main text develops, this level gap is read as a $\approx$3–4-point registration offset of the firing-rate readout (§2.1) rather than a distributional failure, and it is distinct from the separate mismatch between the model's severity *slope* and the flat slope of Cohen et al.[47].

*The offset is a readout registration, not a distributional artifact.* Two checks show the $\approx$3–4-point gap between the Bloch-matching mean ($\mu \approx$ 28–29) and the clinical mean ($\approx$25) is not an artifact of the assumed lower tail. First, it is insensitive to distribution *shape*: replacing $\mathcal{N}$ with a uniform law over the same support, or pooling the thirteen calibration arms as a mixture of per-arm normals, both return $\approx$43% at the empirical mean, unchanged. Second, a trial that enrolled only above a Y-BOCS floor would truncate the lower tail and raise the effective mean; applying inclusion floors of 16, 18, 20 and 22 to the pooled mixture moves the responder fraction only to 41, 38, 33 and 24%, so the band is reached only at a floor of $\approx$22. Such a floor is excluded by the trials' own reported dispersion: truncating at 22 compresses the enrolled SD to $\approx$3.6, whereas the calibration arms report baseline SD $\approx$4–5.5, consistent only with a floor near 16–18 (at which the responder fraction is 38–41%). The offset is therefore a property of the firing-rate-to-Y-BOCS readout, which we do not tune away (the exponent $\gamma$ is fixed structurally (§S9), and a steeper $\gamma$ would close the gap) and not a hidden freedom in the baseline law.

*Width (at fixed $\mu$): the effect is severity-dependent.* Because the response is nonlinear in baseline, widening the distribution moves mass both below the fold (adding strong responders) and above it (adding non-responders); which dominates depends on where the mean sits. Near a mild mean ($\mu = 24$) the responder fraction is nearly flat in $\sigma$ ($56\% \to 50\%$ over $\sigma = 3 \to 7$); near a severe mean ($\mu = 28$) it *rises* with $\sigma$ ($12\% \to 30\%$), as a wider distribution pulls a low-baseline tail below the fold. The width matters most precisely where the mean places the bulk of the population above the fold.

*Robust vs sensitive.* What survives the entire $\mu \in [22, 30] \times \sigma \in [3, 7]$ grid is the *qualitative* severity-gating (responder and remission fractions fall with severity) and the fold-versus-graded *direction* (variance inflation, VR$> 1$, for the fold at every cell). What is population-*sensitive* is the exact responder/remission fraction, which we therefore report as ranges rather than single values; agreement with any particular meta-analytic point requires a particular assumed population and should not be read as calibration. (The complementary *time*-dependence of the endpoint BC/VR is in §S6.3.)

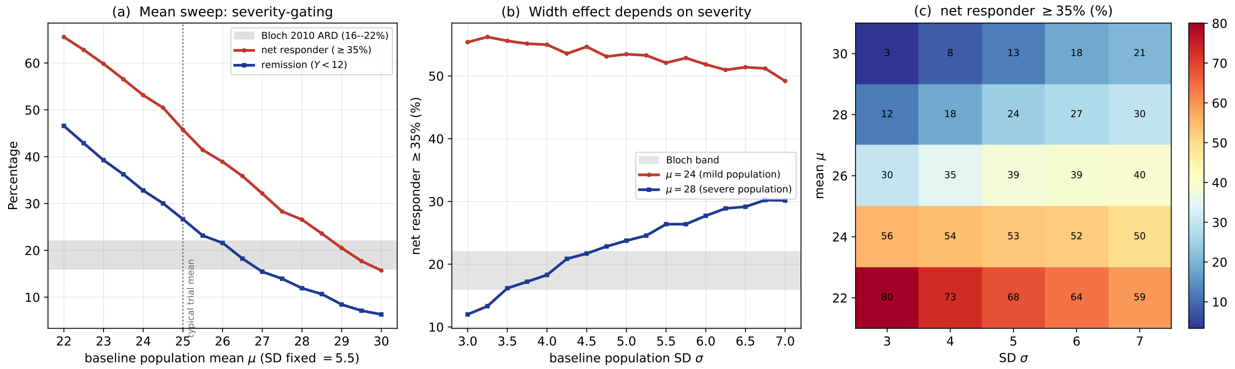

Figure S10: **Population-level predictions depend on the assumed baseline distribution $\mathcal{N}(\mu, \sigma)$.** *(a)* Mean sweep (fixed $\sigma = 5.5$): net $\geq 35\%$ responder and remission fractions fall with the population mean; Bloch's ARD band (16–22%, shaded) is reproduced only near $\mu \approx 28$, above the typical trial mean (dotted). *(b)* SD sweep: the width effect is severity-dependent: flat at a mild mean ($\mu = 24$), rising at a severe mean ($\mu = 28$). *(c)* Net $\geq 35\%$ responder fraction over the full $(\mu, \sigma)$ grid. Frozen parameters; representative SSRI dose. Script `fig_population_sensitivity.py`.

Table S4 reports the drug-attributable $Y_{\text{model}}$ drop and its spread at the Montgomery baseline population $\mathcal{N}(28, 4)$, the population that reproduces the meta-analytic responder band.

Table S4: **Population drug-attributable response** at the baseline population $Y_0 \sim \mathcal{N}(28, 4)$ ($\sigma = 4$ is the baseline Y-BOCS SD of Montgomery et al.[67], whose baseline distribution this matches on the $Y_{\text{model}}$ scale), where the model reproduces the meta-analytic responder band (Bloch et al.[66]), about three to four points above the empirical trial mean on the $Y_{\text{model}}$ scale, the readout's registration offset. Mean ± SD of the drug-attributable $Y_{\text{model}}$ drop, at two SERT occupancies and the 12- and 24-week endpoints; % is mean/28. The SD grows with time and dose as the responder/non-responder split widens (near-fold patients descend fast, severe ones barely move).

| SERT occupancy | week | mean $\Delta Y_{\text{model}}$ (pts) | SD (pts) | % of baseline |
|---|---|---|---|---|
| $u = 0.8$ | 12 | 1.83 | 0.65 | 7% |
| $u = 0.8$ | 24 | 2.04 | 0.93 | 7% |
| $u = 0.9$ | 12 | 3.04 | 1.09 | 11% |
| $u = 0.9$ | 24 | 3.46 | 1.67 | 12% |

*Robustness of the offset to the remodeling time $\tau_G$.* The $\approx$3–4-point registration offset is not an artifact of the assumed onset timescale. Fixing $\tau_G$ at 12, 15, 20 and 25 weeks and refitting the drug coupling $b$ to the 13-arm trajectories at each value, the 12-week responder fraction and the offset are essentially unchanged (46–47% at the empirical mean; offset 3.7–4.0 points): the descent velocity out of the OCD basin scales as $b/\tau_G$, so refitting $b$ preserves the 12-week response, while the readout map that sets the offset is a steady-state quantity blind to any timescale. The trajectory RMS mildly favors the shorter value (0.83 at 12 wk, rising to 0.90 at 25 wk), so $\tau_G$ is loosely constrained toward the adopted 12 weeks. The offset is therefore a property of the firing-rate-to-Y-BOCS readout, invariant to the onset timescale.

### S11 Reading a patient-level mechanism from population-level data

The model's severity-gating is a patient-level, nonlinear (threshold) prediction; the strongest population evidence is the individual-patient-data meta-analysis of Cohen et al.[47] (eleven double-blind, placebo-controlled short-term SSRI trials; $n = 2372$; mean baseline Y-BOCS 24.5, SD 4.5; 28% mild [$\leq 21$], 56% moderate [22–29], 16% severe [$\geq 30$]). Cohen reported two treatment endpoints and found *both* unmodified by baseline severity. The primary endpoint is the drug-attributable *mean difference* in Y-BOCS change between arms, 2.65 points favouring the SSRI (95% CI $[1.85, 3.46]$; SMD 0.33), with a baseline-severity×treatment interaction of $\beta = +0.071$ (95% CI $[-0.049, +0.192]$, $p = 0.22$). The secondary endpoint is *response*, a $\geq 35\%$ reduction in Y-BOCS *from baseline* scored per patient; this is a *placebo-inclusive* total change (drug, placebo, natural waning and regression to the mean all count), summarized as an active-versus-placebo odds ratio of 2.21 (95% CI $[1.72, 2.83]$) against a placebo responder rate of 0.18, its interaction likewise null ($\beta = +0.032$, $[-0.034, +0.099]$). Two features set the terms of any model comparison. First, the response criterion is placebo-inclusive, so a matched comparison must isolate the drug-attributable contrast rather than the model's drug-only crossing rate. Second, the estimate is *pooled* across unnamed SSRIs at various, often flexible doses (neither drug identity nor dose is reported) so the model can only stand in for this mixture at a single representative occupancy, and only the severity *slope* (the flatness), not the absolute level, is a well-posed target.

Rather than argue that the population instrument is simply blind to the mechanism, we state what the model actually predicts on this target, quantify the disagreement, and locate its origin. None of what follows impugns the validity or rigour of the meta-analysis, whose design and execution we do not question.

*The model makes a definite prediction here that differs from Cohen.* At a single representative occupancy the model's drug-attributable mean improvement falls with baseline severity, with a slope of $\approx -0.31$ Y-BOCS points per baseline point (range $-0.22$ to $-0.35$ across the SSRI dose range; §S12). In Cohen's coding (positive = drug helps *less* at higher baseline) this is +0.31, the same sign as Cohen's +0.071 but about 1.6× above the upper bound of Cohen's 95% interval (+0.192); even the weakest dose (+0.22) exceeds it. Over the mild-to-severe span ($\sim 15$ points) Cohen's interval permits a drug-benefit difference of at most $\sim 2.9$ points, whereas the model predicts $\sim 4.6$. The mismatch is, however, *localized and one-sided*: the model reproduces the severe end (drug-attributable $\sim 2.2$ points, close to Cohen's overall 2.65) and over-predicts the *mild*-patient benefit (about three-fold larger than at the severe end). The honest statement is therefore "the model over-predicts the benefit for mild patients," not "severe patients never respond."

*The mean-difference comparison is valid without a placebo model.* The between-arm mean difference cancels regression to the mean (present in both arms) and, if the drug acts approximately additively with the natural course, cancels spontaneous waning as well, so it can be set against the model's drug-attributable prediction directly. We verified the additivity: adding a natural-course sink that reproduces a placebo/waning trajectory changes the drug-attributable slope by less than 0.01 (from $-0.302$ drug-alone to $-0.296$ to $-0.299$), with a residual below $\sim 0.4$ points concentrated at the mild end. The severity gradient is thus a genuine prediction of the model, not an artefact of omitting the placebo arm.

*The pooled data narrow the gap in two ways.* First, dose is confounded with severity: in flexible-dose arms more severe patients are titrated to higher doses, so exposure rises with severity and a severity–dose gradient partly compensates the fixed-dose gating the model predicts (this bites hardest for the flexible-dose component; strictly fixed-dose arms avoid the within-trial confound but span a narrower severity range). Second, the tested endpoint is $\geq 35\%$ *response* at 10–13 weeks, whereas the model's sharpest severity signal is in *remission* (fold-crossing), which for milder patients completes only over the slower remodelling timescale (critical slowing); the response endpoint is also genuinely threshold-shaped, so a linear interaction term has reduced power against it. These narrow the discrepancy but do not on their own eliminate the over-prediction of mild benefit in the *mean*, which is approximately linear over the populated range and so is shielded by neither point.

*The residual gradient is not a signature of bistability.* This is what fixes the interpretation. The severity gradient is *not* peculiar to the fold: it persists, essentially unchanged, in every variant of the model we examined (the graded (fold-free) null of §S6, alternative monotone readouts, and metaplastic (sliding-threshold) plasticity rules) and traces to a feature they all share, that a more severe patient is placed on a stronger plasticity source and therefore moves proportionally less under a given serotonergic sink (§S12). Because the graded null over-predicts the *same* gradient, the mismatch with Cohen does not distinguish the fold from its alternatives and is not grounds to reject bistability. We therefore retain the bistable model on the individual-level signatures that a population mean cannot resolve, discontinuous and bimodal endpoints, a durable off-drug remission subgroup, and pre-transition critical slowing (§3.3, §3.4), while recording the mean-gradient over-prediction as a genuine, quantified limitation of the current model class, to be tested directly when individual-patient trajectories with known drug and dose become available.

### S12 Origin of the severity gradient: a model-class property, not a fold artefact

*This section supports §S11 and the main-text claim that the model's over-prediction of the severity gradient, relative to Cohen et al.[47], is shared by fold-free and alternative-readout variants and so does not weigh against bistability. Every quantitative statement below is a model computation at the calibrated operating point; the scripts are listed at the end.*

#### S12.1 The gradient lives in the dynamics, not the readout

The model's dynamics are entirely in the space of the corticostriatal weight $G_{\mathrm{CS}}$ (equivalently the $D_1$ firing rate $\phi_{d_1}$): the per-patient gain is fixed by the baseline balance $\alpha = G^0_{\mathrm{CS}}/S(G^0_{\mathrm{CS}})$, the plasticity source $S$ and its threshold $\theta_M$ are defined on $\phi_{d_1}$, and the serotonergic drive enters through the occupancy $u$: none of these refers to the Y-BOCS readout. The readout $Y = 40\,p^\gamma$ is a downstream, monotone relabelling of the endpoint. The drug-induced *movement* of a patient, and how it varies with baseline severity, is therefore a property of the map $G^0_{\mathrm{CS}} \mapsto \Delta G_{\mathrm{CS}}$; a readout can compress or stretch the axis afterwards but cannot create or remove the gradient.

That movement is steeply severity-dependent. At a strong SSRI dose the treated fall in $\phi_{d_1}$ is $\approx 25\%$ for a mild patient ($Y_{\mathrm{model},0} = 16$), $\approx 10\%$ at moderate (25), and only $\approx 2.6\%$ at severe (34); in the underlying weight $G_{\mathrm{CS}}$ the corresponding drug-driven excursions are $\approx 27\%$ versus $\approx 3.5\%$. A more severe patient sits on a stronger source and barely moves under a given sink. This is the gradient, present before any readout is applied.

#### S12.2 No readout removes it

Because the movement itself is severity-graded, no monotone readout $Y = F(\phi_{d_1})$ can flatten the drug-attributable severity profile without destroying the trajectory fit. We refit the two Y-BOCS-constrained constants ($\theta_M$ and the sink strength) at each of a family of readouts and recomputed both the pooled trajectory discrepancy $\chi^2$ and the drug-attributable fraction reaching $\geq 35\%$ improvement by stratum (a drug-only crossing rate, distinct from Cohen's placebo-inclusive response and used here only as a movement probe):

| readout $Y = F(\phi_{d_1})$ | $\chi^2$ | mild | moderate | severe |
|---|---|---|---|---|
| power $\gamma = 2$ (calibrated) | 6.18 | 41% | 0% | 0% |
| power $\gamma = 3$ | 6.23 | 33% | 0% | 0% |
| power $\gamma = 4$ | 6.67 | 27% | 0% | 0% |
| power $\gamma = 6$ | 7.73 | 16% | 0% | 0% |
| power $\gamma = 8$ | 8.60 | 12% | 0% | 0% |
| power $\gamma = 12$ | 9.77 | 7% | 0% | 0% |
| sigmoid ($k = 8$) | 6.13 | 27% | 0% | 0% |
| sigmoid ($k = 12$) | 6.24 | 33% | 0% | 0% |

Moderate and severe patients cross at 0% under *every* readout: raising the power exponent $\gamma$ only suppresses the mild rate (from 41% to 7%) without lifting the others, so a "flat" profile is reached only near 0% overall, never at a realistic rate, and the fit degrades monotonically. Sigmoidal readouts are the wrong shape (saturated exactly where severe patients live) and behave identically.

A readout *linear in the weight*, $Y \propto G_{\text{CS}}$, halves the per-patient gradient but does so by suppression (it drives the mild rate to zero as well), and even in the raw weight the drug-driven excursion is $\approx$3.5% (severe) versus $\approx$27% (mild); a Hill-in-$G_{\text{CS}}$ readout can flatten the slope only by mapping the OCD range onto Y-BOCS 2–13 (the wrong clinical range), and at the correct range it *steepens* the slope (to $-0.43$).

The one readout that comes closest is a self-consistent high-exponent map: refitting $\theta_M$, $\tau_G$ and the sink strength at $\gamma = 7$ fits the trajectories as well as the calibrated $\gamma = 2$ fold (RMS 0.82 Y-BOCS points in both). It still does not resolve the discrepancy. The best-fitting $\gamma = 7$ solution rails $\theta_M$ and $\tau_G$ to implausible values ($\theta_M \approx 45$ s$^{-1}$, $\tau_G \approx 30$ weeks, longer than any trial) and retains a severity slope of $-0.26$ (Cohen-coded $+0.26$, outside Cohen's interval); constraining the slope down to Cohen's boundary ($-0.19$) costs fit ($\chi^2$ from 6.2 to 8.2) and rails the same parameters. The readout exponent trades fit, flatness and parameter plausibility against one another; it cannot deliver all three.

##### S12.3 No plasticity rule, and no fold, removes it either

The gradient is equally insensitive to the two remaining structural choices. Replacing the constant modification threshold with the canonical activity-slaved *sliding* threshold (metaplasticity), swept across the full range from fast to frozen adaptation, never flattens the drug-attributable profile while preserving the graded dose–response (§S5.1). And the fold itself is not the culprit: the matched *graded* (fold-free) null of §S6, built from the identical pharmacokinetics, circuit map, readout and per-patient placement, over-predicts the *same* severity gradient, if anything a slightly steeper one. Because a model with no fold reproduces the gradient, the gradient cannot be a signature of bistability.

Moving the serotonergic action from the sink to the *source* does not help either. A proportional serotonergic reduction of the LTP gain, $\dot{\alpha} = F_\alpha - \alpha - b_\alpha(e_C - e_C^h)\alpha$, gives at drug steady state $\alpha_0\, S(G_{\text{CS}}) = (1 + b\,\Delta e_C)(1 + b_\alpha\,\Delta e_C)\, G_{\text{CS}}$: a source term multiplies the drug *potency* uniformly across patients and cannot change *who* crosses (§S1.5). Every serotonergic route we can write leaves the ordering (mild move most, severe least) intact.

##### S12.4 Patient heterogeneity does not flatten it either

The natural remaining proposal is that patient-to-patient variability smooths the transition. Patients do differ in their effective serotonergic sensitivity (through pharmacokinetic and metabolizer differences and serotonin-transporter genotype) so we let the sink coupling vary across patients, drawing $b_i$ from a mean-preserving lognormal distribution (coefficient of variation $\approx 0.4$). This broadens the distribution of individual outcomes, as expected: it softens the all-or-nothing crossing, so the spread of endpoints within a stratum widens. Because the distribution is mean-preserving in $b$, whether it also shifts the stratum-*mean* improvement is a question of the map's curvature (Jensen's inequality), not settled a priori; we therefore checked it directly. Averaging over $b_i$ leaves the stratum-mean drug-attributable improvement (and hence the severity slope that Cohen's null concerns) essentially unchanged, with the moderate and severe crossing rates staying at or near zero. Heterogeneity in $b$ is a realistic ingredient and the obvious candidate smoother, but it acts on the *variance* of response, not on the *mean* gradient.

##### S12.5 Root cause and consequence

The common origin is how baseline severity is represented. A patient's presenting severity is encoded in the plasticity source through the gain $\alpha$: a severe patient sits at a high baseline weight, which requires a stronger source, which then resists the proportional serotonergic sink $-b\,\Delta e_C\,G_{\mathrm{CS}}$ that treatment supplies. The drug therefore moves severe patients proportionally less, a structural consequence shared by the fold, the graded null, every monotone readout, and both placements of serotonin. The severity gradient is thus a property of this *model class*, not of the saddle-node fold. This carries two implications. First, the mismatch with Cohen's flat modifier does not discriminate the fold from its alternatives and gives no reason to abandon bistability; the discriminating evidence is the individual-level, dynamical signatures (§S6; main text §3.3, §3.4). Second, the over-prediction is a genuine, quantitative limitation of how this whole class maps severity to drug response; its natural resolutions (a weaker-than-proportional coupling for the most severe, or severity-dependent drug exposure) are targets for the individual-patient data that would test them.

*Scripts:* `readout_sweep.py` (readout family, $\chi^2$ and crossing rates), `step_readout_shape.py` and `step_gamma7_v2.py` (Hill-in-$G_{\mathrm{CS}}$ and the self-consistent $\gamma = 7$ refit), and `step1a_meandiff.py` (stratum-resolved drug-attributable slopes), `increment3_bhet.py` (lognormal $b$-heterogeneity), together with the sliding-threshold sweep of §S5.1.

### S13 Serotonin-syndrome thresholds and the ceiling on serotonergic augmentation

*This section supports the Discussion claim that no serotonin-raising intervention can remit severe OCD without approaching serotonin toxicity. There is no sharply-defined fold-onset of serotonin toxicity; we use a conservative, illustrative supraphysiological band ($\gtrsim 10\times$ baseline) as the toxicity-risk zone, grounded in the serotonin-syndrome literature (Nisijima et al.[68]). The uptake-2/OCT3 pharmacology is the verified material of S2.*

#### S13.1 The plateau as a toxicity-bounded ceiling, and the OCT3 prediction

The calibration attributes the SSRI plateau not to an unresponsive circuit but to a ceiling on how far transporter blockade can raise extracellular serotonin. Treating $e_C$ as a free parameter makes this explicit (Fig. S11): the $e_C$ needed to fold a patient's diseased attractor rises steeply and super-linearly with severity: from $\approx 6\times$ baseline at $Y_{\text{model}} = 24$ to $\approx 12\times$ at 28 and $\approx 25\times$ at 32 (these fold-multiples are illustrative: they extrapolate beyond the data, which reach at most $\approx 5.8\times$ baseline, and they inherit a region- and receptor-uniform proportional-elevation assumption, so only the qualitative ceiling, not the exact multiples, is a model claim; S4), while the excursion is bounded by toxicity, serotonin syndrome developing many-fold above baseline into the supraphysiological range (Nisijima et al.[68]). Because the fold-multiple and the toxicity threshold both scale with the same proportional-elevation mapping, the load-bearing claim is not either absolute multiple but their *ordering* (that folding a severe attractor demands an $e_C$ excursion into the toxicity range) which is more robust to the shared assumption than the individual multiples. For OCD at Y-BOCS $\gtrsim 28$ the remission threshold is squarely supraphysiological, so raising $e_C$ further (a higher dose, uptake-2/OCT3 blockade (Horton et al.[16]), or MAO inhibition) buys less benefit at greater risk. The ceiling nonetheless yields the framework's one novel, committed *serotonergic* prediction: because uptake-2/OCT3 is an SSRI-insensitive clearance floor, blocking it raises $e_C$ where a saturated transporter cannot, augmenting mild-to-moderate patients whose SERT is already occupied yet whose fold is still reachable (untested in humans; bounded by the toxicity ceiling for the severe; S2). If the serotonin *level* is the wrong target for those patients, the right one is the plasticity balance (main text).

**The remission threshold in extracellular serotonin.** Treated as a free parameter (bypassing the balance equation), the extracellular-serotonin level $e_C$ at which the diseased attractor of a patient presenting at baseline $Y_0$ is annihilated rises steeply and super-linearly with severity:

| $Y_0$ | 24 | 26 | 28 | 30 | 32 |
|---|---|---|---|---|---|
| $e_C^{\text{fold}}/e_C^h$ | $6.4\times$ | $8.8\times$ | $12.3\times$ | $17.3\times$ | $25.1\times$ |

(These are the same $e_C^{\text{fold}}$ multiples reported in main-text Fig. 5 and its table.) A maximally-dosed SSRI reaches only $\approx 5.6\times$ baseline (occupancy $u = 0.95$), folding the attractors of the mildest patients (up to $Y_0 \approx 23$); beyond that, further remission requires raising $e_C$ by other means.

**Serotonin toxicity as the ceiling (fold-change).** Because the model's clearance scale is arbitrary ($e_C^h = 2.8$ nM is a model unit, not the microdialysis absolute, and the dynamics are invariant under a common rescaling of release and clearance) the toxicity comparison is made in

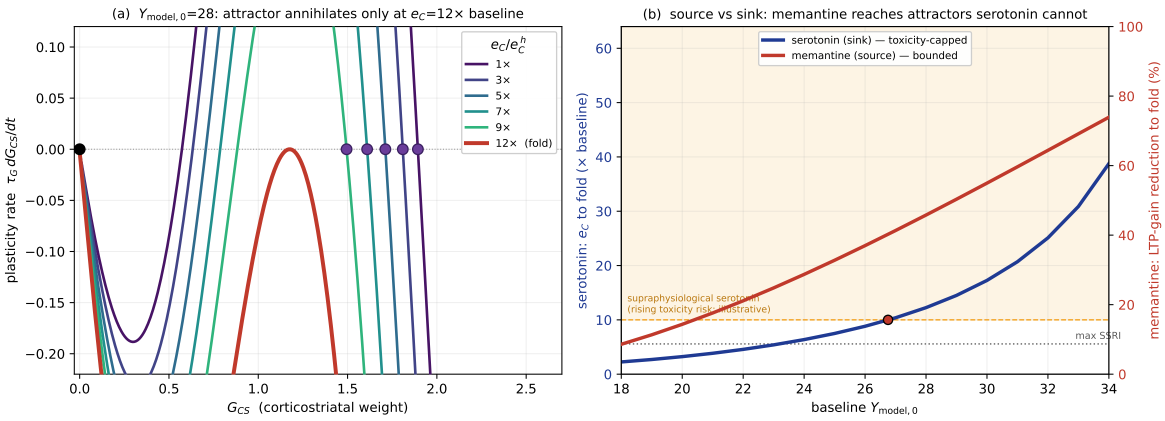

Figure S11: **A toxicity-bounded serotonin ceiling.** *(a)* With $e_C$ a free parameter, raising it steepens the plasticity sink for a $Y_{\text{model}} = 28$ patient; the diseased attractor (purple) slides down but annihilates only at $e_C \approx 12\times$ baseline (red). *(b)* **Source vs sink.** The serotonin $e_C$ needed to fold the attractor (blue, left axis) rises steeply with severity, entering the supraphysiological range ($\gtrsim 10\times$ baseline, shaded) around $Y_{\text{model},0} \approx 27$ (maximal SSRI reaches only $\approx 5.6\times$); the memantine LTP-gain reduction that folds the *same* attractor (red, right axis) stays a bounded, serotonin-independent 9–74% across $Y_{\text{model},0} = 18$–34 (the memantine route; S14). The shaded zone marks supraphysiological serotonin carrying rising serotonergic-toxicity risk (illustrative) and applies to the serotonin route only.

fold-change, not absolute concentration. In animal models, serotonin syndrome (hyperthermia that can be lethal) develops when hypothalamic extracellular serotonin rises many-fold above baseline. In serotonin syndrome the severity is graded and strongly drug-dependent, with no sharply-defined fold-onset of toxicity (Nisijima et al.[68]). We therefore do *not* assert a precise toxicity threshold; we adopt a conservative, illustrative supraphysiological band beginning at $\sim 10\times$ baseline as the zone of rising serotonergic-toxicity risk. The remission threshold $e_C^{\text{fold}}(Y_0)$ enters this band at $Y_0 \approx 27$–28: remitting OCD at or above this severity by serotonin elevation alone requires driving $e_C$ to an order of magnitude or more above baseline, into the range where serotonergic toxicity becomes a material risk.

**OCT3 (uptake-2) augmentation is one instance, and is graded rather than curative.** Blocking the SSRI-insensitive uptake-2/OCT3 clearance floor (S2) raises $e_C$ past the SERT ceiling and is the mechanistically "correct-axis" serotonergic augmenter. Modeled as a fractional reduction of $k_{\text{rem}}$, it does not annihilate the severe attractor at physiological $e_C$; it slides it down, lifting drug-only non-response toward the response threshold (up to $\approx 29\%$) but not to remission (Table S5). At $u = 0.9$ an 80% OCT3 block already drives $e_C$ to $\approx 9\times$ baseline (just below the illustrative supraphysiological band) so any illustrative use is capped at 50% ($\leq 6\times$).

**Available OCT3 blockers are tool compounds, not therapeutics.** The established uptake-2/OCT3 inhibitors are non-selective and unsuitable as therapeutics: decynium-22 (the most potent blocker) inhibits OCT1/2/3 and PMAT with similar potency and remains a pharmacological tool for which selective ligands are still needed (Daws[19]); corticosterone and other glucocorticoids block OCT3 (Gründemann et al.[69]) but carry an untenable endocrine burden for chronic use. The

Table S5: Drug-attributable Y-BOCS drop at 24 weeks for the two treatment-resistant baselines under SSRI + a hypothetical OCT3 blockade, with (% of baseline, extracellular-serotonin multiple reached). OCT3 lifts response but does not remit; the 80%/$u$=0.9 cell sits just below the illustrative $\sim 10\times$ band ($\approx 9\times$).

| $Y_0$ | $u$ | SSRI only | +50% OCT3 | +80% OCT3 |
|---|---|---|---|---|
| 28 | 0.8 | 1.9 (7%, 3.0×) | 2.7 (10%, 3.9×) | 3.5 (12%, 4.7×) |
| 28 | 0.9 | 3.1 (11%, 4.3×) | 5.2 (18%, 6.3×) | 8.2 (29%, 9.0×) |
| 32 | 0.8 | 1.1 (4%, 3.0×) | 1.6 (5%, 3.9×) | 2.1 (7%, 4.7×) |
| 32 | 0.9 | 1.9 (6%, 4.3×) | 3.0 (10%, 6.3×) | 4.6 (14%, 9.0×) |

preclinical proof-of-concept (a sub-effective SSRI + decynium-22 giving a near-maximal serotonin rise) is established (Horton et al.[16]; S2), but no OCT3-targeting augmentation trial has been conducted in humans. The model's contribution is thus a *bound*, not a recommendation: it identifies why serotonergic augmentation (OCT3 included) cannot rescue severe OCD within a tolerable window, motivating the non-serotonergic strategies taken up in the Discussion.

### S14 Memantine and glutamatergic augmentation

#### S14.1 Mechanism: the source route

The corticostriatal-plasticity equation is

$$\tau_G\,\dot{G}_{CS} \;=\; \underbrace{\alpha\,\phi_e\,(\phi_{d_1}-\phi^h_{d_1})(\phi_{d_1}-\theta_M)(\phi_{\mathrm{sat}}-\phi_{d_1})}_{\text{NMDA-dependent BCM } \textit{source} \text{ (LTP)}} \;-\; \underbrace{(1+b\,(e_C-e^h_C))\,G_{CS}}_{\textit{sink} \text{ (SSRI acts here)}}.$$

The SSRI folds the diseased attractor by steepening the sink ($\propto e_C$); memantine folds it by shrinking the source. NMDA receptors are the substrate of this source: they set both the modification threshold $\theta_M$: the NMDA-dependent BCM metaplasticity threshold (Bienenstock et al.[28]; Kirkwood et al.[34]; Cooper & Bear[35]; Yashiro & Philpot[36]), and the *magnitude* of potentiation, through the $Ca^{2+}$ influx they gate (Shouval et al.[29]). A proof-of-concept shows the threshold is the wrong handle: at a deep attractor $\phi_{d_1} \gg \theta_M$, so raising $\theta_M$ barely changes $(\phi_{d_1}-\theta_M)$. The operative lever is the *gain* $\alpha$. Memantine therefore enters as a reduction of the LTP gain,

$$\alpha \;\to\; \alpha\,(1-m), \qquad m \in [0,1],$$

with $m$ the drug's fractional suppression of NMDA-gated potentiation. Although $m$ scales $\alpha$ uniformly, its *effect* is state-dependent, because the LTP source it multiplies itself vanishes at health ($\phi_{d_1} \to \phi^h_{d_1}$): the same $m$ removes a large absolute plasticity current at the over-potentiated OCD attractor and essentially none at the healthy fixed point. This state-dependence follows from the pharmacology: memantine is a pathologically-activated, uncompetitive open-channel blocker that engages excessive NMDA activity and spares normal transmission (Lipton[70]), and it is observed in the right region: memantine shifts *striatal* plasticity toward long-term depression (Mancini et al.[71]).

**Onset.** Memantine's channel block is fast, but its plasticity effect builds over weeks via NMDA-subunit remodeling: chronic memantine drives GluN2B↓ at 14 d and GluN2A/PSD-95↑ at 28 d (Wang et al.[72]; a stroke model, timescale transferable in principle, not OCD-specific). A ∼3-week onset ramp then reproduces the delayed, accelerating clinical response (below).

#### S14.2 Source vs sink: memantine reaches attractors serotonin cannot

Comparing the two fold routes for the *same* attractor across presenting severity (Fig. S11b): folding via the sink requires extracellular serotonin of 12× ($Y_{\mathrm{model},0}$=28) to 39× ($Y_{\mathrm{model},0}$=34) baseline, squarely in the supraphysiological, toxicity-risk range (S13), whereas folding the same attractor via the source requires a 46–74% reduction of the LTP gain (over the same $Y_{\mathrm{model},0}$=28–34): large, but bounded, and serotonin-independent. *The source route is accessible exactly where the sink route is toxic.* For the deepest attractors ($Y_{\mathrm{model},0} \gtrsim 34$) even the source route approaches total LTP loss, consistent with memantine only *partially* lowering the most severe patients (Modarresi et al.[73]: 34 → 20, not to remission).

#### S14.3 Clinical evidence (contested)

Five randomised trials and a critical appraisal (Table S6). The pattern is population-specific, not uniformly positive.

Table S6: Memantine-augmentation trials in OCD. The memantine effect appears where the comparator (SSRI) arm does *not* improve, and is null where the SSRI arm itself folds the attractor.

| Trial | Population | Comparator arm | Memantine effect |
|---|---|---|---|
| Modarresi et al.[73] | severe *refractory* (mean 33.7) | flat (33.5 → 33.5) | large (33.9 → 20.0) |
| Ghaleiha et al.[74] | mod.–severe, non-refractory | partial, 8 wk (32% resp.) | large (89% rem.)† |
| Askari et al.[75] | mod.–severe, non-refractory | folds (30 → 9.5) | null (exec. fn. only) |
| Mirzazadeh et al.[76] | mod.–severe, non-refractory | folds (32.8 → 5.3) | null (exec. fn. only) |
| Farnia et al.[77] | mild–moderate, $n$=99 (largest) | improves | null |

† completer analysis; the pooled meta-analysis (Modarresi et al.[78]) is critiqued by Andrade[79] (single-region trials, completer analyses, publication bias, Farnia excluded): verdict: "unestablished."

#### S14.4 The reconciliation, and a two-axis refinement

Read through the model, most of these trials fall into order: memantine helps where the SSRI *cannot fold* the attractor (refractory, comparator flat: Modarresi et al.[73]) and is redundant where the SSRI *does* fold it (non-refractory with a responding comparator: Askari et al.[75], Mirzazadeh et al.[76], Farnia et al.[77]). The one row that does not fit cleanly is Ghaleiha et al.[74]: a nominally *non*-refractory sample with a large memantine benefit (89% remission, completer analysis). We flag it as a partial counterexample rather than fold it into the pattern. The mitigating detail is a matter of horizon: at the eight-week endpoint its comparator (fluvoxamine) arm had reached only ∼32% response (the expected partial response for an SSRI at that short a timescale, which the trial itself describes as "in the expected range for SRIs," not an anomalously low one) so most of these patients had *not yet* folded despite their non-refractory *history*. On the model's own *folding*-by-response criterion (as opposed to a history-based one) a memantine benefit is therefore expected there. Ghaleiha et al.[74] thus expose a distinction the trial labels obscure: refractoriness-by-history and non-folding-by-response need not coincide, and it is the latter the model speaks to. With that caveat, *memantine benefit tracks whether the SSRI folds the attractor, not baseline severity*, a falsifiable, folding-stratified prediction. This forces a refinement of the main text: fold-reachability has two axes, the attractor depth $\alpha$ (baseline severity) *and* the sink-coupling $b$ (SSRI/SERT efficacy = refractoriness). A treatment-naïve severe patient (Mirzazadeh et al.[76], $Y_{\text{model},0} \approx 32$) folds on the SSRI (normal $b$); a refractory severe patient (Modarresi et al.[73], $\approx 34$) does not (weak $b$), same $Y_0$, opposite outcome. R2–R4 use baseline severity as the fold-reachability proxy; the refinement is that $b$ is a second, orthogonal axis, and memantine (acting on the source) bypasses both.

#### S14.5 An illustrative dose-to-engagement map

The mechanism above treats memantine's action *directionally*: the fractional gain suppression $m$ is a mechanism, not a dose-indexed quantity. Unlike the SSRI (SERT occupancy from [$^{11}$C]DASB, S2) and the antipsychotics ($D_2$ occupancy from PET, S15), memantine has no in vivo receptor-occupancy assay. A dose-to-*engagement* map can nonetheless be built from binding affinity and pharmacokinetics, and it bounds $m$; we give it here as illustrative, because the step from channel engagement to LTP-gain suppression is uncalibrated.

Memantine is a moderate-affinity, uncompetitive, voltage- and use-dependent open-channel NMDA blocker: 6 $\mu$M blocks ∼2/3 of the NMDA current at −70 mV, comparable to MK-801 (Bormann[80]), and the block requires prior receptor activation, sparing phasic transmission (Chen & Lipton[81]; the "too little / too much" homeostatic thesis of Parsons et al.[82]). Its binding affinity at the receptor's PCP site is $K_i \approx 0.5$ $\mu$M in human frontal cortex (Kornhuber & Quack[83]). Clinical doses reach concentrations of the same order: across 5–30 mg day$^{-1}$ human serum memantine is 0.03–0.53 $\mu$M ($\approx$ 0.37 at 20 mg, 0.53 at 30 mg) and free cerebrospinal-fluid concentration is about half that (CSF/serum $\approx$ 0.52; $\approx$ 0.21 $\mu$M at 20 mg), with rat plasma $\approx$ 1 $\mu$M at behaviorally active doses (Kornhuber & Quack[83]; concentrations not fully at steady state). A Hill-1 channel block $\beta_{\text{mem}}(D) = C(D)/(C(D) + K_i)$ against the free (CSF) concentration then gives the memantine analogue of SERT occupancy:

| dose (mg day$^{-1}$) | free (CSF) conc. $C$ ($\mu$M) | NMDA block $\beta_{\text{mem}}$ |
|---|---|---|
| 5 | 0.06 | 0.11 |
| 10 | 0.12 | 0.19 |
| 20 | 0.21 | 0.30 |
| 30 | 0.31 | 0.38 |

At clinical doses this is a $\sim$30–40% block of *resting* receptors, rising beyond half-block for the tonically over-activated receptors the voltage- and use-dependence selects (serum-based concentrations give $\beta_{\text{mem}} \approx$ 0.43–0.51 at 20–30 mg). Memantine's clinical concentration is therefore of the same order as its NMDA affinity: a substantial, preferentially pathological block, not a marginal one.

The remaining step is uncalibrated. What a dose *engages* ($\beta_{\text{mem}}$) is not identical to what it *does* to the LTP gain ($m$): the gain suppression follows from reduced NMDA-gated $Ca^{2+}$ influx, related to but not equal to fractional current block. Identifying $m \approx \beta_{\text{mem}}$ to leading order, clinical doses give $m \approx$ 0.3–0.4 at rest and more at the pathological attractor, of the order needed to fold moderate attractors (S14.2: $m \approx 0.46$ at $Y_{\text{model},0}$=28) but short of the deepest. No dataset fixes the $\beta_{\text{mem}} \to m$ relation (the analogue of the DASB series for SERT or the PET series for $D_2$), which is why the fold-route magnitudes stay illustrative. The map's value is a bound: because $\beta_{\text{mem}} \lesssim 0.4$ clinically, folding an attractor that needs $m \gtrsim 0.5$ requires the block-to-gain coupling to be near-complete or supra-linear, a demanding and testable condition consistent with memantine benefit being confined to the refractory tail.

#### S14.6 Limitations

(i) The positive evidence is weak and contested (Andrade[79]). We present memantine as mechanism + prediction and do *not* calibrate the model to these trials. (ii) A quantitative proof-of-concept shows the model's severe attractors are over-robust (the deepest need near-total LTP loss to fold), so the fold-route magnitudes are directional, and Modarresi et al.'s[73] exact trajectory is reproduced only illustratively (with the subunit-remodeling onset ramp), not fitted. A dose-to-engagement map exists (S14.5), but unlike the antipsychotic (which carries a full dose$\to D_2$-occupancy$\to$fold bridge, S15) the further step from NMDA engagement to LTP-gain suppression is uncalibrated, so every gain-reduction magnitude here (e.g. the 46% of the combination figure) is illustrative: only the mechanism and the refractoriness-tracking *direction* are model claims. (iii) Pasquini's contrasting refractory cases[84] suggest symptom *subtype* (washing vs. checking) is a third heterogeneity axis the model does not capture. (iv) The onset-timescale evidence (Wang et al.[72]) is from a stroke model, transferable only in principle.

#### S14.7 The glutamatergic-augmentation class

Memantine is one of a class of off-serotonin glutamate modulators used as SSRI-refractory augmenters (riluzole[85], ketamine[86,87], N-acetylcysteine[88], amantadine[89], and (augmenting exposure therapy rather than an SSRI) D-cycloserine[90]). In the model each maps to source-shrinking ($\alpha\downarrow$) and, for release modulators, a change in the excitatory drive, a unifying account of why glutamatergic agents cluster as augmenters.

### S15 Antipsychotic augmentation: from $\mathbf{D_2}$ occupancy to the fold

Antipsychotics enter the model not through the plasticity equation (as the SSRI and memantine do) but through the *circuit.* $D_2$ blockade disinhibits the indirect-pathway $D_2$-MSNs; in the van Albada–Robinson mean-field this is a drive $a_{\mathrm{AP}}$ added to the $d_2$ population. Through the closed cortico-striato-pallido-thalamo-cortical loop this lowers cortical and caudate-$D_1$ activity ($\phi_{d_1}$ falls from 48 to 25 at $G_{\mathrm{CS}} = 1.5$ as $a_{\mathrm{AP}} : 0 \to 15$), which enters the plasticity *source* $S(G_{\mathrm{CS}}) = \phi_e(\phi_{d_1} - \phi^h_{d_1})(\phi_{d_1} - \theta_M)(\phi_{\mathrm{sat}} - \phi_{d_1})$ and folds the diseased attractor. This supplement (i) specifies the clinical $\to a_{\mathrm{AP}}$ pharmacology and (ii) shows the fold model reproduces the Bloch et al.[91] augmentation meta-analysis.

#### S15.1 Pharmacology (portable): dose $\to$ $\mathbf{D_2}$ occupancy $\to$ effective blockade

The clinical-dose $\to$ receptor pharmacology is drug chemistry and is model-independent; it is carried over unchanged from the Paper 2 AP module (validated against PET data). Steady-state striatal $D_2$ occupancy follows a Hill function calibrated to PET (haloperidol: Kapur et al.[92]; risperidone: Nyberg et al.[93]; aripiprazole: Mamo et al.[94]). Partial agonism is corrected by the Black–Leff operational model (Black & Leff[95]): a partial agonist at full occupancy still elicits a fraction $\tau/(1+\tau)$ of the maximal response, so the *effective blockade* is

$$\text{eff. blockade} \;=\; \frac{f_{D_2}}{1+\tau}, \qquad \tau_{\text{full antagonist}} = 0 \;\; \text{(ris, hal)}, \quad \tau_{\text{aripiprazole}} = 0.40.$$

Table S7: Antipsychotic PK at representative augmentation doses. $f_{D_2}$ from PET-calibrated Hill functions; effective blockade from the Black–Leff correction. The final column, $\alpha_{AP} = 0.222\times$eff. blockade, is the *prior-model* (Paper 2) circuit coupling and is shown for provenance only: it is **not** used in the present model (see S15.2).

| Drug | Dose | $f_{D_2}$ (%) | $\tau$ | Eff. block. (%) | $\alpha_{AP}$ (prior model) |
|---|---|---|---|---|---|
| Risperidone | 2.0 mg | 54 | 0.00 | 54 | 0.121 |
| Aripiprazole | 10.0 mg | 90 | 0.40 | 64 | 0.142 |
| Haloperidol | 3.0 mg | 76 | 0.00 | 76 | 0.169 |

#### S15.2 Bridge to the current model: $F_{\mathrm{OCC}}$ is *not* reused

The pharmacology stops at effective blockade; converting that to a circuit effect requires one model-specific constant. In Paper 2 that constant was $F_{\mathrm{OCC}} = 0.222$, defined by $\alpha_{AP} = F_{\mathrm{OCC}} \times$eff. blockade, where $\alpha_{AP}$ *reduced a cortico-$D_2$ synaptic weight* ($w_{CD_2} \to w_{CD_2}(1 - \alpha_{AP})$) in the earlier limit-cycle model. The present model does not have that weight: it injects $D_2$-MSN *drive* $a_{\mathrm{AP}}$, with different units. $F_{\mathrm{OCC}}$ therefore does not transfer, and we do not reuse its value. We instead recalibrate a single fresh bridge constant $C$,

$$a_{\mathrm{AP}} \;=\; C \times \text{eff. blockade},$$

fixed by *one* anchor (the Bloch pooled-AP response) to $C = 8$. Everything else below (placebo arm, drug ordering, the occupancy fold-window) is then a consequence, not a fit.

##### S15.3 The Bloch et al. comparison

Bloch et al.[91] pooled 9 double-blind RCTs (278 participants) of antipsychotic augmentation in OCD patients on a stable, inadequately-responding SRI (the defining feature of the trial population). We represent this population as refractory (the stable SRI is present but does *not* fold the attractor, so we set $e_C = e_C^h$ for the backbone) with baseline severity $Y_{\text{model},0} \sim \mathcal{N}(26, 5)$. Response is attractor annihilation (remission). One constant ($C$) is calibrated to the pooled arm; the rest are interpolations from that single anchor.

Table S8: Bloch et al.[91] augmentation meta-analysis vs. the fold model. Response = ≥35% Y-BOCS reduction (Bloch) / attractor annihilation (model). Only the pooled-AP row is a calibration target.

| Arm | Bloch et al.[91] | Model | Note |
|---|---|---|---|
| SSRI + placebo (refractory) | 11% | ∼0–4% | backbone $e_C = e_C^h$ (refractory) |
| SSRI + AP (pooled) | 32% | 36% | *calibration anchor* ($C = 8$) |
| + risperidone 2 mg | 50% | 39% | interpolation (eff. block. 54%) |
| + haloperidol 3 mg | 50% | 52% | interpolation (eff. block. 76%) |

The refractory backbone gives a low placebo response (∼0–4%), of the same order as (though somewhat below) Bloch's 11% (both far under the ∼30% placebo response of non-refractory OCD trials), so the model *under*-predicts the refractory placebo arm; only the pooled-AP anchor is a calibration target, and stronger $D_2$ blockade yields higher interpolated response (haloperidol > risperidone at these doses), consistent with the strong-AP arms of Bloch (risperidone/haloperidol both 50%). Risperidone augmentation spans 0.5–3 mg clinically; at 2 mg the model gives 39%, rising with dose.

##### S15.4 An emergent occupancy fold-window (model-derived, contingent on $C$)

Mapping the $a_{\text{AP}}$ that folds each attractor back through the bridge into $D_2$ occupancy gives a clinically-legible model output (Table S9). A patient with $Y_{\text{model},0} \leq 26$ can be folded at $\lesssim 73\%$ striatal $D_2$ occupancy (*consistent with* Kapur's 65–72% therapeutic window; with a single calibrated bridge constant this is a consistency check, not an out-of-sample prediction) whereas $Y_{\text{model},0} \geq 28$ requires supra-physiological occupancy (> 100%), i.e. cannot be folded by $D_2$ blockade at all. This is the mechanism behind the ∼1/3 ceiling of AP augmentation: the reachable population is exactly the milder tail, and the refractory-severe core is beyond $D_2$'s reach, the same fold-unreachability that motivates the off-dopamine (source) agents.

Table S9: Occupancy needed to fold the attractor by severity (full-antagonist, $\tau = 0$, so eff. block. = occupancy). Values above 100% are unreachable: $D_2$ blockade cannot fold these attractors.

| $Y_{\text{model},0}$ | 24 | 26 | 28 | 30 |
|---|---|---|---|---|
| $a_{\text{AP}}$ to fold | 4.2 | 5.9 | 8.7 | 13.1 |
| $D_2$ occupancy to fold | 53% | 73% | *>100%* | *>100%* |

##### S15.5 Limitations

(i) The bridge constant $C$ is calibrated to a single pooled meta-analytic number; the drug-specific rows are interpolations that rest on that one anchor, not independent predictions. (ii) The refractory population is modeled by $e_C = e_C^h$ and an assumed $\mathcal{N}(26, 5)$ baseline (Bloch does not report a per-patient baseline distribution); the placebo agreement is therefore a consistency check, not an independent validation. (iii) AP response here is attractor annihilation only; the acute readout effect ($\phi_{d_1}\downarrow \Rightarrow Y\downarrow$ immediately) would push additional non-folded patients over the 35% threshold, so the model response is a lower bound. (iv) Side-effect liabilities (EPS, the aripiprazole/risperidone difference) are deliberately excluded: see the main-text inclusion rule; the Black–Leff $\tau$ enters only through efficacy (effective blockade), not through a side-effect prediction.

### S16 Ondansetron and further extensions of the framework

#### S16.1 Ondansetron (5-$HT_3$ receptor antagonism)

Ondansetron is the sharpest test of the serotonin-ceiling thesis: serotonergic in name, it helps *without* raising extracellular serotonin. Blocking the 5-$HT_3$ channel reduces excitatory and dopaminergic drive into the corticostriatal loop (Hamanaka et al.[96]; dose-dependent reduction of cortical activation, Stern et al.[97,98]), lowering $\phi_e$ and hence $\phi_{d_1}$ and so reaching the fold by the same circuit route as the antipsychotic (lowering the plasticity source's circuit argument) but without $D_2$ blockade or its extrapyramidal and metabolic burden. Which circuit node the 5-$HT_3$ block acts through is not pinned by the present data, but the framework can examine each candidate route: any intervention that lowers $\phi_{d_1}$ shrinks the source and folds the attractor, so the qualitative prediction is route-independent. Adjunctive 5-$HT_3$ antagonists reduce Y-BOCS in controlled trials (pooled $-5.1$ points; Hamanaka et al.[96]), and worsening on discontinuation (Pallanti et al.[99]) is the signature of a re-emerging attractor. With no occupancy model and a flat dose–response, we place ondansetron in the framework qualitatively.

#### S16.2 Extending the framework

The same landscape supports two extensions. *Non-pharmacological therapies* enter the circuit off the serotonin axis: rTMS modulates the cortical drive into the loop, and exposure and response prevention is extinction learning that lowers $G_{\mathrm{CS}}$ directly. Because rTMS acts from the cortical side and ERP through the amygdala-to-cortex drive, both shrinking $G_{\mathrm{CS}}$ through the shared orbitofrontal node, the framework predicts their combination to be super-additive, testable in combined rTMS-plus-ERP protocols such as those of Fitzsimmons et al.[100] and Postma et al.[101]. *Comorbid depression and anxiety* share the serotonergic engine and overlapping circuitry; coupling the circuit to limbic and mood loops would let the *same* calibrated engine drive two bistable circuits at once, a natural account of why one drug moves depressive and obsessive-compulsive symptoms on different timescales.

### S17 Combination therapy: complementary routes to the fold

#### S17.1 Three mechanisms, three different terms

The augmentation strategies of the Discussion reach the *same* saddle-node fold by acting on *different* terms of the corticostriatal-plasticity equation

$$\tau_G \dot{G_{\rm CS}} = \underbrace{\alpha\,\phi_e\,(\phi_{d_1} - \phi_{d_1}^h)(\phi_{d_1} - \theta_M)(\phi_{\rm sat} - \phi_{d_1})}_{\text{LTP source}} - \underbrace{(1 + b\,(e_C - e_C^h))\,G_{\rm CS}}_{\text{sink}}.$$

The SSRI steepens the *sink* through $b\,e_C$; memantine shrinks the *source gain* $\alpha$ (S14); the antipsychotic lowers the *source argument* $\phi_{d_1}$ through $D_2$-mediated circuit feedback (S15); ondansetron acts on the same circuit balance from the 5-$HT_3$ side (Discussion, §4.3). Because these levers are largely separable, their effects on the fold approximately stack: interventions each individually sub-threshold can jointly annihilate an attractor that none folds alone. This is the mechanistic rationale for the sequential and combination augmentation that is already standard (if off-label) practice in treatment-resistant OCD (antipsychotic augmentation: Veale et al.[102]; glutamatergic/memantine augmentation, well-tolerated in treatment-resistant OCD: Aboujaoude et al.[103]).

#### S17.2 Each mechanism alone is sub-threshold

For the refractory patient of the main-text combination figure ($Y_{\rm model,0} = 32$), none of the three interventions at the illustrative levels folds the attractor on its own (Fig. S12): the SSRI at SERT occupancy $u = 0.8$ (a $\sim 3\times$ elevation of $e_C$, far below the $\sim 25\times$ the sink route would need at this severity, S13), the antipsychotic at its $D_{50}$ dose, and a 46% memantine LTP-gain reduction each leave a diseased fixed point standing.

#### S17.3 Quantitative complementarity

The stacking is quantitative: the memantine LTP-gain reduction required to fold this $Y_{\rm model,0} = 32$ attractor falls as the other mechanisms are added (Table S10). Memantine alone needs a 64% reduction; with the SSRI on board, 58%; with both the SSRI and the antipsychotic, only 43%, so the antipsychotic is exactly what lets an illustrative 46% memantine tip the attractor over (the SSRI+memantine two-drug path, without the antipsychotic, still leaves it standing). Each mechanism lowers the intensity the others must supply.

Table S10: Memantine LTP-gain reduction required to fold the $Y_{\rm model,0} = 32$ attractor, as complementary mechanisms are added. SSRI at $u = 0.8$; antipsychotic at $D_{50}$. Canonical sink coupling $b = 0.0268$ nM$^{-1}$.

| Background on board | Memantine reduction to fold |
|---|---|
| none (memantine alone) | 64% |
| + SSRI ($u = 0.8$) | 58% |
| + SSRI + antipsychotic ($D_{50}$) | 43% |

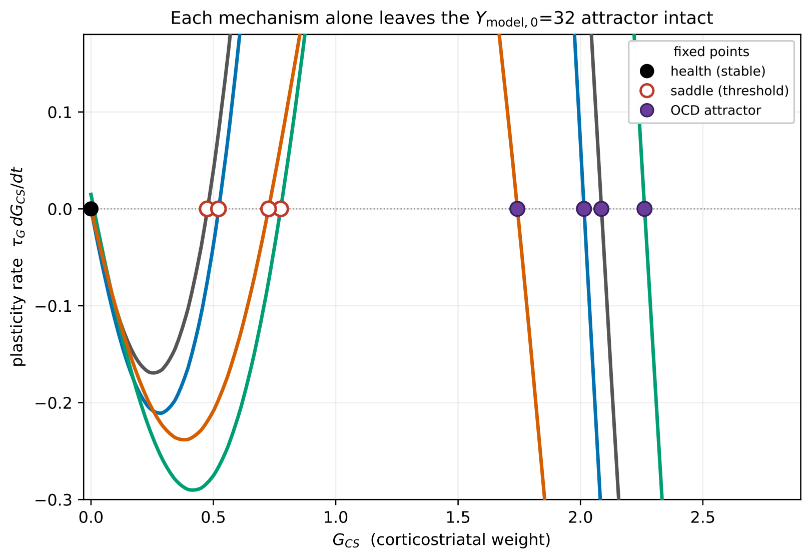

Figure S12: **Each mechanism alone leaves the $Y_{\text{model},0} = 32$ attractor intact.** Plasticity rate $dG_{\text{CS}}/dt$ vs. $G_{\text{CS}}$; a filled circle on the zero line marks a diseased attractor. The untreated case (gray) and each monotherapy at the illustrative level, SSRI at $u = 0.8$ (blue), antipsychotic at $D_{50}$ (green), memantine at 46% LTP-gain reduction (vermillion), all retain the elevated attractor. Only the combination of all three removes it (main-text combination figure).

#### S17.4 Limitations

This is a proof of principle of mechanistic complementarity, not a dosing rule. (i) The model treats the three plasticity terms as separable and additive; it does not capture pharmacokinetic interactions, shared side-effect burden, or adherence, all of which bound combination therapy in practice. (ii) The illustrative levels ($u = 0.8$, $D_{50}$, 46%) are chosen to demonstrate sub-threshold stacking at a single presenting severity; the memantine LTP-gain reduction is not calibrated to a dose (S14). (iii) The demonstration uses the uniform gain reduction $\alpha \to \alpha(1 - m)$ of S14; because the LTP source vanishes at health, this already concentrates memantine's effect at the over-potentiated attractor.

#### S18 The fast serotonergic cortical channel $\gamma_C$ is not identifiable

The circuit admits, *a priori*, a direct serotonergic drive $\gamma_C\, e_C$ onto the cortical excitatory population (main-text Eq. (3)): because serotonin can modulate cortical pyramidal excitability, a fast action of the drug on the circuit is biologically plausible. We test whether the trajectories support a nonzero $\gamma_C$ by profiling it: refitting the five structural constants together with $\gamma_C$ and comparing the pooled discrepancy to the $\gamma_C = 0$ baseline.

The data do not support a fast channel. At the adopted $\gamma = 2$, the free-$\gamma_C$ optimum sits at $\gamma_C = 1.3 \times 10^{-4}$ with pooled discrepancy 6.41 versus 6.40 at $\gamma_C = 0$, a *negative* marginal gain ($-0.2\%$, i.e. no improvement), and the induced cortical shift at maximal drug is $< 0.001$ mV, dynamically negligible. This has a structural cause: at the OCD fixed point the caudate rate $\phi_{d_1}$ is hypersensitive to cortical drive (positive-feedback amplification through the cortico–striato–thalamo–cortical loop, $\partial\phi_{d_1}/\partial V_e \approx 160\ \mathrm{s}^{-1}/\mathrm{mV}$), so $\gamma_C$ is a *near-singular* channel: an infinitesimal coupling produces a large, ill-conditioned response, and the calibration can neither identify nor benefit from a nonzero value. We therefore set $\gamma_C = 0$; the therapeutic effect flows entirely through the slow corticostriatal plasticity of the main text §2, and drug onset is set by the plasticity and autoreceptor timescales rather than by any fast cortical action.

### S19 The expectation (placebo) response

**Summary.** In the frozen model the placebo/expectation response is carried *entirely at the observational layer* as an additive correction $\varepsilon(t)$ to the disease-state readout; it does not touch the corticostriatal weight $G_{\mathrm{CS}}$. It is fit *per trial*, $\varepsilon(t) = \Delta\varepsilon\,(1 - e^{-t/\tau_\varepsilon})$ with $\varepsilon(0) = 0$, and cancels identically in the net (drug−placebo) quantity used for calibration. This supersedes the v1 non-specific-plasticity model (a distributed drift $\kappa_{NS}$ on $G_{\mathrm{CS}}$), which was introduced only to *mechanistically simulate* a trial pre-selection step that the final analysis does not perform.

#### S19.1 Where placebo enters the model

The observed Y-BOCS score is the disease-state readout minus an expectation correction:

$$Y_{\text{obs}}(t) = \underbrace{Y_p\big(G_{\text{CS}}(t)\big)}_{\text{disease state}} - \varepsilon(t), \qquad \varepsilon(t) = \Delta\varepsilon\left(1 - e^{-t/\tau_\varepsilon}\right). \tag{9}$$

The layer at which placebo acts is set by two structural choices:

1. **Observation layer only.** $\varepsilon(t)$ shifts the *measured* score; it does *not* change $G_{\text{CS}}$. The underlying attractor is untouched, so a placebo "responder" in this model has an unchanged disease state.
2. **Zero at enrollment.** $\varepsilon(0) = 0$: there is no expectation before treatment begins, so the observed baseline maps directly to the disease state and the baseline inversion $Y_0 \mapsto G_{\text{CS}}(0)$ carries no placebo offset. This removes the intercept parameter of the earlier form $\varepsilon_0 + \Delta\varepsilon(1 - e^{-t/\tau_\varepsilon})$ (fitted $\varepsilon_0 \approx 0.08$ pt on the Tollefson arm, i.e. negligible); one fewer parameter per trial.

The asymptotic value $\varepsilon(\infty) = \Delta\varepsilon$ is the correction used in the steady-state (attractor) responder criterion; the full $\varepsilon(t)$ is used whenever a time-course is compared to trial timepoints.

#### S19.2 Why an observation-layer correction, not a dynamical placebo

Placebo could instead be modeled dynamically, as a non-specific recovery drift added to the $G_{\text{CS}}$ sink so that the disease state itself relaxes toward health under placebo. Such machinery does real work only when one mechanistically simulates a response-gated pre-selection and propagates each survivor's disease state forward. Our analysis takes any pre-selected cohort *as reported* (§S19.5) rather than simulating the selection, so a dynamical placebo does no work here while adding a distributional parameter that only two placebo arms could constrain. The observation-layer correction keeps the placebo model at the resolution the data support.

#### S19.3 Grounding: placebo as an expectation phenomenon

The additive expectation form is grounded in the neurobiology of placebo, reviewed by Benedetti et al.[104] The real placebo effect is a psychobiological phenomenon driven by expectation, anxiety-reduction, reward, and Pavlovian conditioning, "not a single but many placebo effects, with different mechanisms and in different systems, medical conditions." Two consequences shape our use of $\varepsilon(t)$:

**It is a correction for causes outside the modeled biology.** Following Benedetti et al.'s[104] Figure 1, the improvement seen in a trial's placebo group is the *sum* of spontaneous remission, regression to the mean, measurement biases, unidentified co-interventions, and the expectation effect. Our $\varepsilon(t)$ absorbs all of these; it is explicitly a final correction for improvement not attributable to the drug's action on $G_{\text{CS}}$, not a claim to model the psychology of placebo.

**It is trial-specific, not universal.** Because the placebo response is context-, population-, and conditioning-dependent (Benedetti et al.[104]), $\Delta\varepsilon$ is fitted *per trial*; the trial-to-trial variation we model is that context, population, and conditioning dependence.

##### S19.4 Calibration of $\Delta\varepsilon$

$\Delta\varepsilon$ is fit per trial and $\tau_\varepsilon$ is shared, against the *seven* OCD trials in the calibration panel whose placebo arms are reported (Y-BOCS change from baseline). Three provide full mean time courses that constrain $\tau_\varepsilon$: Stein et al.[105] (their Table 2: ten timepoints with SE), Montgomery et al.[67] (Figure 1 shape with Table 3 endpoint $-5.6$, SD 6.9, $n_{\text{plac}} = 101$), and Tollefson et al.[106] (8-point arm, $24.3 \to 23.6$), and the remainder provide week-12 endpoints: Zohar & Judge[46] ($-5.0$, responders 35.4%), Greist et al.[43] ($-3.4$), Kamijima et al.[107] ($-3.5$), Hollander et al.[108] ($-5.6$). The drug arms of Stein and Zohar enter the drug calibration as *net* (drug$-$placebo) trajectories digitized from the same papers; the two extractions are mutually consistent (e.g. Stein paroxetine week-12 net $3.2 = 11.7 - 8.5$).

A joint least-squares fit of Eq. (9) (shared $\tau_\varepsilon$, per-trial $\Delta\varepsilon$) gives:

| Trial (drug) | $\Delta\varepsilon$ (Y-BOCS pt) | placebo $\Delta Y_{12}$ | responders$_{12}$ |
|---|---|---|---|
| Tollefson et al.[106] (FLX) | 1.7 | 0.7 | 8.5% |
| Greist et al.[43] (SRT) | 3.8 | 3.4 | n/a |
| Kamijima et al.[107] (PAR) | 4.1 | 3.5 | n/a |
| Zohar et al.[46] (PAR) | 5.6 | 5.0 | 35.4% |
| Montgomery et al.[67] (CIT) | 5.9 | 5.6 | 36.6% |
| Hollander et al.[108] (FLV) | 6.3 | 5.6 | n/a |
| Stein et al.[105] (ESC/PAR) | 9.6 | 8.5 | $\sim$50% |
| shared $\tau_\varepsilon$ | 5.5 wk (independent per-arm $\tau$: 5–11 wk) | | |

Table S11: **Per-trial expectation calibration ($\varepsilon_0 = 0$), seven placebo arms.** The asymptotic correction $\Delta\varepsilon$ spans 1.7–9.6 Y-BOCS points (a 6$\times$ range), from the near-flat Tollefson placebo to the very large Stein placebo ($\sim$50% responders). Three arms (Stein, Montgomery, Tollefson) carry the $\tau_\varepsilon$ constraint; a single shared $\tau_\varepsilon \approx 5.5$ wk is an approximation, the per-arm $\tau$ ranges $\sim$5 wk (Stein) to $\sim$11 wk (Montgomery).

The 6$\times$ spread in $\Delta\varepsilon$ across seven trials is the quantitative statement of non-universality: no single $\Delta\varepsilon$ can span the near-flat Tollefson placebo and the very large Stein placebo. This is exactly the context/population dependence Benedetti et al.'s[104] account predicts, and it is why $\Delta\varepsilon$ carries a trial index.

##### S19.5 Use in the responder criterion and in predictions

**Responder criterion (attractor form).** A patient enters at baseline $Y_0$, which inverts (with $\varepsilon(0) = 0$) to the elevated stable fixed point $G_{\text{CS}}(0)$. A drug at SERT occupancy $u = D/(D_{50} + D)$ raises steady-state serotonin, tilts the plasticity right-hand side, and moves the attractor to $G_{\text{CS}}(\infty)$ (or annihilates it at the fold). The predicted observed endpoint is $Y_\infty = Y_p(G_{\text{CS}}(\infty)) - \Delta\varepsilon$, and the patient is a responder if $Y_0 - Y_\infty$ exceeds the threshold (25%/35%). This criterion is $t \to \infty$ and therefore set by the attractor location alone; it gives the achievable ceiling, which for near-fold patients can exceed the 12-week response (critical slowing).

**Hypothetical interventions are reported net.** For a hypothetical agent whose expectation response is unknown (notably a hypothetical OCT3 blocker used to augment SSRI) $\Delta\varepsilon$

cannot be assigned. Such predictions are therefore reported *strictly as net* (drug−placebo, or augmented−unaugmented) improvements, in which $\varepsilon$ cancels identically and no assumption about the expectation response is required.

**Pre-selected cohorts.** A continuation/augmentation cohort of prior SSRI non-responders (e.g. Bloch et al.[91]) has no placebo pre-trial arm from which to calibrate $\Delta\varepsilon$. We set the phase-2 expectation $\varepsilon \approx 0$: by Benedetti et al.'s[104] conditioning principle, a documented prior non-response provides no positive drug-conditioning and blunts the expectation of benefit, and any fast expectation component is exhausted during the lead-in. We treat the phase-2 response as pure fixed-point motion and state this as an assumption; where a phase-2 control arm exists it provides a direct check.

#### S19.6 Relationship to pharmacometric placebo models

The observation-layer correction is the mechanistic-circuit analogue of the disease-progression term in pharmacometric disease-progression+drug-action models (Holford[109]; Friberg et al.[110]; Gomeni & Merlo-Pich[111]), with $\Delta\varepsilon(1 - e^{-t/\tau_\varepsilon})$ the saturating-exponential placebo form. Working from aggregate group means, we fit a per-trial mean $\Delta\varepsilon$ and a shared $\tau_\varepsilon$; joint drug+placebo estimation is respected by the net-cancellation structure, in which the drug parameters are set from trajectories with $\varepsilon$ removed identically.

#### S19.7 Limitations

1. $\varepsilon(t)$ absorbs spontaneous remission, regression to the mean, measurement bias, and unidentified co-interventions in addition to the expectation effect (Benedetti et al.[104]); it is not a pure biological placebo term.
2. Seven placebo arms constrain the calibration (three with full time courses), but a single shared $\tau_\varepsilon \approx 5.5$ wk remains an approximation: independent per-arm $\tau$ ranges $\sim 5$ wk (Stein) to $\sim 11$ wk (Montgomery), so $\tau_\varepsilon$ itself carries trial-to-trial variation the shared value averages over. Individual-patient data would be required to resolve it.
3. The Tollefson placebo arm is noisy and non-monotone; a smooth saturating $\varepsilon(t)$ captures its magnitude but not its week-to-week fluctuations.
4. The phase-2 $\varepsilon \approx 0$ assumption for pre-selected cohorts is grounded in conditioning theory but is not independently calibrated for any specific continuation trial.

### S20 Stability and OCD/PD dynamical separation

**Purpose.** The main text treats OCD as a *stable, elevated fixed point* of the van Albada and Robinson basal-ganglia–thalamocortical mean-field model (Parts I–II), read out through the caudate ($D_1$-MSN) rate $\phi_{d_1}(G_{\mathrm{CS}})$. Two claims must be discharged: (i) that fixed point is stable and non-oscillatory over the clinical range of the corticostriatal-overdrive parameter $G_{\mathrm{CS}}$, so the quasi-static reduction used in the main text is self-consistent; and (ii) this is *not* inconsistent with the fact that the same circuit, under dopamine loss, produces the Parkinsonian theta/beta rhythms of van Albada et al.[112] (their Paper II). We establish both with a delay-resolved linear-stability (dispersion) analysis, first *validated* against van Albada et al.'s[112] own published oscillation thresholds.

#### S20.1 Dispersion relation

Linearizing the full dynamics about a fixed point $\phi_a^0$ and passing to the Fourier variable $s = \sigma + i\omega$ gives the characteristic equation

$$\det\big[\,\mathbf{D}(s) - \mathbf{K}(s)\,\big] = 0, \qquad K_{ab}(s) = G_{ab}\, e^{-s\tau_{ab}}, \quad G_{ab} = \rho_a\, \nu_{ab}, \tag{10}$$

where $\rho_a = \phi_a^0(1 - \phi_a^0/Q_a)/\sigma'$ is the sigmoid slope (gain) of the *receiving* population $a$, $\tau_{ab}$ are the axonal conduction delays, and $\mathbf{D}(s) = \mathrm{diag}\big(D_{\alpha\beta}(s), \dots\big)$ is the dendritic filter $D_{\alpha\beta}(s) = (1 + s/\alpha)(1 + s/\beta)$ on every population, multiplied on the cortical row by the damped-wave factor $(1 + s/\gamma_e)^2$ (spatially uniform mode $k = 0$, the least stable in this corticothalamic model, as finite wavenumbers add spatial damping). Here $\gamma_e = v_e/r_e$ is the cortical damping rate, the temporal decay rate of the corticothalamic damped-wave propagator, set by the axonal conduction velocity $v_e$ and the mean-field axonal range $r_e$ of the excitatory cortical population. We use van Albada and Robinson's healthy parameters ($\alpha = 160$, $\beta = 640$, $\gamma_e = 125$ s$^{-1}$; corticothalamic delays $\tau_{se} = \tau_{re} = 50$, $\tau_{es} = \tau_{is} = 35$ ms; basal-ganglia-internal delays 1–3 ms). The rightmost root $s^*$ of (10) governs stability: $\Re s^* < 0$ is a stable fixed point whose damped resonances sit at $f = \mathrm{Im}\, s^*/2\pi$; $\Re s^* = 0$ with $\mathrm{Im}\, s^* \neq 0$ is a Hopf boundary (self-sustained oscillation).

Van Albada et al.[112] (Paper II) list, for the healthy circuit, the critical gain at which each individual connection destabilizes the fixed point and the frequency of the resulting instability (their Table 2). We validated our numerical encoding of (10) by reproducing these results.

#### S20.2 The Parkinsonian state is a stable fixed point with damped resonances

Van Albada et al.'s Parkinsonian "oscillations" are not a limit cycle our steady-state solver would miss. Their *full Parkinsonian parameter set*, weaker direct and stronger indirect pathways, reduced intrapallidal inhibition, lowered GPe and STN firing thresholds, a stronger striatum→GPe projection, and weaker cortical interactions (their §5.3), yields a *stable* fixed point: the roots of the dispersion relation (10) all lie in the stable half-plane ($\Re s^* < 0$), for the Parkinsonian state exactly as for the healthy one. Van Albada et al. state this explicitly: "each of the states represents a stable system, since all roots are found in the lower half plane" (their Fig. 10b,c). The enhanced Parkinsonian rhythms (elevated relative theta (∼3–7 Hz) and beta (∼15–25 Hz) power) are *noise-driven damped resonances* of this stable fixed point: dopamine loss moves the least-damped roots *closer to, but not across,* the real axis, sharpening those spectral peaks (their Figs. 10–11), and the rhythms are sustained by ongoing brainstem/noise input rather than by an autonomous oscillator.

Limit cycles do exist in the model, but only under *extreme, single-loop gain excursions*: for example an indirect-loop gain far exceeding any corticothalamic gain, giving a ∼5 Hz cycle (their Fig. 8a,

§7.1.2). The authors regard these as "unlikely to occur in the system as a whole," arising if at all only "in subcircuits"; their Table 2 lists the corresponding single-gain instability thresholds, all of which lie *beyond* the physiological Parkinsonian operating point. Our solver therefore returns the full Parkinsonian state as a stable fixed point by construction, in agreement with van Albada et al.[112] The distinction that matters for the present work is *spectral*, Parkinsonian resonances sharpen, whereas the OCD fixed point stays over-damped (S20.3), not a difference between a fixed point and a limit cycle.

##### S20.3 The OCD fixed point stays over-damped for all $G_{\mathrm{CS}}$

Scanning the OCD parameter $G_{\mathrm{CS}}$ from 0 to 3 (well past the clinical range), the rightmost root remains firmly in the left half-plane and pinned near the corticothalamic alpha resonance (∼9 Hz): $\Re s^*$ moves only over $[-14.2, -13.0]$ $\mathrm{s}^{-1}$ and never approaches the imaginary axis (Fig. S13). No delta or theta resonance sharpens; the modest rise in relative low-frequency power (delta 25 → 31%, theta 12 → 16%) is the $1/f$ gain of the spectrum rising as the fixed point translates, not an emerging oscillator. The OCD state is thus a *graded, elevated, over-damped fixed point* for all $G_{\mathrm{CS}}$, and the quasi-static reduction is self-consistent throughout.

##### S20.4 Why PD resonates but OCD does not

A spectral resonance requires a *delayed negative-feedback loop* whose gain is large enough to bring a complex eigenvalue pair close to the imaginary axis; the loop delay sets the frequency, the loop gain its sharpness. The two diseases act on different loops in opposite directions.

*A. Topology (primary).* Dopamine loss, by construction, *raises* the gain of the resonant loops (the indirect pathway (cortex→$D_2$→GPe→STN→GPi→thalamus) and the reciprocal STN–GPe loop) via stronger cortex→$D_2$ and $D_2$ →GPe transmission, weaker intrapallidal inhibition, and lowered STN and GPe thresholds. This pushes those eigenvalue pairs toward the imaginary axis and sharpens the theta/beta resonances. OCD's knob $G_{\mathrm{CS}}$ instead augments the *direct, feed-forward* pathway (cortex→$D_1$→GPi), which is not a resonant loop and touches none of the pallidal changes that boost the indirect loops. Even at OCD's peak striatal loop gain (Table S12), the dominant mode barely moves ($\Re s^* : -13.5 \to -13.0$).

*B. Saturation (secondary).* The gain of any loop through the $D_1$-MSN population is proportional to its sigmoid slope $\rho_{d_1} = \phi_{d_1}(1 - \phi_{d_1}/Q_{d_1})/\sigma'$. As $G_{\mathrm{CS}}$ drives $\phi_{d_1}$ up through the sigmoid, $\rho_{d_1}$ first rises, peaks near $G_{\mathrm{CS}} \approx 1$, then *collapses* $\sim 10\times$ into saturation (Table S12). Severe OCD is therefore *more* over-damped, not less, the opposite of what a developing oscillator would require.

| $G_{\text{CS}}$ | $\phi_{d_1}$ ($\text{s}^{-1}$) | $\rho_{d_1}$ | $D_1$ loop gain $G_{d_1e}$ |
|---|---|---|---|
| 0.0 | 7.4 | 1.72 | 1.72 |
| 1.0 | 33.0 | 4.28 | **8.55 (peak)** |
| 2.0 | 58.0 | 1.64 | 4.93 |
| 3.0 | 64.2 | 0.22 | **0.86 (collapsed)** |

Table S12: **The corticostriatal loop gain is non-monotonic in severity and collapses in severe OCD.** As the overdrive $G_{\text{CS}}$ increases (column 1), the caudate rate $\phi_{d_1}$ rises toward its ceiling $Q_{d_1} = 65\ \text{s}^{-1}$ (column 2). The gain of any loop through the $D_1$-MSN population scales with the sigmoid slope $\rho_{d_1} = \phi_{d_1}(1 - \phi_{d_1}/Q_{d_1})/\sigma'$ (column 3), so the effective cortico-$D_1$ loop gain $G_{d_1e} = \rho_{d_1}(\nu_{d_1e} + G_{\text{CS}})$ (column 4) first *rises*, peaks near $G_{\text{CS}} \approx 1$, then *collapses* nearly ten-fold as $\phi_{d_1}$ saturates toward $Q_{d_1}$. The table reveals that severe OCD is therefore *more* over-damped, not less (the opposite of the gain increase a developing oscillator would require) so corticostriatal overdrive cannot push the fixed point toward an oscillatory instability.

**Conclusion.** The same circuit hosts two dynamically distinct diseases: PD as a gain increase in delayed negative-feedback loops that sharpens noise-driven theta/beta resonances about a still-stable fixed point, and OCD as a feed-forward, $D_1$-saturating corticostriatal overdrive that translates the fixed point to an elevated, over-damped operating point without any oscillatory instability. This dynamical separation matches the clinical phenotypes (PD as a rhythm disorder, OCD as caudate/striatal hyper-activity that scales with severity) and is consistent with the fixed-point framing on which the main text relies.

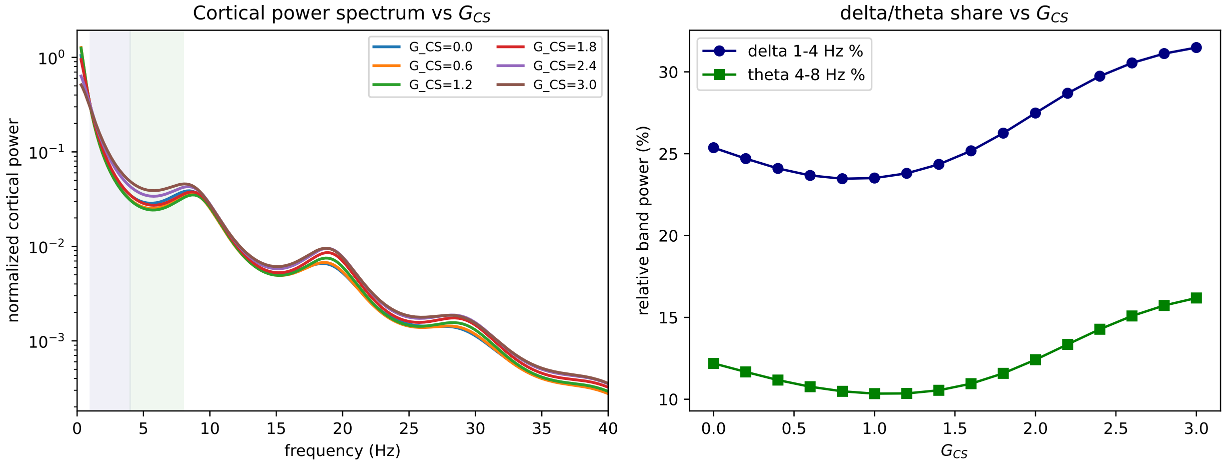

Figure S13: **The OCD fixed point stays over-damped as severity increases; no low-frequency oscillator emerges.** Both panels are computed from the noise-driven linear response of the van Albada and Robinson circuit at the OCD operating point, as the corticostriatal-overdrive parameter $G_{\mathrm{CS}}$ is scanned from 0 (healthy) to 3 (well past the clinical range). The *cortical power spectrum* $P(f)$ is the mean-field EEG proxy, the power spectral density of the cortical excitatory rate $\phi_e$ driven by a spatially-uniform stochastic input at the thalamic relay, $P(f) = |\phi_e(2\pi i f)|^2$, obtained from the linearised dynamics [Eq. (10)] evaluated on the imaginary axis. *Relative band power* in a band is the fraction of total spectral power it contains, $\int_{\mathrm{band}} P\,df \,/ \int_{0.3}^{40\,\mathrm{Hz}} P\,df$ (delta = 1–4 Hz, theta = 4–8 Hz). *Left panel*, each spectrum is normalized to unit total power, so the curves compare spectral *shape* across severities ($G_{\mathrm{CS}} = 0, 0.6, 1.2, 1.8, 2.4, 3.0$; delta/theta bands shaded). It reveals that increasing $G_{\mathrm{CS}}$ leaves the spectral shape essentially unchanged: the dominant $\sim$9 Hz corticothalamic alpha shoulder neither sharpens nor migrates to lower frequency, and no delta or theta peak emerges (the signature of a fixed point that remains over-damped rather than approaching an oscillatory (Hopf) instability. *Right panel*) relative delta and theta power rise only modestly with severity (delta $\sim$25→31%, theta $\sim$12→16% over $G_{\mathrm{CS}} = 0 \to 3$) and do so as a smooth, broadband $1/f$-gain tilt (the low-frequency shoulder lifting slightly as the fixed point translates), *not* as a spectral peak. Thus even severe OCD produces a small, broadband low-frequency increase rather than a delta/theta rhythm: the circuit stays a stable, over-damped fixed point throughout, in contrast to the sharpening resonances of the Parkinsonian state (S20.2, S20.4).

#### S21 Robustness to the $D_1$:$D_2$ lesion ratio

The corticostriatal overdrive augments the cortico-$D_1$ weight by $G_{\mathrm{CS}}$ and the cortico-$D_2$ weight by $\rho\, G_{\mathrm{CS}}$, with $\rho = 0.7$ *inherited* from the van Albada–Robinson base ratio $\nu_{d_2 e}/\nu_{d_1 e} = 0.7/1.0$ rather than fitted (main text §2). This raises two questions: does the OCD elevation depend on this ratio, and in particular is a $D_1$*-selective* lesion ($\rho = 0$, direct pathway only) qualitatively different? We scan $\rho$ from 0 (fully $D_1$-selective) to 1 (balanced) and continue the fixed point over the overdrive $G_{\mathrm{CS}} \in [0, 6]$ in both directions (Fig. S14, Table S13). Three features hold across the whole range.

*A. The severity readout is ratio-independent.* The caudate/$D_1$ rate rises with $G_{\mathrm{CS}}$ for *every* $\rho$: because $\rho$ scales only the $D_2$ augmentation while $D_1$ always receives the full $G_{\mathrm{CS}}$, the readout $\phi_{d_1}(G_{\mathrm{CS}})$ is driven up regardless of the ratio. The OCD elevation therefore does *not* require augmenting $D_2$ in proportion, and a purely $D_1$-selective lesion reproduces it.

*B.* $D_1$*-selectivity maximizes the cortical rise.* The $D_1$-selective lesion ($\rho = 0$) produces the *largest* elevation of the (single, lumped) cortical population ($\phi_e : 12 \rightarrow 20.6\ \mathrm{s}^{-1}$), because augmenting the direct pathway alone maximally releases the thalamocortical loop; a balanced lesion ($\rho = 1$) leaves cortex near baseline as the direct and indirect augmentations offset (Fig. S14). In every case the cortical rate rises *less*, and *downstream of*, the caudate, the circuit basis for treating cortical hyperactivity as a loop consequence of the striatal lesion rather than its driver.

*C. The fast circuit stays monostable for all $\rho$.* Forward and reverse continuations coincide (no separation, hence no fold) at every ratio. Multistability is thus a property of the slow $G_{\mathrm{CS}}$ plasticity (§2), not of the fast circuit, and is independent of the lesion ratio.

Adopting $\rho = 0.7$ is therefore an inheritance from the base parameterization, not a load-bearing choice: the qualitative OCD picture (a graded, elevated, over-damped caudate fixed point with no fast-circuit multistability) holds across the entire $D_1$-selective-to-balanced range.

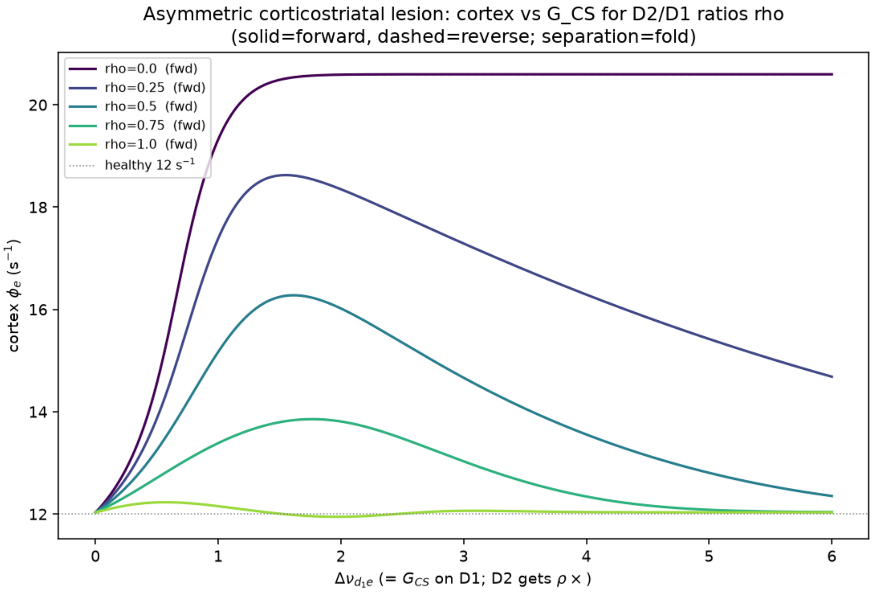

Figure S14: **The corticostriatal lesion drives the circuit for any $D_1$:$D_2$ ratio, without a fast fold.** Cortical rate $\phi_e$ versus the overdrive $G_{\rm CS}$ applied to $D_1$ (with $D_2$ receiving $\rho\, G_{\rm CS}$), for $D_2$:$D_1$ ratios $\rho = 0$ ($D_1$-selective) to 1 (balanced). Solid = forward continuation (from health), dashed = reverse (from the elevated end); their coincidence at every $\rho$ means the fast circuit is monostable (no hysteresis, no fold). Smaller $\rho$ (more $D_1$-selective) yields a larger cortical elevation; $\rho = 0$ saturates at $\phi_e \approx 20.6$ s$^{-1}$ while $\rho = 1$ stays at the healthy 12 s$^{-1}$.

| $\rho$ ($D_2$:$D_1$ augmentation ratio) | cortical $\phi_e$ at $G_{\rm CS} = 6$ (s$^{-1}$) | fast-circuit fold? |
|---|---|---|
| 0.00 ($D_1$-selective) | 20.6 | none (monostable) |
| 0.25 | 14.7 | none |
| 0.50 | 12.4 | none |
| 0.75 ($\approx$ adopted 0.7) | 12.0 | none |
| 1.00 (balanced) | 12.0 | none |

Table S13: **Cortical elevation and monostability across the $D_1$:$D_2$ lesion ratio.** For each ratio $\rho$, the cortical rate at the top of the clinical overdrive range ($G_{\rm CS} = 6$) and whether the fast circuit exhibits a fold (bistability). The adopted value $\rho = 0.7$ sits between the 0.5 and 0.75 rows ($\phi_e \approx 12.1$). The caudate/$D_1$ readout (not shown) rises with $G_{\rm CS}$ for every $\rho$; only the cortical projection depends on the ratio, and no ratio produces a fast-circuit fold.

### S22 Digitized trajectory data

Net (drug−placebo) Y-BOCS response used for calibration (13 SSRI arms; the two clomipramine arms are held out and used only for out-of-sample falsification).

| drug | trial | dose (mg) | $\bar{Y}_0$ | $\sigma_{Y_0}$ | net Y-BOCS improvement, $t(\text{wk}) : \Delta Y \pm \text{SE}$ |
|---|---|---|---|---|---|
| FLX | Tollefson et al.[106] | 20 | 23.6 | 5.5 | 1:0.30±0.63; 3:0.60±0.69; 5:2.50±0.75; 7:1.90±0.80; 9:2.70±0.89; 11:3.90±0.94; 13:4.00±0.96 |
| FLX | Tollefson et al.[106] | 40 | 23.5 | 5.5 | 1:-0.20±0.65; 3:0.60±0.77; 5:3.10±0.81; 7:3.50±0.81; 9:4.00±0.93; 11:4.30±0.92; 13:4.70±0.97 |
| FLX | Tollefson et al.[106] | 60 | 24.4 | 5.5 | 1:0.70±0.59; 3:1.10±0.68; 5:4.00±0.74; 7:4.80±0.81; 9:5.80±0.83; 11:7.20±0.87; 13:6.90±0.95 |
| CIT | Montgomery et al.[67] | 20 | 25.4 | 3.9 | 1:0.70±1.50; 3:1.20±1.50; 5:2.10±1.50; 7:2.60±1.50; 9:2.70±1.50; 12:2.80±1.50 |
| CIT | Montgomery et al.[67] | 40 | 25.4 | 3.9 | 1:1.00±1.50; 3:1.70±1.50; 5:2.80±1.50; 7:3.30±1.50; 9:3.30±1.50; 12:3.30±1.50 |
| CIT | Montgomery et al.[67] | 60 | 25.4 | 3.9 | 1:1.50±1.50; 3:2.70±1.50; 5:3.80±1.50; 7:4.80±1.50; 9:4.90±1.50; 12:4.80±1.50 |
| ESC | Stein et al.[105] | 10 | 27.0 | 5.0 | 1:-0.45±0.40; 2:-0.20±0.58; 4:0.56±0.75; 6:0.58±0.87; 8:1.28±0.95; 10:2.00±1.03; 12:2.97±1.09; 16:3.18±1.22; 20:2.31±1.36; 24:3.10±1.48 |
| ESC | Stein et al.[105] | 20 | 27.0 | 5.0 | 1:-0.08±0.40; 2:0.58±0.59; 4:1.44±0.73; 6:1.97±0.87; 8:2.34±0.95; 10:2.61±1.03; 12:3.68±1.08; 16:3.67±1.20; 20:3.32±1.34; 24:3.12±1.44 |
| PAR | Stein et al.[105] | 40 | 27.0 | 5.0 | 1:-0.01±0.40; 2:0.48±0.58; 4:1.66±0.73; 6:2.48±0.87; 8:2.52±0.95; 10:1.97±1.03; 12:3.21±1.09; 16:4.12±1.22; 20:3.35±1.34; 24:4.24±1.46 |
| PAR | Zohar et al.[46] | 37.5 | 25.5 | 4.0 | 2:1.50±0.98; 4:2.00±0.98; 6:3.00±0.98; 8:3.50±0.98; 10:4.00±0.98; 12:3.00±0.98 |
| SRT | Greist et al.[43] (pooled) | 50–200 | 23.8 | 5.3 | 1:0.60±1.10; 2:0.90±1.10; 4:1.40±1.10; 6:1.70±1.00; 8:1.90±1.00; 10:2.05±0.90; 12:2.16±0.80 |
| PAR | Kamijima et al.[107] | 20→40 | 24.3 | 4.5 | 1:0.60±1.10; 2:1.50±1.10; 4:2.90±1.10; 6:4.12±1.00; 8:4.35±1.00; 10:4.50±1.00; 12:4.65±1.00 |
| FLV | Hollander et al.[108] | 200 | 26.6 | 5.0 | 12:2.90±1.00 |
| CMI† | Greist et al.[45] | 226 | 26.2 | 5.2 | 1:0.50±1.00; 2:1.50±1.00; 3:2.50±1.00; 4:3.80±1.00; 5:5.00±1.00; 6:6.50±1.00; 7:7.50±1.00; 8:8.50±1.00; 9:9.50±1.00; 10:10.00±1.00 |
| CMI† | Zohar et al.[46] | 113 | 25.5 | 4.0 | 2:2.00±1.16; 4:3.00±1.16; 6:3.50±1.16; 8:4.00±1.16; 10:4.00±1.16; 12:3.00±1.16 |

$\bar{Y}_0, \sigma_{Y_0}$: reported baseline Y-BOCS mean and SD. Net = drug-arm minus placebo-arm improvement from baseline. Standard errors are as reported per timepoint where available (e.g. Stein et al.[105]), otherwise estimated from the reported endpoint SD and arm size (Montgomery et al.[67], Tollefson et al.[106]) and held constant across visits. †Clomipramine arms held out of the fit (out-of-sample falsification test). Values digitized from the source figures/tables and cross-checked against reported endpoints. Net trajectories are digitized from the reported (observed-cases / completer) figures rather than the LOCF primary-endpoint summaries, for consistency across trials; the two coincide in direction and magnitude but can differ by a fraction of a point at individual visits. Where a trial reports baselines by arm (Montgomery et al.[67], Stein et al.[105]), $\bar{Y}_0$ here is taken as the study/placebo reference baseline; the small per-arm departures (order 1 point) do not affect the net trajectories.
