## Supplementary material for "SSRI treatment of obsessive-compulsive disorder as motion across a bistable fold: A calibrated circuit–plasticity model of response, remission, and augmentation": One-page summary of the work

### A summary

Obsessive-compulsive disorder is usually treated first with an SSRI, and the response has a shape every clinician recognizes: it is partial, it takes weeks to appear, it improves little once the dose is high enough, and it helps some patients far more than others. When a first drug is not enough, treatment turns to augmentation with an agent from another class. These observations are well established; what is harder is to hold them together in one account that runs from the dose a patient takes to the change in their Yale-Brown score.

**What we did.** We built one model that follows an SSRI along that whole chain: how a dose sets serotonin-transporter occupancy and raises serotonin, how that perturbs the cortico-striatal-thalamic circuitry implicated in OCD, how the affected synapses remodel over weeks, and how all of this shows up in the recorded symptom score. Almost every constant is fixed from independent measurements; only two are fit to trial data. With those two, the model reproduces the response trajectories of six SSRIs across thirteen dose-arms, and predicts a trial it was never shown.

**What it revealed, in clinical terms.** In the model, OCD is a self-sustaining state the brain settles into and holds, separated from health by a threshold, so partial gains that do not clear it slip back. Treatment works by reshaping that landscape. If a drug carries a patient past the threshold, the OCD state is dissolved, not just suppressed, and full, medication-free remission becomes possible. If it only lowers the OCD state without erasing it, the patient improves, but the gain is held up by the medication and relapses when the drug is stopped. This is the model's account of the familiar split between a responder who relapses off drug and one who stays well: the second has crossed the threshold, the first has not.

**The severity ceiling, and where augmentation fits.** The same picture explains why the benefit depends on baseline severity. Mild-to-moderate patients can be carried across; for the most severe, raising serotonin reaches a safety ceiling before it can dissolve the state, so serotonin alone cannot get them there. Because the disorder is sustained by the plasticity balance rather than the serotonin level itself, agents that shift that balance in other ways can reach the same point by a different route. Antipsychotics, memantine, and ondansetron each enter at a distinct point, a mechanistic reason why combining classes can carry a severe patient across a threshold no single agent reaches alone. We offer this as a rationale for augmentation, not a claim of proven efficacy; for some of these agents the trial evidence is still limited.

**What is testable.** We asked whether the threshold reading earns its keep. A smoother model without a threshold fits the average trial curves equally well, so trial averages alone cannot decide between them. The two part ways only at the level of the individual patient: who reaches a durable off-drug remission, whether a treated group separates into two, and whether a patient's symptom ratings slow and grow more variable in the weeks before they shift. The last is within reach of the frequent symptom monitoring already used in practice, and the paper lays out the analyses that would test these predictions rather than treating the question as settled.

**Extensions.** The framework also points beyond SSRIs. Because exposure and response prevention (ERP) and transcranial magnetic stimulation (TMS) act on the same plasticity landscape by other routes, the model can be extended to include them. It could go further, to comorbid conditions, where each disorder is represented as its own elevated state and the states interact. The obstacle there is practical rather than conceptual: obtaining the data needed to calibrate such a model and validate its predictions.
